# Robust CD4^+^ CAR T cell Expansion Is Associated with Non-ICANS Neurotoxicities Following Ciltacabtagene Autoleucel

**DOI:** 10.1101/2025.10.28.25338924

**Authors:** Eric M. Jurgens, Sneha Mitra, Kevin Herrera, David Nemirovsky, Brenden Bready, Andriy Derkach, Kinga Hosszu, Devin McAvoy, Ross S. Firestone, Sridevi Rajeeve, Alexander M. Lesokhin, Neha Korde, Carlyn R. Tan, Hamza Hashmi, Hani Hassoun, Kylee Maclachlan, Urvi A. Shah, Malin Hultcrantz, Maximilian Merz, Francesco Maura, Sergio A. Giralt, Gunjan Shah, Heather J. Landau, Michael Scordo, Bianca D. Santomasso, Jae Park, Christina Leslie, Saad Z. Usmani, Karlo Perica, Sham Mailankody

## Abstract

Non-ICANS neurotoxicities (NINTs) are serious, atypical toxicities associated with ciltacabtagene autoleucel, a commercial chimeric antigen receptor (CAR) T cell therapy approved for relapsed/refractory multiple myeloma. Risk factors contributing to the development of NINTs are poorly understood. In a cohort of 109 patients, we identify predisposing risk factors and propose strategies to mitigate NINTs. We show that high peak absolute lymphocyte count is a strong NINT predictor which directly correlates with flow cytometry-based peripheral blood CAR T cell quantitation. The observed CAR lymphocytosis was polyclonal with a bias towards CD4^+^ CAR T cells rich in memory marker expression. We then identified CAR lymphocytosis associated CD4^+^ CAR T cell populations which exhibited increased inflammatory pathway gene expression. Finally, we characterize NINT associated CD4^+^ CAR T cell populations which are potential therapeutic targets for future exploration.

**One Sentence Summary:** Ciltacabtagene autoleucel associated non-ICANS neurotoxicities are driven by high CD4^+^ CAR T cell expansion exhibiting memory marker expression and upregulated inflammatory gene signaling pathways.

## INTRODUCTION

Ciltacabtagene autoleucel (cilta-cel) is a chimeric antigen receptor (CAR) T cell therapy targeting B-cell maturation antigen (BCMA) on myeloma cells. In relapsed/refractory multiple myeloma (RRMM), cilta-cel has demonstrated high response rates (>90%) (*1–3*) and progression-free survival (PFS) (*4*), which exceed those of alternative BCMA-targeted cell therapies including idecabtagene vicleucel (ide-cel) (*5*) and bispecific antibodies (BsAbs) (*6, 7*) (BsAbs) in comparable lines. Recent, long-term follow-up data at ≥5 years after cilta-cel treatment revealed ongoing complete responses in 33% of patients (*4*) suggesting curative potential in advanced RRMM. Additionally, cilta-cel has demonstrated an overall survival benefit in earlier lines of treatment (*8*) prompting evaluation as a frontline therapy in ongoing trials. The efficacy of cilta-cel is, however, tempered by serious toxicities.

CAR T cell therapy is associated with common toxicities observed with nearly all products encompassing different target antigens and diseases. These typical toxicities include but are not limited to cytokine release syndrome (CRS), immune effector cell (IEC)-associated neurotoxicity syndrome (ICANS), IEC-hemophagocytic lymphohistiocytosis-like syndrome (IEC-HS), IEC-associated hematotoxicity (ICAHT), and infections (*9*). In addition to these typical toxicities, BCMA CAR T cell therapy, in particular cilta-cel, is associated with uncommon, serious atypical toxicities. These toxicities include non-ICANS neurotoxicities (NINTs) (*10, 11*) such as cranial nerve palsy (CNP), peripheral neuropathy (PN), Guillain-Barré Syndrome (GBS), neuropsychiatric changes, and parkinsonism. IEC-associated enterocolitis (*12*), and CAR^+^ T cell lymphomas (*13, 14*) have also been reported. While these atypical toxicities have been observed occasionally following ide-cel, the respective events rates appear to be higher following cilta-cel (*12, 15–17*).

In the initial phase 1b/2 CARTITUDE-1 trial, delayed, atypical neurotoxicities distinct from ICANS were reported in 12/97 (12%) of patients treated with cilta-cel (*1, 11*). Five (5%) patients presented with parkinsonian features categorized as motor and neurocognitive treatment-emergent adverse events (MNTs) (*1*). Since this initial report, additional NINTs have been described including CNPs, GBS, and PN (*10*). The development of MNTs may be an on-target, off-tumor toxicity related to BCMA expression in the basal ganglia (*18*), however, the pathophysiology of other NINTs is not well understood. While high tumor burden (*10, 11*) and CAR T cell expansion (*10, 19*) have been implicated as predisposing risk factors, more data are needed to help predict which patients may develop NINTs.

Here we present retrospective analysis of 109 patients with RRMM treated with cilta-cel to identify risk factors associated with NINTs. We performed translational analyses of patient CAR T cells, including flow cytometry and cellular indexing of transcriptomes and epitopes by sequencing (CITE-Seq), to further elucidate the mechanisms and CAR T cell populations driving NINTs.

## RESULTS

### Patient Characteristics and Outcomes

In total, 109 patients with RRMM treated with standard of care (SOC) cilta-cel between July 21, 2022, and October 31, 2024, at Memorial Sloan Kettering Cancer Center were included. The median age at CAR T cell infusion was 66 years, 13 (12%) were Black, and 49 (45%) were female (Table 1). Patients were treated with a median 5 lines of therapy (LOT) including 3 patients previously treated with investigational CAR T cell therapy and 21 patients previously treated with a BsAb. Extramedullary disease (EMD) was present in 19 (24%) patients, high-risk cytogenetics in 69 (64%) patients, and high tumor burden in 10 (9.2%) patients prior to receiving cilta-cel. Additional patient characteristics are detailed in Table 1.

**Table 1:** Patient Characteristics by NINT.

| Characteristic | No NINT | NINT | P-Value |
| --- | --- | --- | --- |
| Peak ALC<sup>a</sup> |  |  | <0.001 |
| > 3.2×10<sup>3</sup>/μL | 20 (63%) | 12 (38%) |  |
| ≤ 3.2×10<sup>3</sup>/μL | 77 (100%) | 0 (0%) |  |
| Steroid Exposure<sup>b</sup> | 31 (84%) | 6 (16%) | 0.3 |
| Age at Cilta-cel Infusion | 66 (58, 72) | 65 (60, 75) | 0.6 |
| Race |  |  | >0.9 |
| Asian or Pacific Islander | 6 (86%) | 1 (14%) |  |
| Black or African American | 12 (92%) | 1 (7.7%) |  |
| Other or Multiracial | 5 (100%) | 0 (0%) |  |
| White or Caucasian | 70 (88%) | 10 (13%) |  |
| Sex |  |  | 0.4 |
| Female | 45 (92%) | 4 (8.2%) |  |
| Male | 52 (87%) | 8 (13%) |  |
| Tumor Burden<sup>c</sup> |  |  | 0.4 |
| Low | 68 (91%) | 7 (9.3%) |  |
| Intermediate | 4 (80%) | 1 (20%) |  |
| High | 25 (86%) | 4 (14%) |  |
| High-Risk Cytogenetics<sup>d</sup> | 63 (91%) | 6 (8.7%) | 0.3 |
| Clonal Hematopoiesis | 30 (91%) | 3 (9.1%) | >0.9 |
| Number of Lines of Prior Therapy |  |  | 0.057 |
| 4 or less | 34 (81%) | 8 (19%) |  |
| 5+ | 62 (94%) | 4 (6.1%) |  |
| Bridging Therapy | 74 (87%) | 11 (13%) | 0.5 |
| Alkylator Bridging Therapy<sup>e</sup> | 52 (93%) | 4 (7.1%) | 0.2 |
| Prior ASCT |  |  | 0.026 |
| ≤ 2 Years | 18 (75%) | 6 (25%) |  |
| > 2 Years | 67 (93%) | 5 (6.9%) |  |
| Prior BsAb | 20 (95%) | 1 (4.8%) | 0.5 |
| Prior CAR T-cell Therapy | 3 (100%) | 0 (0%) | >0.9 |
| Prior ADC | 10 (91%) | 1 (9.1%) | >0.9 |

NINT = non-ICANS neurotoxicity; ALC = absolute lymphocyte count; cilta-cel = ciltacabtagene autoleucel; ASCT = autologous stem cell transplant; BsAb = bispecific antibody; CAR = chimeric antigen receptor; ADC = antibody drug conjugate
<sup>a</sup>Absolute lymphocyte count observed within 30 days of cilta-cel infusion
<sup>b</sup>Steroid treatment within two weeks of cilta-cel infusion
<sup>c</sup>Tumor burden classified according to baseline bone marrow plasma cell (PC) infiltrate, serum M-spike, and involved serum free light chains (iFLC); high = any of the following - PC≥80%, M spike≥5g/dL, or iFLC≥5000mg/L; low = PC<50%, M-spike<3g/dL, iFLC<3000mg/L; intermediate = does not fit criteria for high or low (11)
<sup>d</sup>High-risk cytogenetics defined by the presence of 1q gain, t(4;14), t(14;16) or 17p deletion (86)
<sup>e</sup>Alkylator therapy included melphalan, cyclophosphamide (not including lymphodepletion), cisplatin

### Non-ICANS Neurotoxicities

We performed a retrospective chart review identifying all NINT diagnoses amongst the 109 patients treated with cilta-cel including MNTs, CNPs, GBS, and PN in accordance with the European Society for Blood and Marrow Transplant (EBMT) Practice Harmonization and Guidelines Committee guidelines (*10*). We also included atypical ICANS presentations, specifically delayed onset or prolonged course. All NINTs were evaluated by the treating oncologist and a consulting neurologist.

At a median follow-up of 17 months, 12 patients (11.0%) were diagnosed with a NINT. Of these 12 patients, 6 experienced 1 NINT and 6 experienced ≥2 NINTs (Figure 1A). In total, 22 NINTs were observed including 13 cranial nerve palsies (CNPs), 3 MNTs, 2 GBS, 2 PN, 1 delayed ICANS and 1 prolonged ICANS (Figure 1B; Table S1). The median time to onset of first NINT was 21 days (range 14-72 days; Figure 1C). At the time of last follow-up, 15/23 (65%) NINTs resolved with 7 ongoing cases amongst 6 patients including one patient with two ongoing NINTs (CNVII palsy and PN; Figure 1D). All 3 patients with MNTs had ongoing symptoms at the time of last follow-up; 247, 336, and 468 days respectively. Clinical data for each patient is further described in Table S1.

**Fig. 1.**
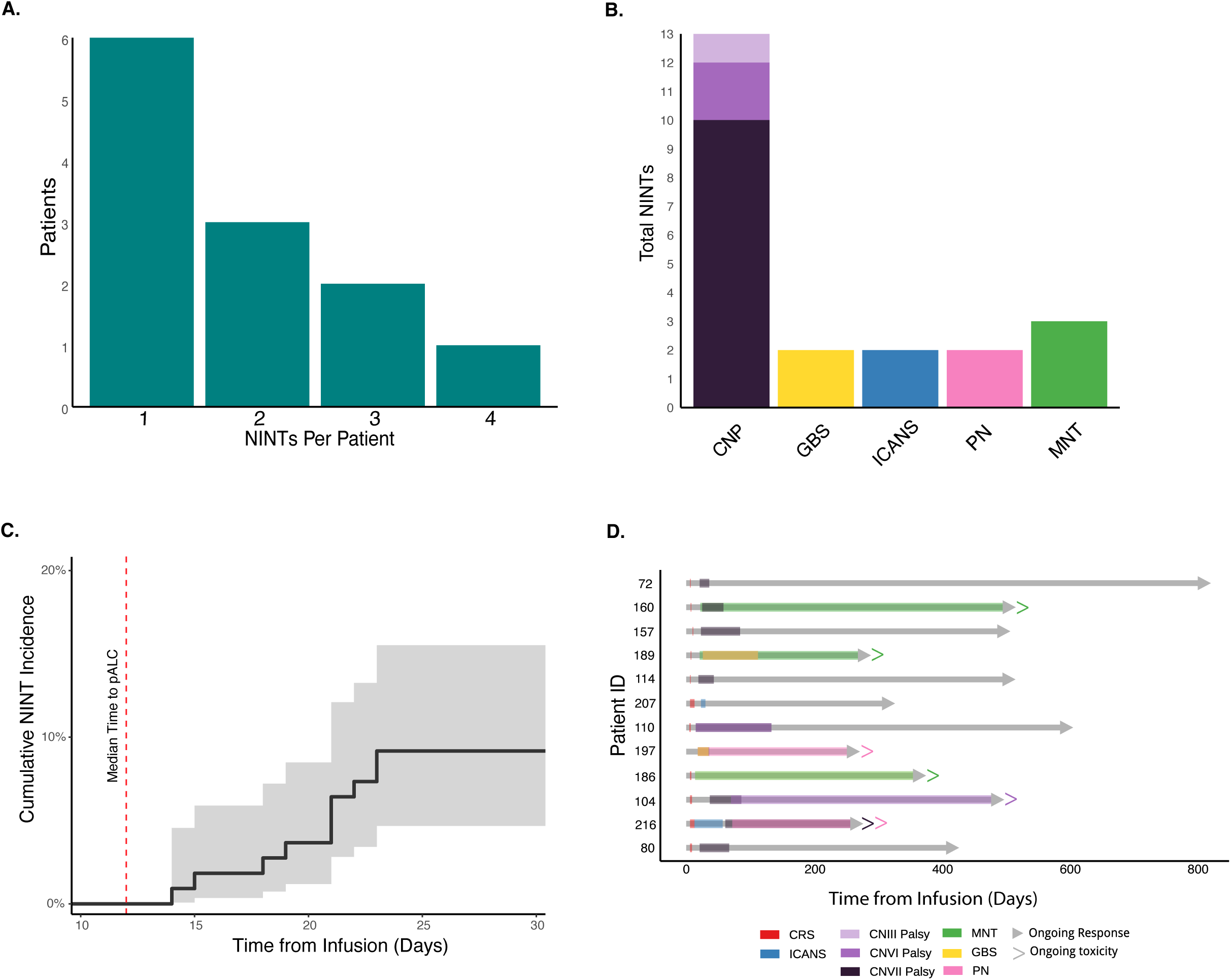
Distribution of NINTs amongst patients treated with cilta-cel (n=109). **(A)** Number of patients experiencing one or multiple NINTs. **(B)** Total events observed for each specific NINT amongst affected patients (n=12). ICANS included delayed presentation (n=1) and abnormally prolonged course (n=1). **(C)** Estimated cumulative incidence of first NINT within 30 days of cilta-cel infusion. Red dotted line indicates the median time to peak ALC (12 days) **(D)** Swimmer plot depicting the onset and duration of individual NINTs (colored coded on bottom) for each affected patient relative to cilta-cel infusion and duration of response (gray bar). NINT = non-ICANs neurotoxicity; CNP = cranial nerve palsy; GBS = Guillain-Barré Syndrome; ICANS = Immune effector cell-associated neurotoxicity syndrome; PN = peripheral neuropathy; MNT = movement and neurocognitive toxicities; CRS = cytokine release syndrome; CAR = chimeric antigen receptor.

### Lymphocytosis is a key risk factor for Non-ICANS Neurotoxicity

We sought to identify patient or disease characteristics that could select patients at risk of NINT. Pre-treatment baseline characteristics such as tumor burden (*10, 11*) and inflammatory signatures (*10, 19*) have been reported as potential contributing factors. However, we did not identify any association between NINT and EMD, tumor burden, or high-risk cytogenetics (Table 1). There was a weak association with a history of autologous hematopoietic cell transplantation (AHCT) and prior lines of therapy (LOT). In a univariate analysis, a recent history (≤2 years) of AHCT was associated with an increased risk of NINT (Odds Ratio [OR] 4.47, 95% Confidence Interval [CI] 1.22-17.2; p=0.025). Additionally, the median time since last AHCT was significantly shorter in patients with NINT compared to those without NINT (17 vs 62 months; p=0.025). In a multivariate analysis, fewer prior LOT (≤4) was associated with an increased risk of NINT (OR 4.82, 95% CI 1.07-25.5; p=0.04). Prior CAR T cell therapy, prior BsAb antibody, or use of bridging therapy was not associated with risk of NINT.

Patients with NINT compared to those without NINT exhibited a higher median peak absolute lymphocyte count (pALC), 7.50 vs 1.50 ×10^3^/µL. The median time to pALC was 12 days after cilta-cel infusion. Maximally selected Wilcoxon rank statistics determined a pALC of 3.2×10^3^/µL best identified patients at increased risk of developing NINT and thereby was defined as a “high pALC.” NINTs were observed in 12/32 (37.5%) patients with pALC>3.2×10^3^/µL compared to 0/77 (0.0%) patients with pALC≤3.2×10^3^/µL. In both a univariate and multivariate analysis, pALC>3.2×10^3^/µL was associated with a significantly increased risk of NINT (p<0.001 and p<0.001 respectively).

To validate high pALC as a significant NINT risk factor, we identified 51 additional patients treated with cilta-cel after the data cutoff. High pALC was observed in 20/51 (39%) patients of whom 8/20 (40%) developed NINTs. This closely reproduced the 37.5% NINT rate seen in our primary cohort. Furthermore, only 1/31 (3.2%) patients with pALC<3.2×10^3^/µL (0.86×10^3^/µL) developed a NINT (CNVII palsy). Observed NINTs in this cohort included 5 CNPs, 3 MNTs and 1 PN.

Since high pALC was strongly associated with NINT, we sought immune-related factors that might be predispose to NINT. Baseline inflammatory markers including ferritin, CRP, and interleukin (IL)-6 were not associated with an increased risk of NINT. Additionally, we performed extended cytokine analysis via Olink on baseline serum samples from a subset of patients (N=41; Table S2). Amongst 92 included cytokines, no significant differences were noted between patients with (N=12) and without NINT (N=29).

Genes implicated in clonal hematopoiesis (CH) such as *TET2*, *DNMT3A*, and *ASXL1*, have been reported in association with CAR T cell mediated toxicities including severe ICANS (*20*) and CRS (*21*). Additionally, disruption of CH associated genes may drive clonal CAR T cell expansion (*22–27*) and, in rare cases, CAR T cell lymphomas (*13, 14, 28, 29*). In our cohort, CH mutations were detected using a targeted next generation sequencing panel (NGS; MSK-IMPACT) (*30*) performed on bone marrow samples from 83/109 patients. CH mutations were common, observed in 40% of all patients, but not associated with an increased risk of NINT.

### High Peak ALC Risk Factors and Outcomes

Since high pALC was a strong predictor of NINT, we assessed if any baseline characteristics were associated with developing pALC>3.2×10^3^/µL. In a univariate analysis, only male sex was associated with high pALC (p=0.023); however, men were ultimately not at higher risk of developing NINT in our cohort (Table 1).

Interestingly, patients with high pALC compared to those without demonstrated a significantly longer PFS (Figure 2A). We hypothesized that inferior PFS was driven by patients with low pALC. This likely reflects poor CAR T cell expansion which is associated with worse outcomes following BCMA CAR T cell therapy (*31*). Using maximally selected Wilcoxon rank statistics we identified pALC>1.6×10^3^/µL as the optimal PFS cut point in this cohort (Figure 2B). Thus, patients with low pALC, ≤1.6×10^3^/µL, accounted for the observed inferior PFS.

**Fig. 2.**
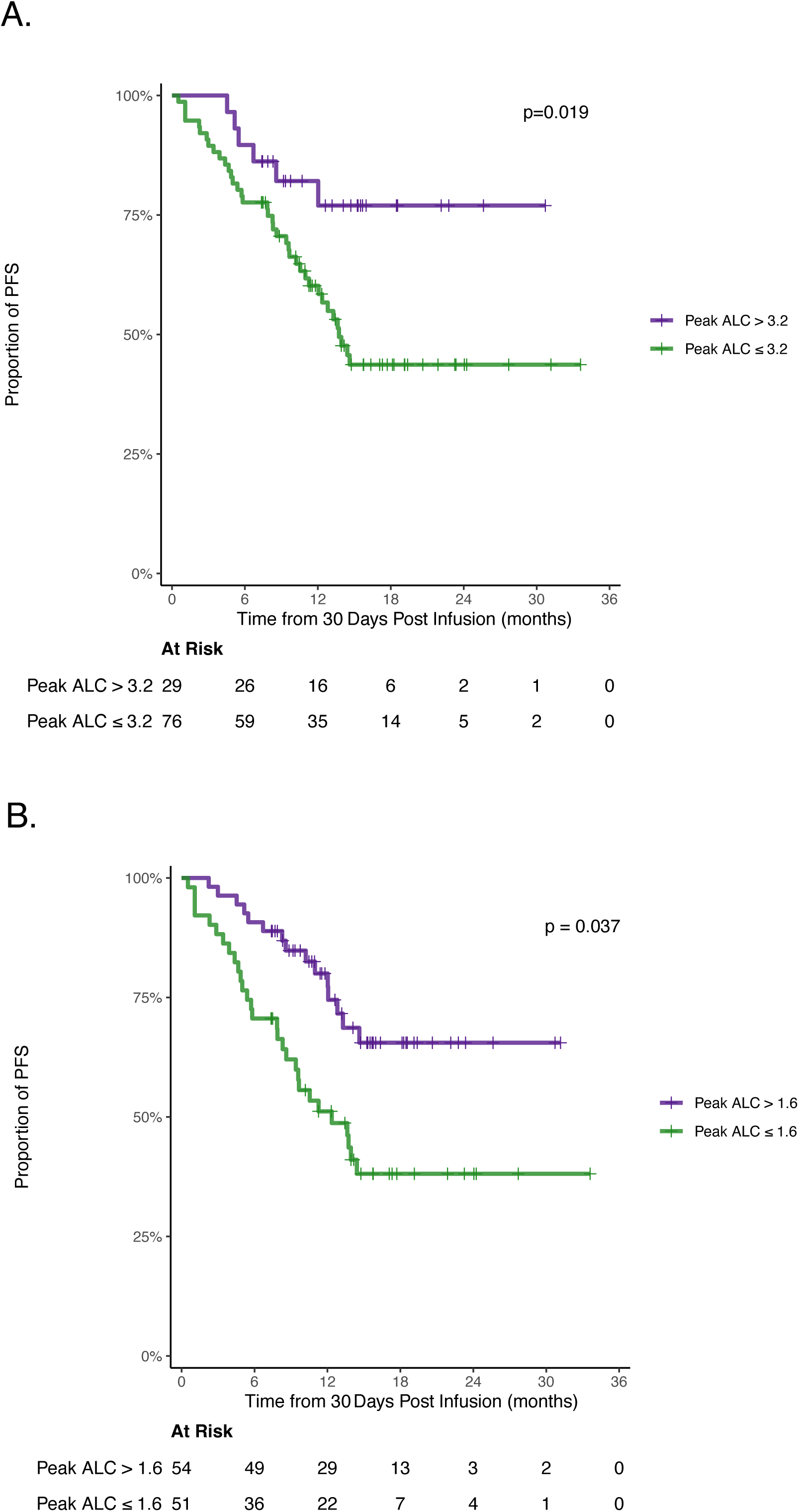
Peak ALC impacts progression free survival. **(A)** PFS of peak ALC groups separated by the identified NINT risk cutoff (3.2×10^3^/µL). **(B)** PFS of peak ALC groups separated by the identified optimal PFS cutoff (1.6×10^3^/µL). PFS = progression free survival; ALC = absolute lymphocyte count

There was no significant OS difference based on pALC though there was a trend towards longer OS in patients with pALC>1.6×10^3^/µL compared to ≤1.6×10^3^/µL (Figure S1A-B). Finally, pALC did not impact the depth of response (Table S3).

### CAR T cell Expansion Underlies High Peak ALC

To determine if high pALC was driven by CAR T cell expansion, we assessed CAR expression on peripheral blood mononuclear cells (PBMCs) using recombinant human BCMA, the target antigen for cilta-cel (Figure S2). T cells were additionally characterized using an 18-color T cell immunophenotyping panel and CAR frequencies and immunophenotypes were compared for patients with and without lymphocytosis at peak (days 7-14), one month (days 21-35), and late (days 60-90) timepoints (Figure S2-4; Table S4-5). Samples from 51 patients were available including 21 high pALC patients and 30 control patients (without high pALC). Of the 21 high pALC patients, 8 were also diagnosed with NINT.

First, we confirmed that clinically observed high ALC values directly correlated with absolute CAR T cells measured by flow at corresponding time points within 30 days from infusion. This association was strongest amongst patients with high pALC (Figure 3A). Thus, during the period of NINT onset, ALC was a reliable surrogate biomarker for robust CAR T cell expansion reflecting the development of CAR lymphocytosis (CL). CAR T cells accounted for a significantly greater proportion of total CD3^+^ T cells amongst CL patients during the peak expansion window (D7-14, 55.85% vs 10%, p=3.7e-05, Mann-Whitney U; Figure 3B). These patients also demonstrated significantly higher absolute CAR T cells including both CD4^+^ and CD8^+^ CAR T cells which persisted through D60-90 (Figure S5A-C).

**Fig. 3.**
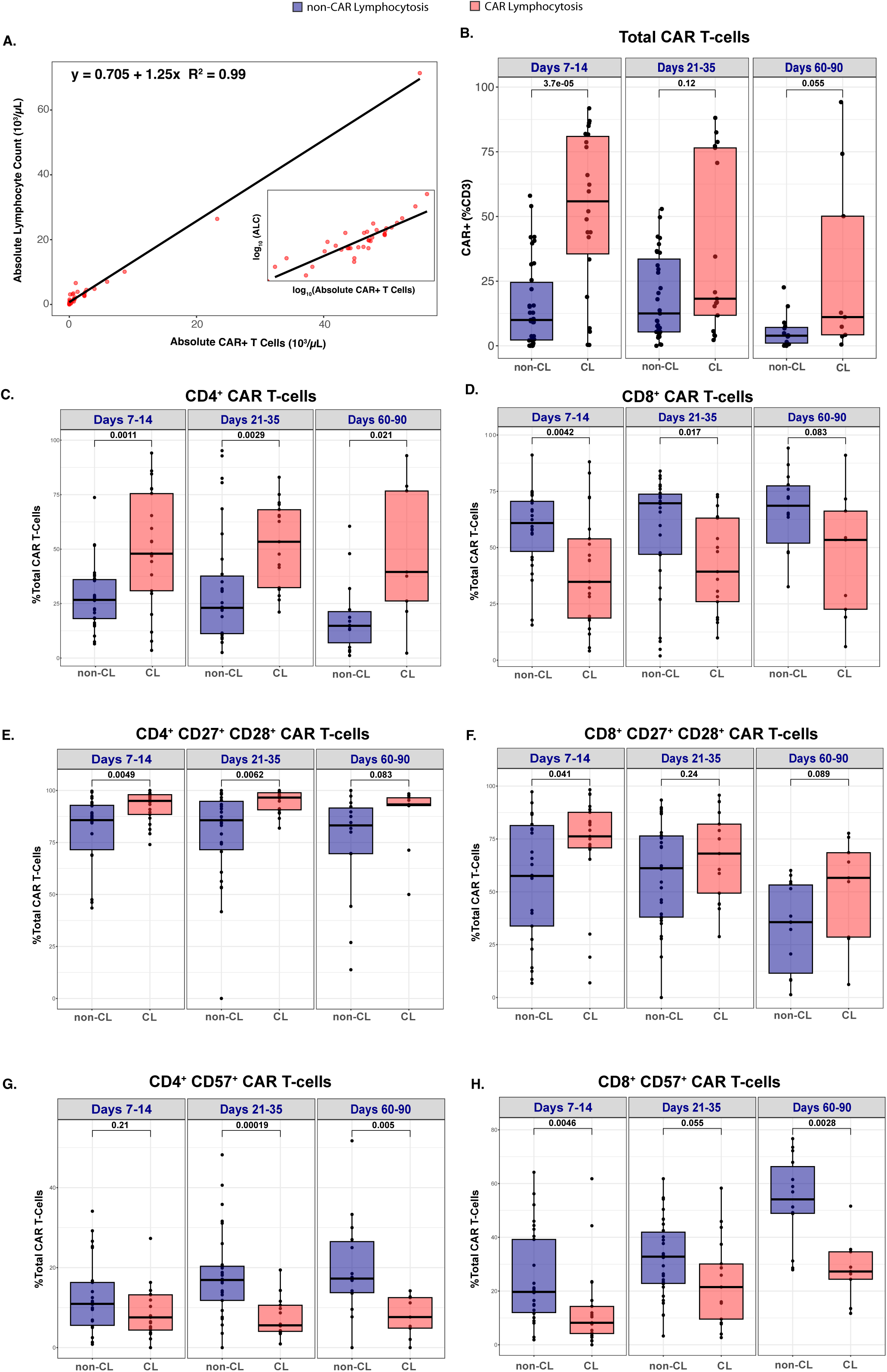
ALC correlates with absolute CAR T cells in patients with CL which exhibit bias towards CD4^+^ CAR T cells expressing memory markers. (A) Flow cytometry quantitation of absolute CAR^+^ T cells in CL patient peripheral blood mononuclear cells (PBMC) samples plotted against absolute lymphocyte count (ALC) at corresponding timepoints within 30 days of cilta-cel infusion (n=36). Subplot depicts ALC vs Absolute CAR^+^ T cells on log-log scale. (B) Box plot comparison of total CAR T cells (CAR^+^ CD3^+^ cells) in PBMC samples from non-CL (blue) vs CL patients (red) within Days 7-14 (n_non-CL_=34, n_CL_=22), Days 21-35 (n_non-CL_=31, n_CL_=17), Days 60-90 (n_non-CL_=15, n_CL_=9). (C-H) Box plot comparison of key CAR T cell populations in CL vs non-CL patients as a percentage of total CAR T cells within Days 7-14 (n_non-CL_=30, n_CL_=21), Days 21-35 (n_non-CL_=30, n_CL_=17), Days 60-90 (n_non-CL_=14, n_CL_=9) including (C) CD4^+^ CAR T cells, (D) CD8^+^ CAR T cells, (E) CD4^+^ CD27^+^ CD28^+^ CAR T cells, (F) CD8^+^ CD27^+^ CD28^+^ CAR T cells, (G) CD4^+^ CD57^+^ CAR T cells, and (H) CD8^+^ CD57^+^ CAR T cells. P values were calculated using Mann-Whitney U test and depicted above boxes. CAR = chimeric antigen receptor; ALC = absolute lymphocyte count; CL = CAR lymphocytosis

### CAR Lymphocytosis is Biased Towards CD4+ CAR T cell Expansion

CAR T cell expansion was CD4-predominant, with greater CD4^+^ bias in CL vs non-CL patients (i.e. D7-14: 47.9% vs 26.7%, p=0.0011; Figure 3C). Conversely, CD8^+^ CAR T cells comprised a significantly smaller proportion of total CAR T cells in CL vs non-CL patients (i.e. D7-14: 34.8% vs 60.9%%, p=0.0042; Figure 3D). Similarly, patients with NINT exhibited significant bias towards CD4^+^ CAR T cell expansion whereas patients without NINT were significantly biased towards CD8^+^ CAR T cell expansion (Figure S6A-B).

### CAR Lymphocytosis is Associated with Memory T cell Marker Expression

CAR T cell differentiation and memory formation were examined using flow cytometric immunophenotyping. Patients with CL exhibited a significantly greater proportion of memory associated CD27^+^ CD28^+^ CAR T cell phenotypes within both CD4^+^ (95.0% vs 85.8%; p=0.0049) and CD8^+^ (76.2% vs 57.5%, p=0.041) CAR T cell subsets at D7-14 (Figure 3E-F). Both CD27 and CD28 were also independently expressed in greater proportions of CD4^+^ and CD8^+^ CAR T cell subsets amongst patients with CL vs without CL (Figure S7A-D).

Consistent with decreased CD27/CD28 memory expression, non-CL patients demonstrated enrichment of CD8^+^ CAR T cells with a terminally differentiated effector memory T cell phenotype (CCR7^low^ CD45RA^+^ T_TEMRA_; Figure S8). Both CD8^+^ and CD4^+^ non-CL CAR T cells also accumulated expression of the senescence marker CD57 over time, starting at D21-35 for CD4 (16.9% vs 5.6%, p=0.00019) and D60-90 for CD8 T cells (54.1% vs 27.3%, p=0.0028; Figure 3G-H).

Thus, patients with CL demonstrated preferential expression of CD27^+^ CD28^+^ memory signatures in both the CD4 and CD8 compartments, whereas patients without CL accumulated CD8^+^ T_TEMRA_ and CD57^+^ senescent cells in both subsets, consistent with short-lived effector differentiation.

Despite these differences in memory/effector differentiation phenotypes, there were no major differences in populations expressing inhibitory or exhaustion markers including PD-1, TIM3, TIGIT, and CD39 in either the CD4^+^ or CD8^+^ subsets (Figures S9A-H).

### Single cell RNA and Protein Expression Analysis

To better understand underlying CAR T cell clonality, differentiation and heterogeneity, we performed CITE-seq with VDJ sequencing, on CD3^+^ CAR^+^ T cells isolated from 16 patients at three different time windows (D7-14, D21-30, D70-90). Of these 16 patients, 7 did not have CL (control), 5 had CL with NINT, and 4 had CL without NINT (Figure S10A).

Two-dimensional reduction of single-cell RNA data showed distinct populations of CAR T cells from control and CL patients (Figure 4A). Notably, no monoclonal populations were detected by VDJ sequencing at any timepoint, including in a single patient (labeled as Pt216) with marked CL (Figure 4B).

**Figure 4:**
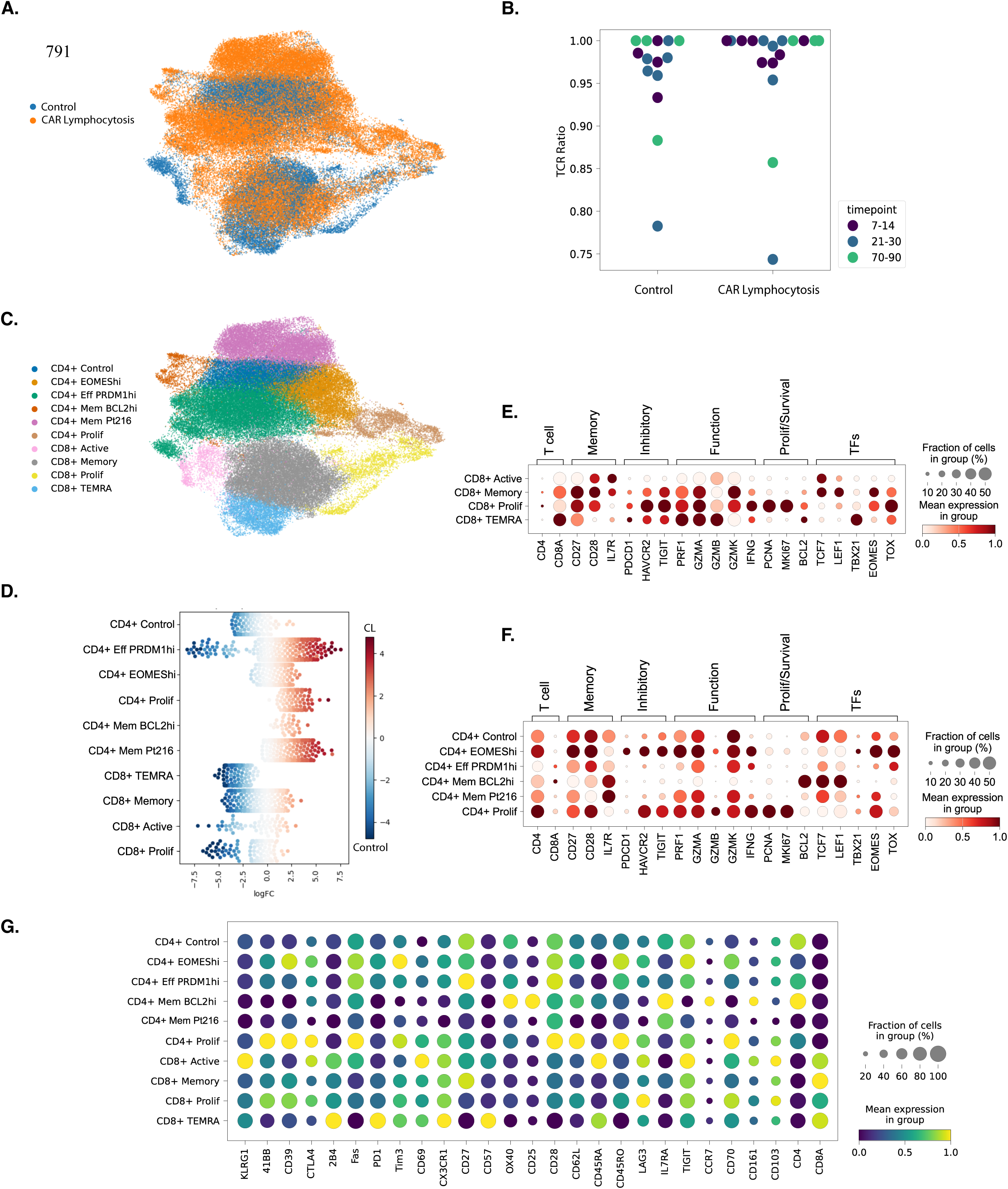
Defining Genetic Determinants of CAR Lymphocytosis. **(A)** UMAP projection of scRNA-seq data from CAR T cells from patients (n=16) treated with cilta-cel including patients without CAR lymphocytosis (control; n=7), CAR lymphocytosis with NINT (n=5), and CAR lymphocytosis without NINT (n=4) collected during three time windows; D7-14, D21-30, D70-90. Individual cells are color coded identifying CAR T cells from control vs CAR lymphocytosis patients. **(B)** T cell receptor (TCR) sequencing ratio to identify frequency and size of CAR T cell clones. **(C)** 10 distinct clusters were identified, color coded, and labeled according to defining features. **(D)** Abundance testing using Milo identifies clusters enriched in CAR T cells from patients with CAR lymphocytosis (red) vs control (blue). **(E)** Dot plot depicting CD8^+^ and **(F)** CD4^+^ CAR T cell gene expression by cluster and grouped by T cell phenotype or function. **(G)** Surface protein expression by CAR T cell cluster. CAR = chimeric antigen receptor; pALC = peak absolute lymphocyte count; NINT = non-ICANS neurotoxicity; UMAP = Uniform Manifold Approximation and Projection

### CD4^+^ Clusters Are Enriched in CAR T cells from Patients with CAR Lymphocytosis

K-means clustering defined ten unique CAR T cell populations consisting of six CD4 and four CD8-predominant populations (Figure 4C). Clusters were manually annotated according to distinguishing characteristics such as T cell phenotype, gene expression, or patient identity. To identify clusters enriched for control and CL populations, we performed abundance testing using Milo (*32*) (Figure 4D). Abundance testing revealed two populations, one in the CD4 subset and one in the CD8, which were enriched for control patients, termed “CD4^+^ control” and “CD8^+^ TEMRA,” respectively. Remaining clusters were either mixed or predominantly enriched for CL.

### CD8^+^ Clusters From Patients without CAR Lymphocytosis are Terminally Differentiated

In the CD8^+^ subset, four distinct groups were identified. CD8^+^ clusters were enriched in CAR T cells from patients without CL. Correlating with our flow cytometry analysis, these clusters exhibited terminally differentiated cytotoxic phenotypes including loss of memory markers *CCR7, TCF7, LEF1* and increased expression of cytotoxic markers *PRF1, GZMA, GZMB, GZMH,* and *NKG7* (*33–35*) (Figure 4E). Notably, inhibitory receptor expression was highest in an early proliferating cluster, CD8^+^ Prolif, with continued lower-level expression in a CD8^+^ TEMRA subset. This cluster demonstrated high CD45RA and CD57 expression at the protein level with low CD45RO and CCR7 protein expression (*36–38*) (Figure 4E).

### CD4^+^ Diversity from Patients with CAR Lymphocytosis

Consistent with our immunophenotyping, several CD4^+^ clusters were highly enriched for CL populations, including CD4^+^ Eff PRDM1^hi^, CD4^+^ EOMES^hi^, CD4^+^ Mem Pt216, CD4^+^ Mem BCL2^hi^, and CD4^+^ Prolif. These clusters universally expressed markers associated with memory programs including *CD28, CD27, SELL, TCF7,* and *LEF1* (*33–35*) (Figure 4F). However, they were defined by unique expression patterns of key regulators of T cell differentiation and survival, including *EOMES* and *TOX* in CD4^+^ EOMES^hi^, *PRDM1* (encoding BLIMP1) in CD4^+^ Eff PRDM1^hi^, and *BCL2* in CD4^+^ Mem BCL2^hi^.

Uniquely, the CD4^+^ Mem Pt216 cluster near exclusively contained CAR T cells from one patient with CL (Figure S10B-D). This patient exhibited a pALC of 71.4×10^3^/µL and developed multiple NINTs (Table S1). Bone marrow aspirate NGS revealed an *ASXL1 G1397S* mutation with variant allele frequency of 51.1% which was ultimately confirmed as a germline mutation. T cell receptor sequencing did not reveal a predominant clonal CAR T cell population.

Despite expression of memory programs, several CD4^+^ CAR T cell clusters from both the control and CL populations concomitantly expressed an atypical GZMK^+^ cytolytic program, with strong expression of *PRF1, GZMA, GZMK,* and *IFNG*, but not *GZMB*. Additionally, the CD4^+^ EOMES^hi^ and CD4^+^ Mem Pt216 clusters exhibited increased expression of gene sets (*39, 40*) associated with interferon (IFN) signaling pathways including IFN-γ and type-I IFNs, IFN-α and IFN-β (Figure S11-12).

Indeed, only the CD4^+^ Mem BCL^hi^ cluster did not have strong GZMK expression (Figure S13). This cluster, as well as CD4^+^ effector PRDM1^hi^, was further characterized by increased expression of tumor necrosis factor-alpha (TNF-α) signaling via nuclear factor-κB (NF-κB) associated gene sets (*39*) (Figure S11).

### NINTs are associated with CD4^+^ memory IL7Rα^hi^ CAR T cells

To determine which CAR T cell subset could be responsible for development of NINT, we repeated abundance testing for NINT vs non-NINT patients. Surprisingly, NINT enriched clusters highlighted CD4^+^ Mem Pt216 and CD4^+^ Mem BCL2^hi^ (Figure S14A-C), rather than the more strongly effector oriented GZMK subsets.

Both clusters exhibited high IL-7 receptor subunit alpha (IL-7Rα; CD127) protein and gene expression which is associated with memory T cell survival (*41–43*) including cilta-cel CAR T cell persistence (*44*) (Figure 4F, G). CD4^+^ Mem BCL2^hi^, uniquely, exhibited increased expression of *BCL2* encoding the anti-apoptotic protein BCL-2 (*45*) (Figure 4F). We repeated flow cytometric analysis on a subset of patients which revealed a significantly higher proportion of IL-7Rα^+^ CD4^+^ CAR T cells in patients with vs without NINT at D21-35 (55.4% vs 29.5%, p=0.0023; Figure S15). This population increased in relative abundance amongst NINT patients over time (Figure S15).

## DISCUSSION

NINTs are atypical and serious complications following cilta-cel and the exact pathophysiology remains to be fully elucidated. Consistent with other reports, we observed NINTs occurring in 11% of patients with a median onset of 21 days after cilta-cel infusion (*10, 19*). We show that robust CAR lymphocytosis (CL) is a highly sensitive marker for the development of NINTs. Furthermore, CL is biased towards CD4^+^ CAR T cell expansion enriched in memory marker expression. Finally, CD4^+^ memory IL7Rα^hi^ CAR T cells were enriched in affected patients and may be the pathologic population driving NINTs.

Contrary to other reports, tumor burden (*10, 11*) and baseline inflammatory markers (*10, 19*) were not associated with a significantly increased risk of NINT or CL in our analysis. The only baseline characteristics associated with NINT were a recent history of AHCT and ≤4 prior LOT. Patients included in our analysis had a median of 5 prior LOT and therefore additional studies are necessary to assess the NINT risk in patients treated with cilta-cel in earlier lines.

The strongest NINT risk factor was CL defined by pALC>3.2×10^3^/µL which accounted for every affected patient. NINTs and pALC shared a close temporal relationship, with median time to pALC preceding median time to first NINT by 9 days. ALC directly correlated with circulating CAR T cells and CD4^+^ CAR T cells accounted for the majority of CD3^+^ cells in patients with NINT. CL was driven by single or small clones even in one patient with a germline *ASXL1* mutation. We also did not see any association with pre-treatment clonal hematopoiesis and risk of CL or NINTs. Thus CL reflects polyclonal CAR T cell expansion which is not necessarily driven by pre-existing clonal mutations.

We next sought to elucidate phenotypic and genetic signatures driving CL to identify potential mechanisms contributing to NINTs. Our scRNA-seq analysis revealed CD4^+^ CAR T cell clusters associated with CL. These clusters all demonstrated upregulated inflammatory signaling pathways such as TNF-α signaling via NF-κB. This pathway is associated with CD4^+^ T cell activation, differentiation, and cytokine secretion implicated in the pathogenesis (*46, 47*) of and a potential therapeutic target for autoimmune diseases including MS (*48*). We also noted increased expression of autoimmunity associated pro-inflammatory gene signatures promoting IFN-γ (*49*) and type-I IFN (*50*) signaling.

Our analysis suggests multiple potential treatment approaches to prevent or treat NINTs after cilta-cel. Non-specific lymphoreduction using chemotherapeutic agents such as cyclophosphamide is one approach to mitigate these toxicities (*11, 18, 51, 52*). Encouragingly, early intervention in symptomatic patients may maximize the possibility of symptom stabilization or even reversal (*51, 52*). Based on our results, lymphoreduction, targeting a pALC<3.2×10^3^/µL, within the first month of CAR T cell infusion is an opportunity to reduce or abrogate the risk of NINT. However, as CL also correlated with superior PFS this may sacrifice response duration. In our cohort, inferior PFS observed amongst patients without CL was driven by patients with low pALC reflecting poor CAR T cell expansion. Thus, an optimal ALC range which maximizes response duration and minimizes toxicity may exist to guide lymphoreduction. An alternative approach includes treating inflammation with corticosteroids or targeting specific upregulated inflammatory signaling molecules such as IFN-γ. Studies evaluating the role of prophylactic dexamethasone (*53*) for patients with NINT are currently ongoing.

We then identified specific CAR T cell populations driving CL and NINTs. Among the CL enriched populations, the memory marker defined clusters, CD4^+^ Mem Patient 2 and CD4^+^ Mem BCL2^hi^, were NINT enriched. Due to the small sample size evaluated in this cohort, other CL enriched clusters should not be excluded as potential NINT drivers. In particular, CD4^+^ EOMES^hi^ is a compelling NINT culprit as CD4^+^ cytotoxic T cells have been reported in association with autoimmune neurological diseases (*54–58*). CD4^+^ cytotoxic T cells are also implicated in the pathogenesis of Parkinson’s disease (*59, 60*) and other neurodegenerative diseases (*61, 62*) drawing a direct corollary to NINTs especially MNTs. Furthermore, a similar CD4^+^ effector CAR T cell population was recently reported in CSF samples from three patients diagnosed with NINTs after cilta-cel (*44*). Thus, CD4^+^ cytotoxic CAR T cells are a suspicious, potentially neurotoxic population which should be considered in future studies with larger NINT populations.

The NINT enriched CD4^+^ memory clusters were distinguished from the other CD4 clusters by high IL-7Rα expression. IL-7 signaling via IL-7Rα activates the JAK/STAT pathway (*63*) and plays a critical role in memory T cell activation, proliferation, and persistence (*42, 43*). Augmented IL-7 signaling has been exploited to mitigate CAR T cell exhaustion (*44, 64*), increase expansion (*65*), and improve efficacy (*65*). A recent report comparing patients treated with cilta-cel versus ide-cel found longer CAR T cell persistence associated with cilta-cel which was enriched in CD4^+^ IL7Rα^+^ CAR T cells at late time points (*44*). Consistent with these findings, we also observed an increase in the relative abundance of CD4^+^ IL7Rα^+^ CAR T cells in patients with NINT over time. Thus increased *IL7R* expression may contribute to CL with robust expansion and persistence of memory CD4^+^ CAR T cells.

In addition to serving as a potential NINT marker, IL-7Rα opens avenues for intervention. The IL-7Rα targeted monoclonal antibody, lusvertikimab, has shown preliminary efficacy against ulcerative colitis (*66*) and IL7Rα^+^ T cell acute lymphoblastic leukemia (T-ALL) (*67*). Alternatively, targeting the downstream JAK/STAT pathway inhibits IL7Rα^hi^ or IL7Rα mutated T-ALL (*68–72*). Furthermore, in a recent report, two patients with cilta-cel associated MNTs demonstrated symptom reversal after treatment with ruxolitinib (*73*), a JAK1/JAK2 inhibitor. Finally, increased *BCL2* expression, as seen in the CD4^+^ Mem BCL2^hi^ cluster, provides another opportunity for intervention. BCL-2 inhibitors such as venetoclax have demonstrated therapeutic synergy with ruxolitinib by blocking IL-7/ IL-7Rα signaling in T-ALL(*68–72*). Thus IL-7Rα and associated downstream effectors may serve as markers for impending NINT as well as a therapeutic targets.

This study has several limitations. First, the small sample size limited the statistical power to identify significant differences between groups with respect to baseline characteristics and outcomes. Second, the retrospective design precluded uniform patient sample collection. Consequently, samples were not available for all patients at standardized time points. Future studies should employ a prospective design with pre-specified PBMC sample collection time points. Third, since we were limited by the availability of previously banked PBMCs, suitable samples for flow cytometry and CITE-seq were only available from a subset of this cohort. Finally, we did not have access to end-of-production CAR T cells or bag washes to identify pre-infusion product specific features that may portend NINT.

In summary, ALC is a convenient, widely available surrogate biomarker of CAR T cell expansion after cilta-cel. A pALC>3.2×10^3^/µL, deemed CAR lymphocytosis, is a strong predicator of NINT. CL was predominantly driven by CD4^+^ CAR T cells with increased expression of inflammatory signaling pathway associated genes which are potential targets to inhibit uncontrolled CL. Finally, CD4^+^ memory CAR T cells with upregulated *IL7R* and *BCL2* expression were most strongly associated with NINT warranting further investigation into this population as a marker and therapeutic target for NINT.

## MATERIALS AND METHODS

### Patient Clinical Characteristics, Outcomes, and Toxicities

This is a single-center retrospective study including all patients with RRMM treated with commercial, standard of care (SOC) cilta-cel between July 21, 2022 and October 31, 2024 at Memorial Sloan Kettering Cancer Center. The validation cohort included all patients with RRMM treated with SOC cilta-cel between November 1, 2024 and June 1, 2025. This study was approved by the Memorial Sloan Kettering Cancer Center Institutional Review Board. Patient samples were collected and analyzed in accordance with the Declaration of Helsinki.

Key baseline characteristics included demographics, cytogenetics, extramedullary disease (EMD), treatment history, clonal hematopoiesis, and tumor burden. Tumor burden was categorized as low, intermediate, or high based on prior reports incorporating bone marrow plasma cell percentage, free light chains, and M spike (*74*). Clonal hematopoiesis (CH) associated mutations were detected by NGS via MSK-IMPACT (*30*) on patient bone marrow samples where available. Patients with CH associated mutations (*75–77*) with variant allelic frequency ≥2% were classified as CH positive. Where available, NGS from CD138 selected cells was used to discriminate myeloma associated mutations from CH mutations.

Treatment responses were determined according to International Myeloma Working Group (IMWG) uniform response criteria (*78*). Bone marrow minimal residual disease (MRD) was assessed via multicolor flow cytometry (sensitivity: 10^-5^) (*78, 79*).

CRS and ICANS were graded according to the American Society for Transplantation and Cellular Therapy consensus grading system (*80*). NINTs included MNTs, CNPs, GBS, and PN in accordance with the European Society for Blood and Marrow Transplant (EBMT) Practice Harmonization and Guidelines Committee guidelines (*10*). Atypical ICANS presentations including delayed onset or prolonged course were also included. All NINTs were evaluated by the treating oncologist and a consulting neurologist.

### Olink Proximity Extension Assay (PEA)

The relative concentrations of 92 proteins were measured using the Olink Target 96 Inflammation panel, a proximity-based immunoassay in which paired oligonucleotide-labeled antibodies bind the target protein. Upon binding, the DNA tags hybridize and are extended by DNA polymerase to generate a unique DNA barcode for each protein, which is subsequently amplified by PCR and quantified by qPCR.

### Peripheral blood mononuclear cells (PBMC) Isolation from Whole Blood

PBMCs were isolated from EDTA-treated peripheral blood by Ficoll-Paque (Cytiva) density centrifugation, in SepMate tubes (StemCell Technologies), according to the manufacturer’s specifications. Briefly, whole blood was diluted 1:1 with PBS supplemented with 2% FBS (PBS+2% FBS). Ficoll was added to SepMate™ tubes, followed by careful layering of the diluted blood sample. Centrifugation was performed at 1200g for 20 minutes at room temperature (RT) with the brake on. The top layer containing enriched PBMCs was collected, washed twice in PBS+A2% FBS and resuspended for further experimentation, or cryogenically preserved in freezing medium that contains 10% DMSO.

### CAR T Cell Immunophenotyping by Spectral Flow Cytometry

Freshly isolated PBMCs were used to enumerate CAR-T cells in patient samples (IDMS-022 CAR-T Cell Screening Panel; Table S4). Cryogenically preserved cells were used for assessing T cell exhaustion and activation in patient samples as well as in apheresis and end-of-production CAR-T cell samples (IDMS T ExAct panel; Supplementary Table S5). For both types of samples, PBMCs were resuspended in PBS at 3 million cells/ml, incubated with Human TruStain FcX Fc receptor blocking solution (Biolegend) and Live/DEAD Fixable Blue Dead Cell Stain (Invitrogen) for 20 minutes at room temperature (RT), shielded from light. After washing in Flow Wash Buffer (FWB; RPMI 1640 no phenol red + 4% FBS + 0.01% sodium azide), cells were incubated with the appropriate antibody mix for 20 minutes at RT in the dark, using Brilliant Staining Buffer (BD) and CellBlox Blocking Buffer (Thermo Fisher Scientific). Following two washes, cells were fixed in 0.5% paraformaldehyde/PBS and immediately analyzed on a Cytek Aurora 5L flow cytometer (Cytek). Antibody concentrations were optimized by titration, with detailed information provided in the relevant Antibody Tables. Data analysis was conducted using FlowJo v10.10.0 according to the indicated schematic (Figure S2: CAR-T Cell Screening Panel, Figure S3: IDMS T Exact Panel). Mann-Whitney U test was used to calculate differences between CAR T cell groups at each timepoint.

### Cellular Indexing of Transcriptomes and Epitopes

Single-cell suspensions were prepared from cryopreserved peripheral blood mononuclear samples. Individual samples were labeled with Total Seq -C anti-human (or anti-mouse) hashtag oligonucleotide (HTO) antibodies (BioLegend), each uniquely barcoded per sample. After incubation, cells were washed three times with cold PBS + 2% BSA to remove unbound antibodies. Following staining, all samples were pooled in equal cell numbers to ensure balanced representation in downstream analysis. Pooled cells were then incubated with a panel of barcoded TotalSeq-C antibodies targeting surface proteins (BioLegend) to enable CITE-seq profiling.

Cells were counted and viability was confirmed (>85%) before loading onto the 10x Genomics Chromium. Cells were sequenced using 5’ single-cell RNA with paired cell surface proteins (CITE) and T cell receptor (TCR) sequencing in five samples. Each sample was individually preprocessed using Seurat v5(*81*) to retain cells with at most 10% mitochondrial reads, at least 500 and at most 6,000 gene transcripts. Clustering was then performed per sample using Louvain clustering in the Seurat framework to identify two broad clusters of CD4 and CD8 T cells. The five samples were then integrated using the scVI (*82*) and scANVI (*83*) pipeline where scVI was first run with the top 2,000 highly variable genes, 2 hidden layers, 30 dimensions of latent space, and negative binomial distribution for estimating the likelihood of gene expression.

After running scVI, scANVI was used with the previously defined sample-specific annotations of broad cell types (CD4 and CD8) to refine the integration. The scANVI latent representation was used to subcluster the CD4 and CD8 T cells. Two of the clusters with high mitochondrial reads and no paired TCR sequences were excluded. Cells with no CAR expression based on CITE-seq were also removed from the analysis. This resulted in a total of 80,569 CAR T cells with 6 clusters of CD4 T cells and 4 clusters CD8 T cells. Milo(*32*) was used to estimate differential abundance in cluster compositions between pairs of subpopulations: CL versus control, late versus early, and NINT versus no NINT. To this end, the pertpy(*84*) framework was used for running Milo where a K nearest neighborhood graph was created using 150 neighbors to generate overlapping neighborhoods of cells. A weighted version of the Benjamini–Hochberg method was applied to estimate spatial false discovery rate of the differentially abundant neighborhoods.

### Statistics

Determination of pALC was within the first 30 days of cilta-cel receipt, with all NINTs occurring after reaching pALC, for patients with NINTs. Optimal cutoffs for pALC were determined using maximally selected Wilcoxon rank statistics for NINTs and using maximally selected log-rank statistics for PFS. The p-values for comparisons between cutoff groups were approximated using maximally selected rank statistics for the respective outcome of comparison(*85*). Baseline characteristics between patients above and below the determined pALC were compared using the Wilcoxon rank sum test for continuous covariates and Pearson’s Chi-squared test along with Fisher’s exact test for categorical covariates. Fisher’s exact test was also used to calculate differences in response rates between patients above and below the pALC optimal threshold. Univariable logistic regression was used to evaluate the association between NINTs and baseline covariates. Multivariable logistic regression was used to evaluate the association between NINTs and pALC, adjusting for number of prior lines of therapy and steroid use within 2 weeks of cilta-cel infusion. Cumulative incidence of NINTs were estimated with disease progression and death as competing risks. OS and PFS were estimated using the Kaplan-Meier method, with median follow-up calculated via the reverse Kaplan-Meier method. The log-rank test was used to evaluate differences in OS and PFS between patients above and below the pALC value of 3.2×10^3^/µL, along with differences in OS between patients above and below the pALC value of 1.6×10^3^/µL.

## Data Availability

All data produced in the present study are available upon reasonable request to the authors

## Acknowledgments

Tasmin Farzana for assisting in patient samples acquisition and data organization

## Funding

National Cancer Institute (NCI) Core Grant P30 CA008748 (All authors).

NCI T32 training grant T32 CA9512-34 (EMJ)

ASCO Young Investigator Award (EMJ, RMF)

International Myeloma Society Young Investigator Award (EMJ, RMF)

Blood Cancer United Career Development Program, Scholar in Clinical Research (SM)

## Author contributions

Conceptualization: EMJ, SM, KP

Methodology: EMJ, SM, KP, DN, BB, AD, KH, SM, CL

Investigation: EMJ, SM, KP, DN, BB, AD, KH, SM, CL

Writing – original draft: EMJ, SM, KP

Writing – review & editing: All authors

## Competing interests

T.S - Honoraria: Roche-Genentech.

R.S.F. – Grants: ASCO YIA and the IMS.

A.M.L. – Grants: Novartis, BMS, Trillium Therapeutics, Pfizer, Janssen; Personal fees: Trillium Therapeutics, Janssen; Patent (number US20150037346A1) with royalties paid.

N.K. - Research funding: Amgen, Janssen, Epizyme, AbbVie; Consultancy: Clinical Care Options, OncLive, and Intellisphere Remedy Health; Advisory board: Janssen, MedImmune.

C.R.T. – Research funding: Janssen, Takeda. Personal fees: Physician Educations Resource and MJH Life Sciences; Advisory boards: Janssen and Sanofi.

H.H. – Grants: Celgene, Takeda, and Janssen.

H.H. - Advisory boards: Sanofi, BMS, and Janssen.

K.M. – Funding: Sebia, Binding site, and Siemens.

U.A.S. - Research support: Celgene/BMS and Janssen; Personal fees: MashUp MD, Janssen Biotech, Sanofi, BMS, MJH Life Sciences, Intellisphere, Phillips Gilmore Oncology Communications, i3 Health, and RedMedEd.

M.H. - Research funding: Daiichi Sankyo, Cosette Pharmaceuticals, GlaxoSmithKline, Abbvie, Beigene; Honoraria for consultancy/participated in advisory boards for Curio Science LLC, Projects in Knowledge, Intellisphere LLC, Bristol Myers Squibb, Janssen, and GlaxoSmithKline.

S.A.G. - Personal fees and advisory board: Actinium, Celgene, BMS, Sanofi, Amgen, Pfizer, GSK, Jazz, Janssen, Omeros, Takeda, and Kite.

G.L.S. - Research funding: Janssen, Amgen, BMS, Beyond Spring; serves on the data safety monitoring board (DSMB) for ArcellX; and receives research funding to the institution from Janssen, Amgen, BMS, Beyond Spring, and GPCR.

H.J.L. – Consultancy: Takeda, Genzyme, Janssen, Karyopharm, Pfizer, Celgene, Caelum Biosciences; Research support: Takeda.

M.S. – Consultancy: McKinsey & Company, Angiocrine Bioscience, Inc, and Omeros Corporation; Research funding: Angiocrine Bioscience, Inc, Omeros Corporation, and Amgen, Inc; Advisory boards: Kite, a Gilead company; Honoraria: i3 Health, Medscape, and CancerNetwork for CME-related activity.

S.Z.U. - Grants and personal fees: AbbVie, 404 Amgen, BMS, Celgene, GSK, Janssen, Merck, Mundipharma, Oncopeptides, 405 Pharmacyclics, Sanofi, Seattle Genetics, SkylineDX, and Takeda.

K.P. - Intellectual Property Rights: NexImmune; Professional Services and Activities: Pfizer, Inc.; RxCure LLC

S.M. - Consultancy: Evicore, Optum, BioAscend, Janssen Oncology, BMS, AbbVie, HMP Education, and Legend Biotech; Honoraria: OncLive, Physician Education Resource, MJH Life Sciences, and Plexus Communications. Research funding: Janssen Oncology, BMS, Allogene Therapeutics, Fate Therapeutics, Caribou Therapeutics, and Takeda Oncology.

## Data and materials availability

All data are available in the main text or the supplementary materials.

---

**Figure S1:**
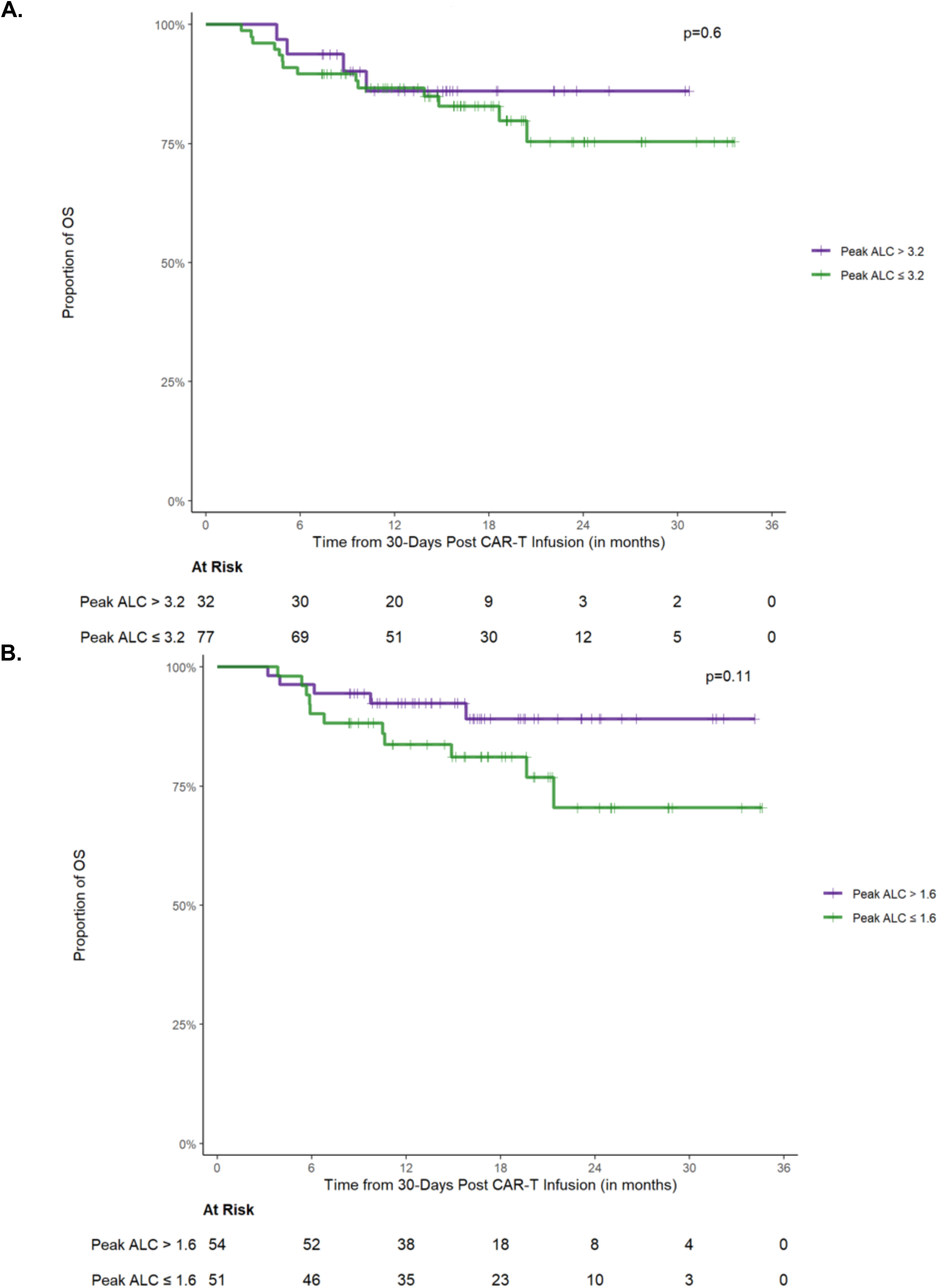
OS by pALC. **(A)** OS of peak ALC groups separated by the identified NINT risk cutoff (3.2×10^3^/µL). **(B)** OS of peak ALC groups separated by the identified optimal PFS cutoff (1.6×10^3^/µL). OS = overall survival; pALC = peak absolute lymphocyte count

**Figure S2.**
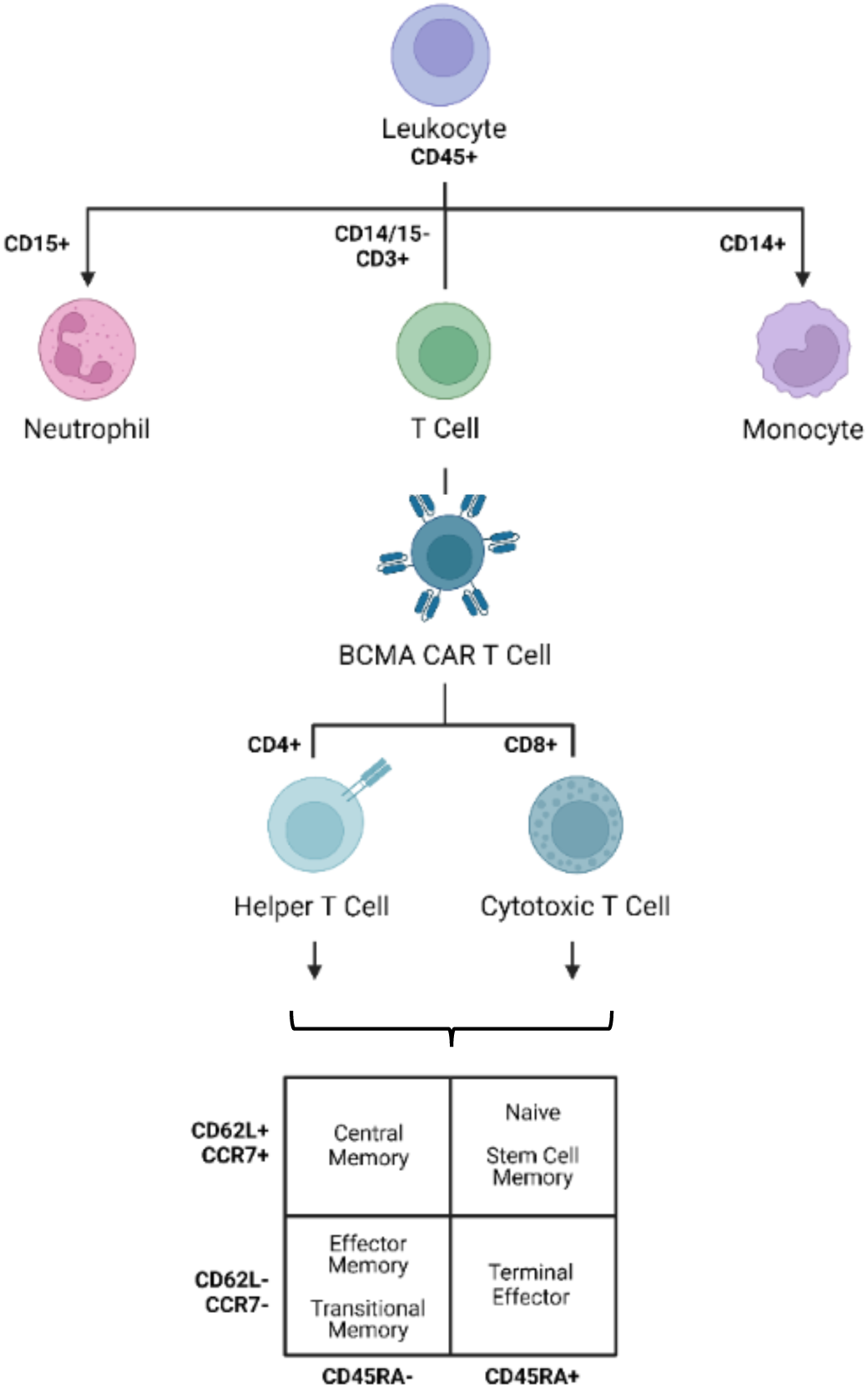
Analysis Scheme for CAR T cell Screening Panel. BCMA = B cell maturation antigen; CAR = chimeric antigen receptor

**Figure S3.**
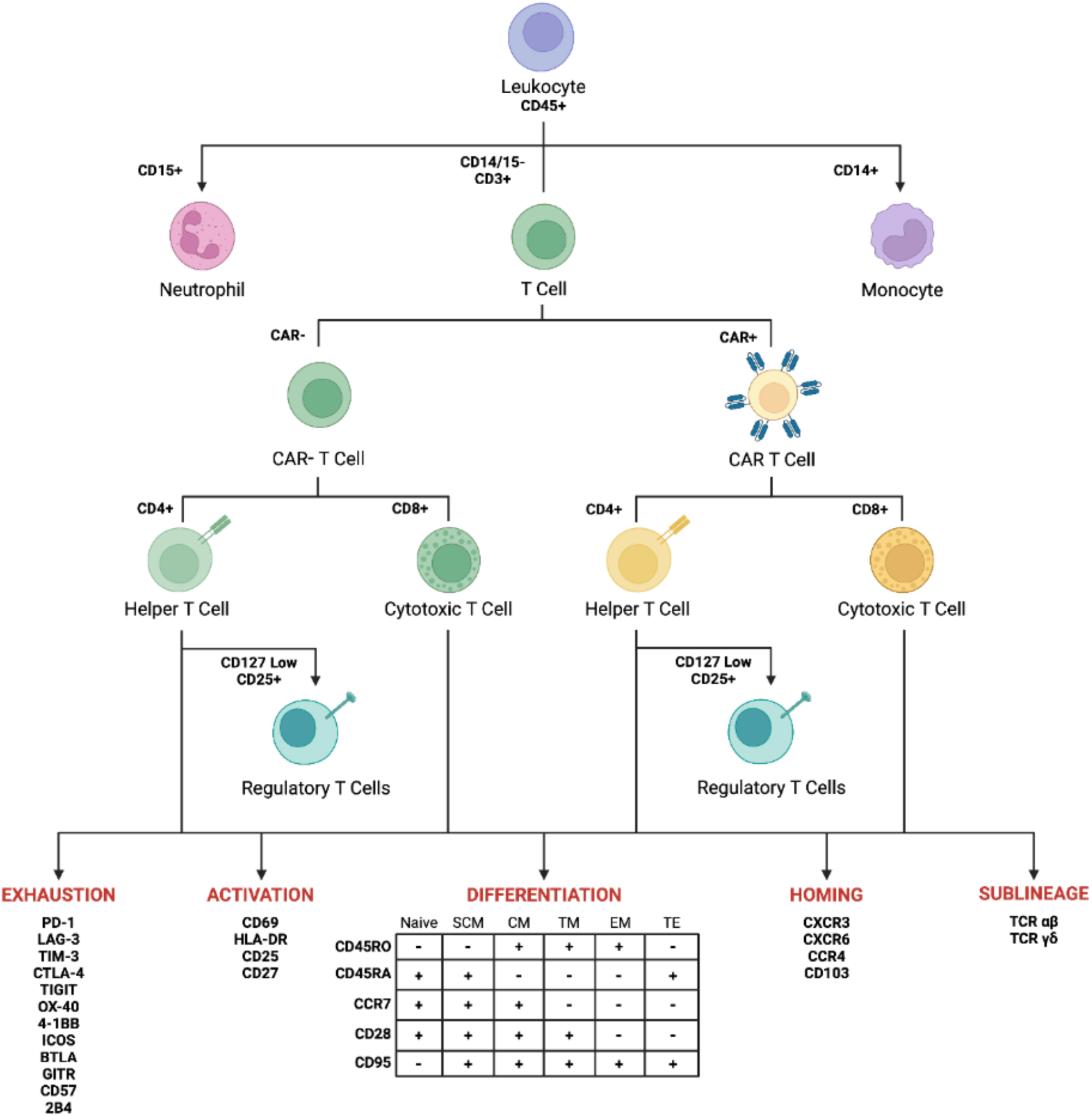
Analysis Scheme for T cell flow cytometry panel. CAR = chimeric antigen receptor

**Figure S4:**
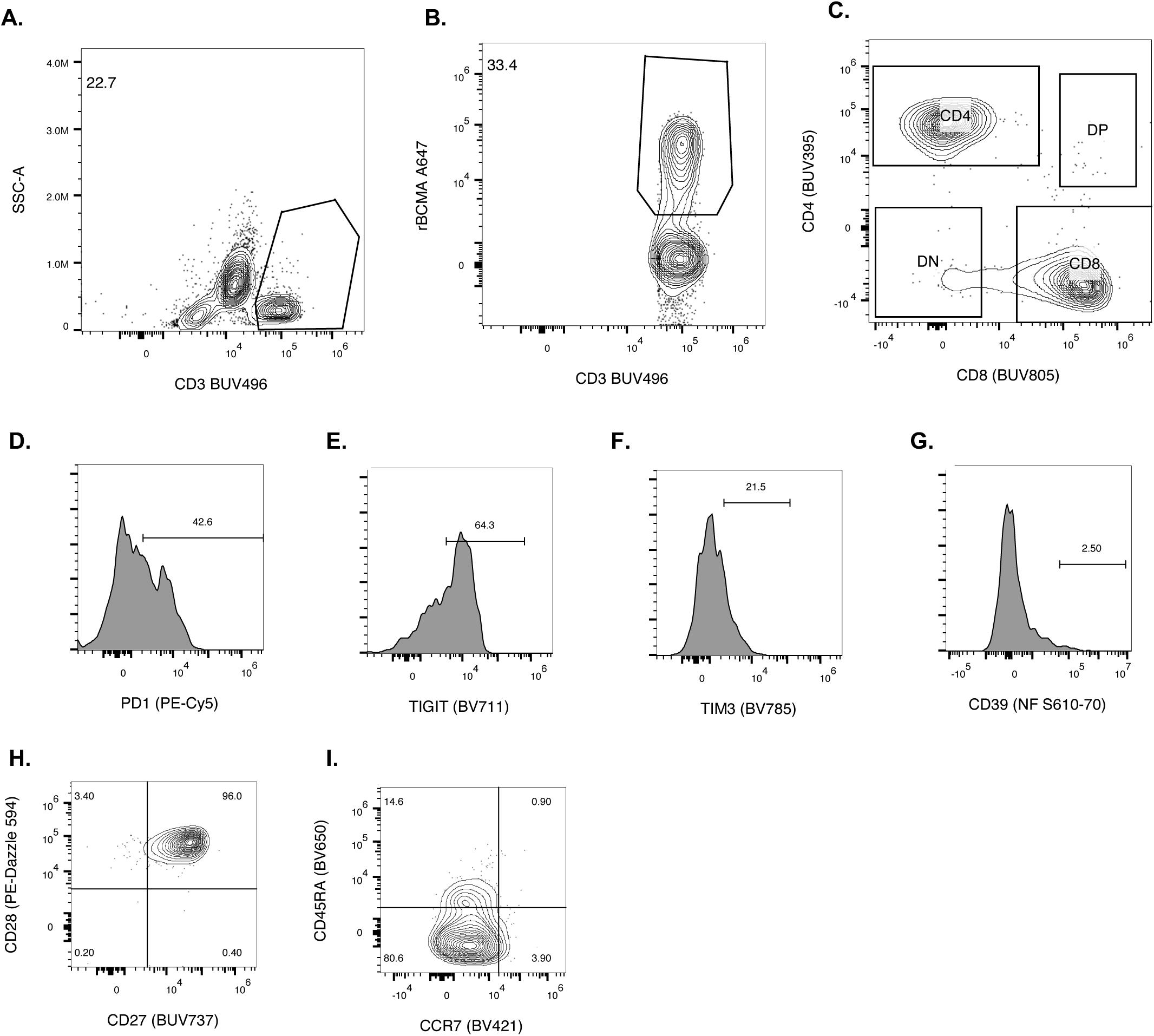
Flow cytometry gating strategy for patient peripheral blood CAR T cell immunophenotyping. **(A)** CD3 selection identified T cells **(B)** recombinant BCMA isolated CAR^+^ T cells which were than categorized based on **(C)** CD4 and CD8 expression. CAR T cells were further characterized by cells surface expression of **(D)** PD-1, **(E)** TIGIT, **(F)** TIM-3, **(G)** CD39, **(H)** CD27 and CD28, **(I)** CD45RA and CCR7.

**Figure S5:**
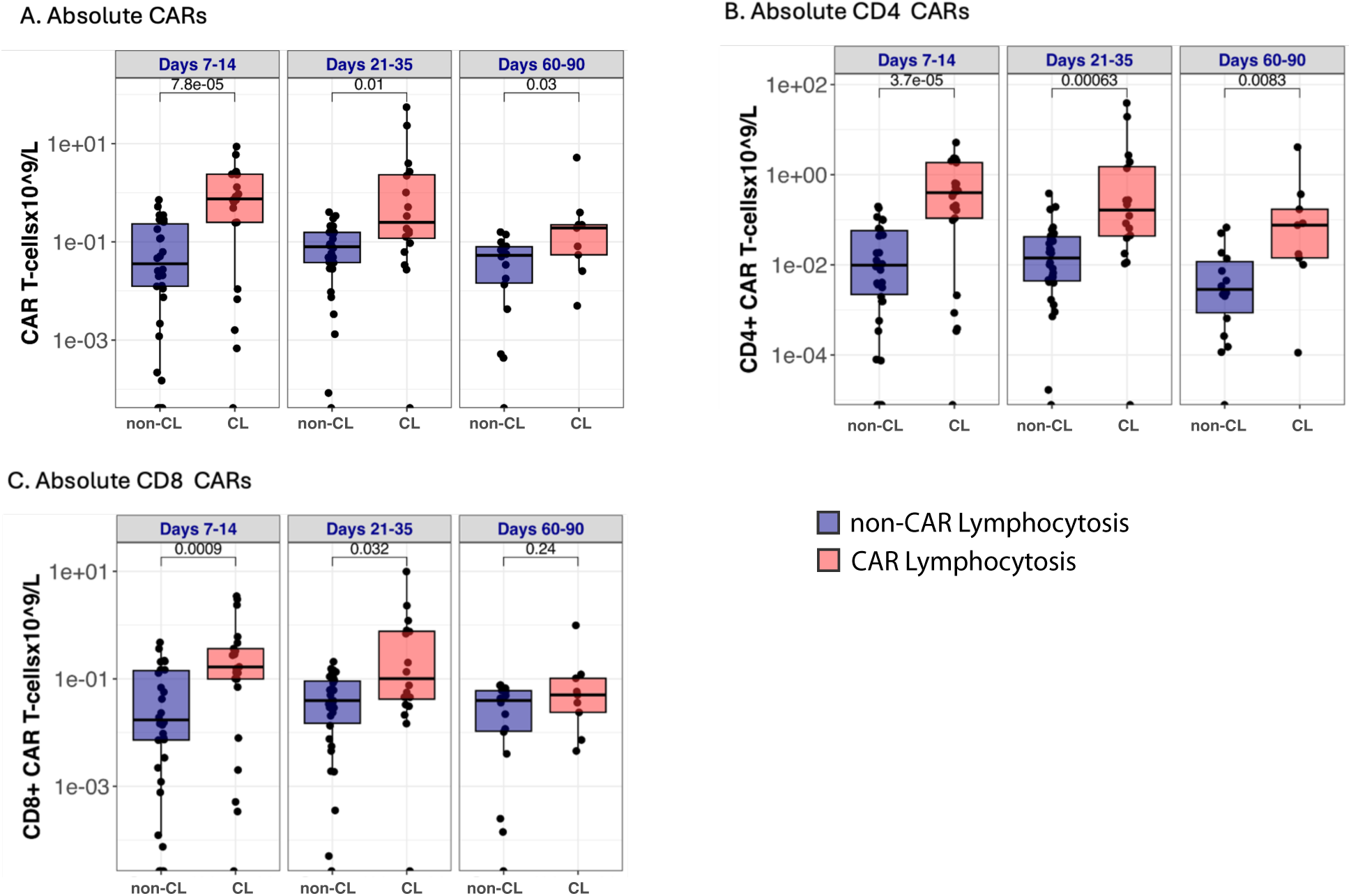
Patients with NINT exhibit bias towards CD4^+^ CAR T cells. **(A-C)** Absolute CAR T cells in in patients with CAR lymphocytosis (red) vs without CAR lymphocytosis (blue) during three time windows; Days 7-14 (n_non-CL_=30, n_CL_=21), Days 21-35 (n_non-CL_=30, n_CL_=17), Days 60-90 (n_non-CL_=14, n_CL_=9) including (A) Total CAR T cells, (B) CD4^+^ CAR T cells, and (C) CD8^+^ CAR T cells. P values were calculated using Mann-Whitney U test and depicted above boxes. CAR = chimeric antigen receptor; CL = CAR lymphocytosis

**Figure S6:**
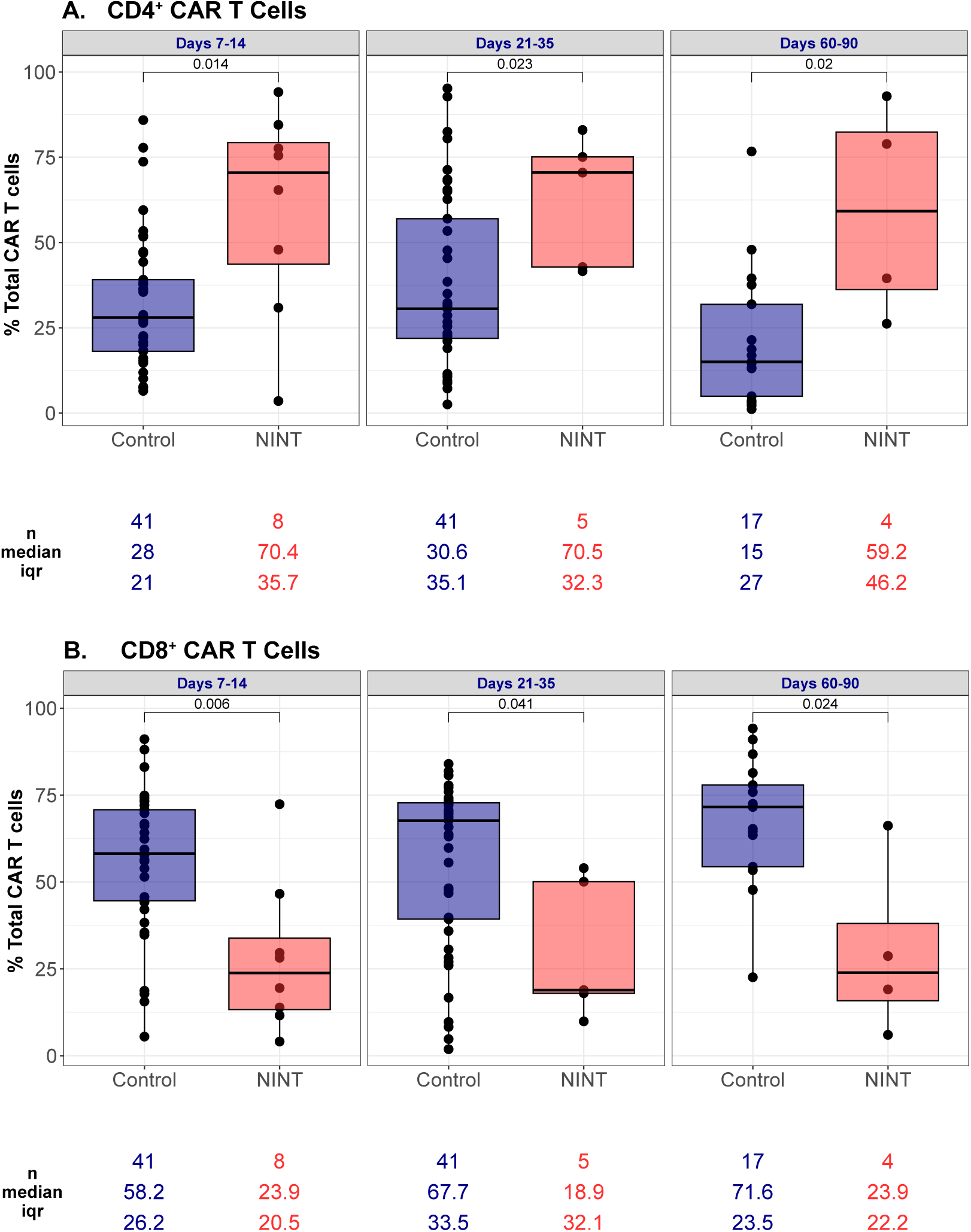
Patients with NINT exhibit bias towards CD4^+^ CAR T cells. **(A-B)** Proportion of peripheral blood CAR T cells in patients with NINT (red) vs without NINT (Control; blue) measured by flow cytometry during three time windows; Days 7-14 (n_control_=41, n_nint_=8), Days 21-35 (n_control_=41, n_nint_=5), Days 60-90 (n_control_=17, n_nint_=4) including (A) CD4^+^ CAR T cells and (B) CD8^+^ CAR T cells. P values were calculated using Mann-Whitney U test and depicted above boxes. CAR = chimeric antigen receptor

**Figure S7:**
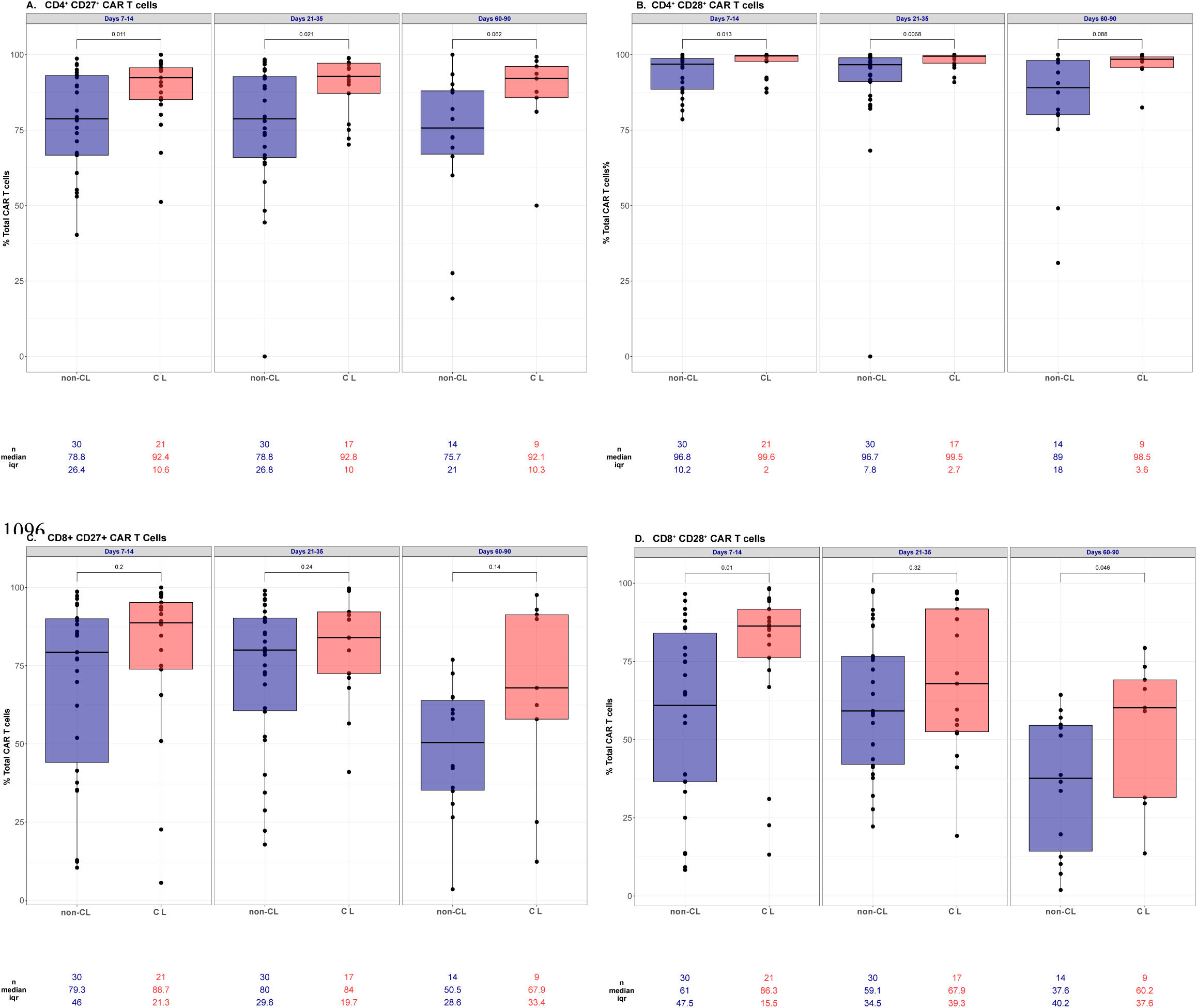
Patients with CL are enriched in CAR T cells expressing memory markers CD27 and CD28. **(A-D)** Proportion of peripheral blood CAR T cells in patients with CL (red) vs without CL (blue) measured by flow cytometry during three time windows; Days 7-14 (n_non-CL_=30, n_CL_=21), Days 21-35 (n_non-CL_=30, n_CL_=17), Days 60-90 (n_non-CL_=14, n_CL_=9) including (A) CD4^+^ CD27^+^ CAR T cells, (B) CD4^+^ CD28^+^ CAR T cells, (C) CD8^+^ CD27^+^ CAR T cells, and (D) CD8^+^ CD28^+^ CAR T cells. P values were calculated using Mann-Whitney U test and depicted above boxes. CAR = chimeric antigen receptor; CL = CAR lymphocytosis

**Figure S8:**
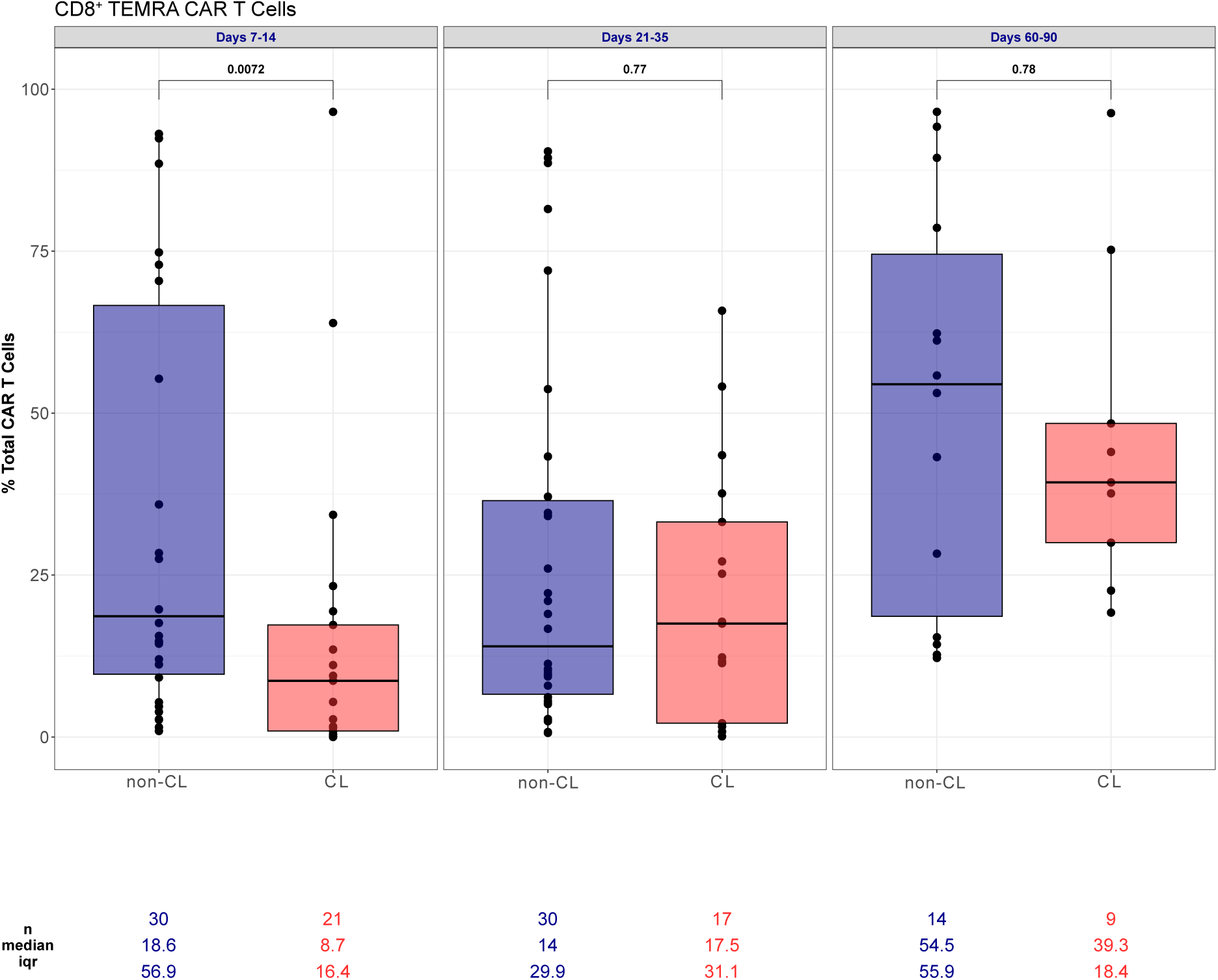
CD8^+^ TEMRA CAR T cells are enriched in patients without vs with CL. **(A)** Peripheral blood CD8^+^ TEMRA CAR T cells in patients with CL (red) vs without CL (blue) measured by flow cytometry as a percentage of total CAR T cells during three time windows; Days 7-14 (n_non-CL_=30, n_CL_=21), Days 21-35 (n_non-CL_=30, n_CL_=17), Days 60-90 (n_non-CL_=14, n_CL_=9). P values were calculated using Mann-Whitney U test and depicted above boxes. CAR = chimeric antigen receptor; CL = CAR lymphocytosis

**Figure S9:**
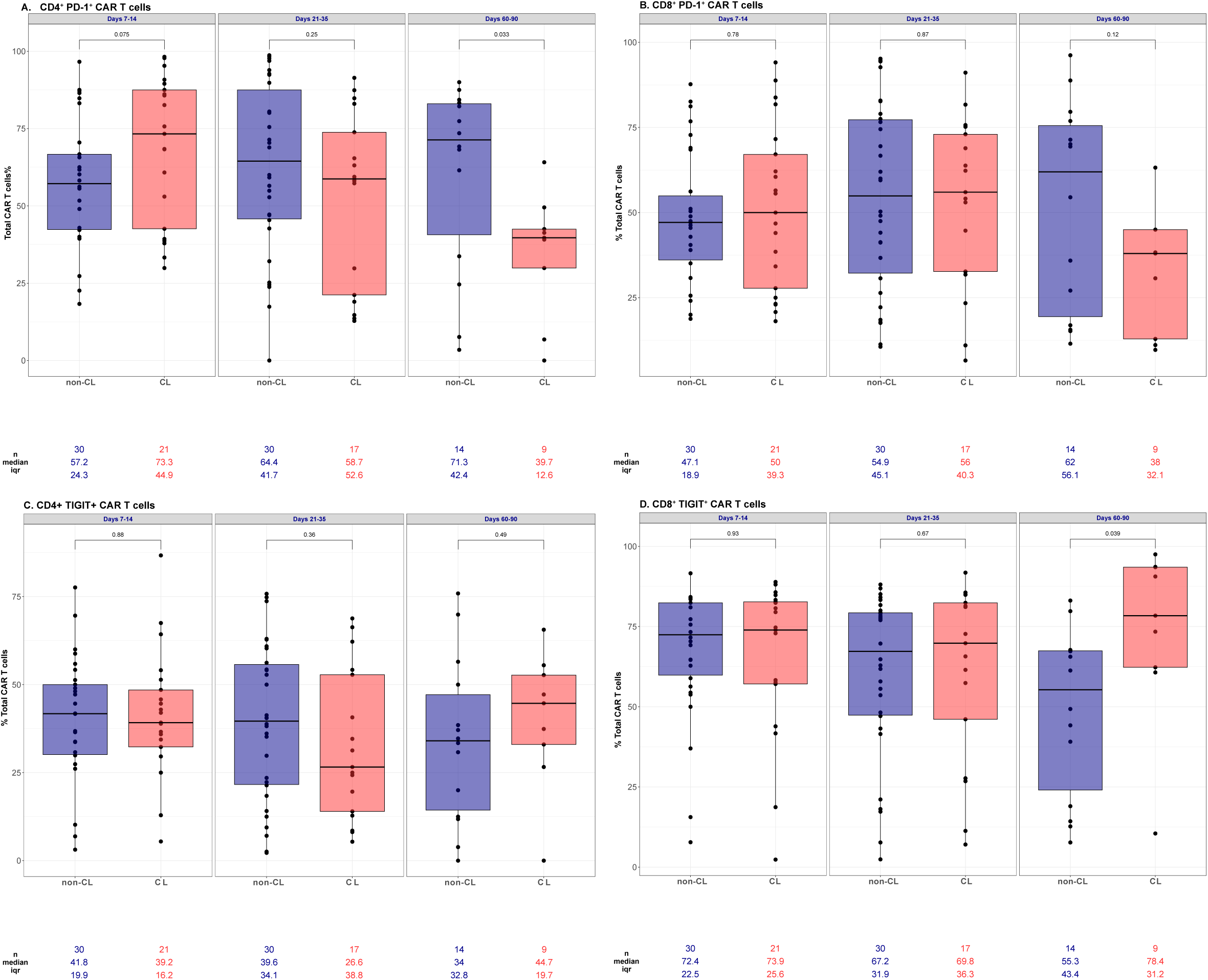
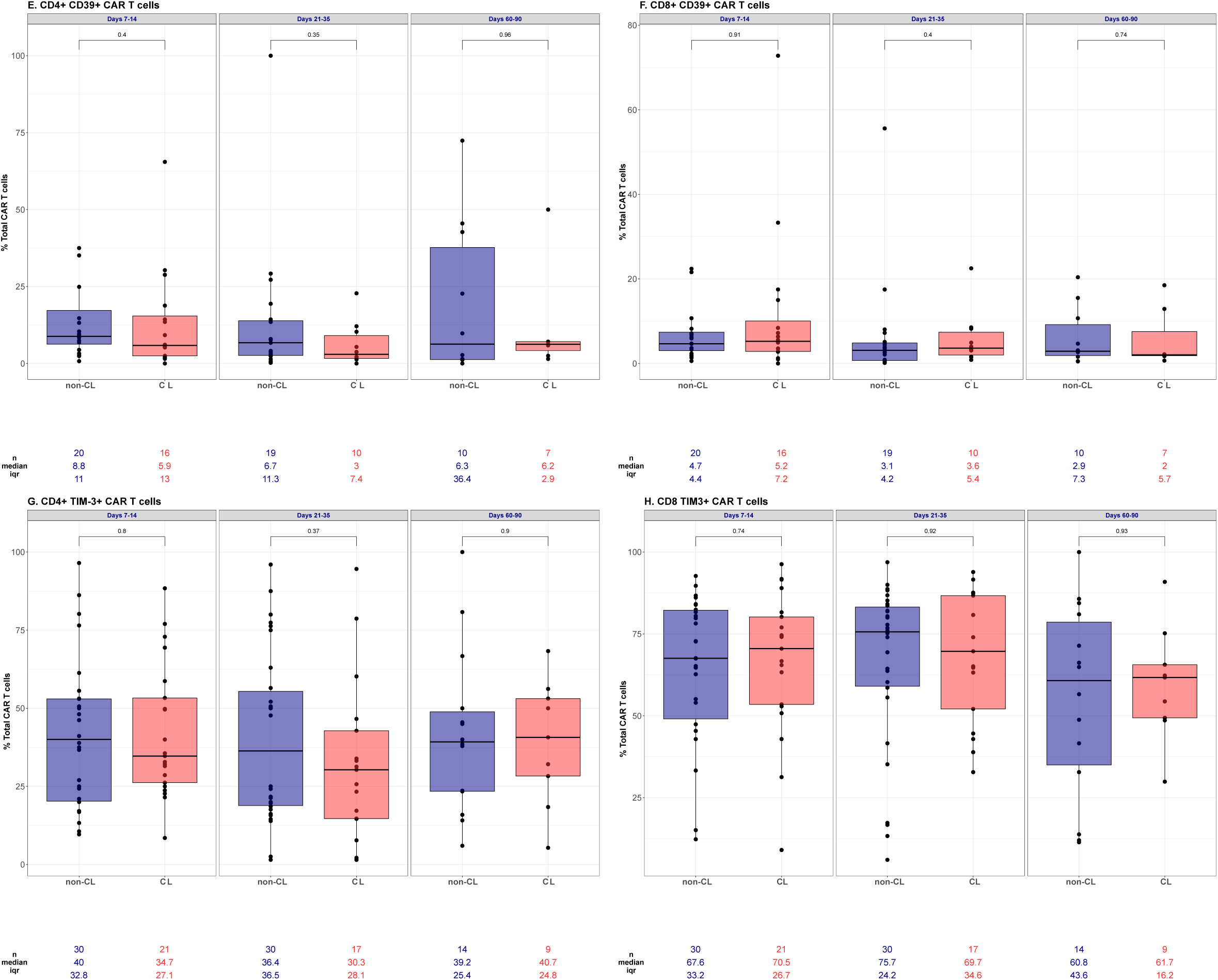
Inhibitory and exhaustion marker expression was similar amongst patients with and without CL. **(A-G)** Proportion of peripheral blood CAR T cells in patients with CL (red) vs without CL (blue) measured by flow cytometry during three time windows; Days 7-14 (n_non-CL_=30, n_CL_=21), Days 21-35 (n_non-CL_=30, n_CL_=17), Days 60-90 (n_non-CL_=14, n_CL_=9) including (A) CD4+ PD-1+ CAR T cells, (B) CD8+ PD-1+ CAR T cells, (C) CD4+ TIGIT+ CAR T cells, and (D) CD8+ TIGIT+ CAR T cells, (E) CD4+ CD39+ CAR T cells, (F) CD8+ CD39+ CAR T cells, (G) CD4+ TIM-3+ CAR T cells, and (H) CD8+ TIM-3+. For CD39+ CAR T cells: Days 7-14 (n_non-CL_=20, n_CL_=16), Days 21-35 (n_non-CL_=19, n_CL_=10), Days 60-90 (n_non-CL_=10, n_CL_=7). P values were calculated using Mann-Whitney U test and depicted above boxes. CAR = chimeric antigen receptor; CL = CAR lymphocytosis

**Figure S10:**
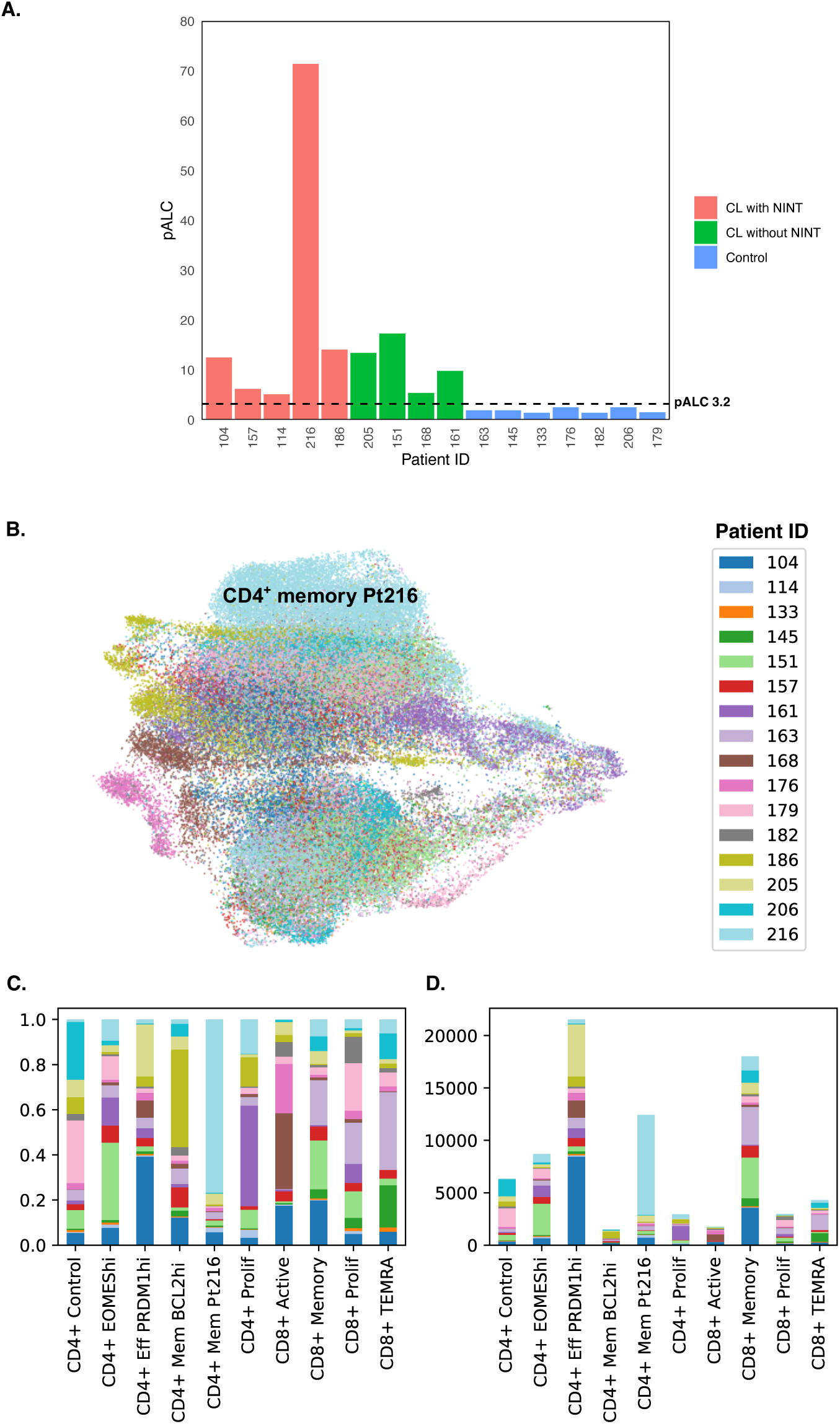
Distribution of CAR T cells from individual patients included in CITE-seq analysis. **(A)** Individual patients arranged on x axis with pALC depicted on y axis, patients are grouped according to history of CL with NINT (red), CL without NINT (green), or neither CL nor NINT (control; blue). **(B)** UMAP projection color coded identifying CAR T cells from individual patients (legend on right). CD4^+^ Mem Pt216 cluster is highlighted demonstrating near complete composition from patient 216. **(C-D)** Proportional and absolute measurement of CAR T cells from individual patients in each cluster. CAR = chimeric antigen receptor; pALC = peak absolute lymphocyte count; NINT = non-ICANS neurotoxicity; UMAP = Uniform Manifold Approximation and Projection

**Figure S11:**
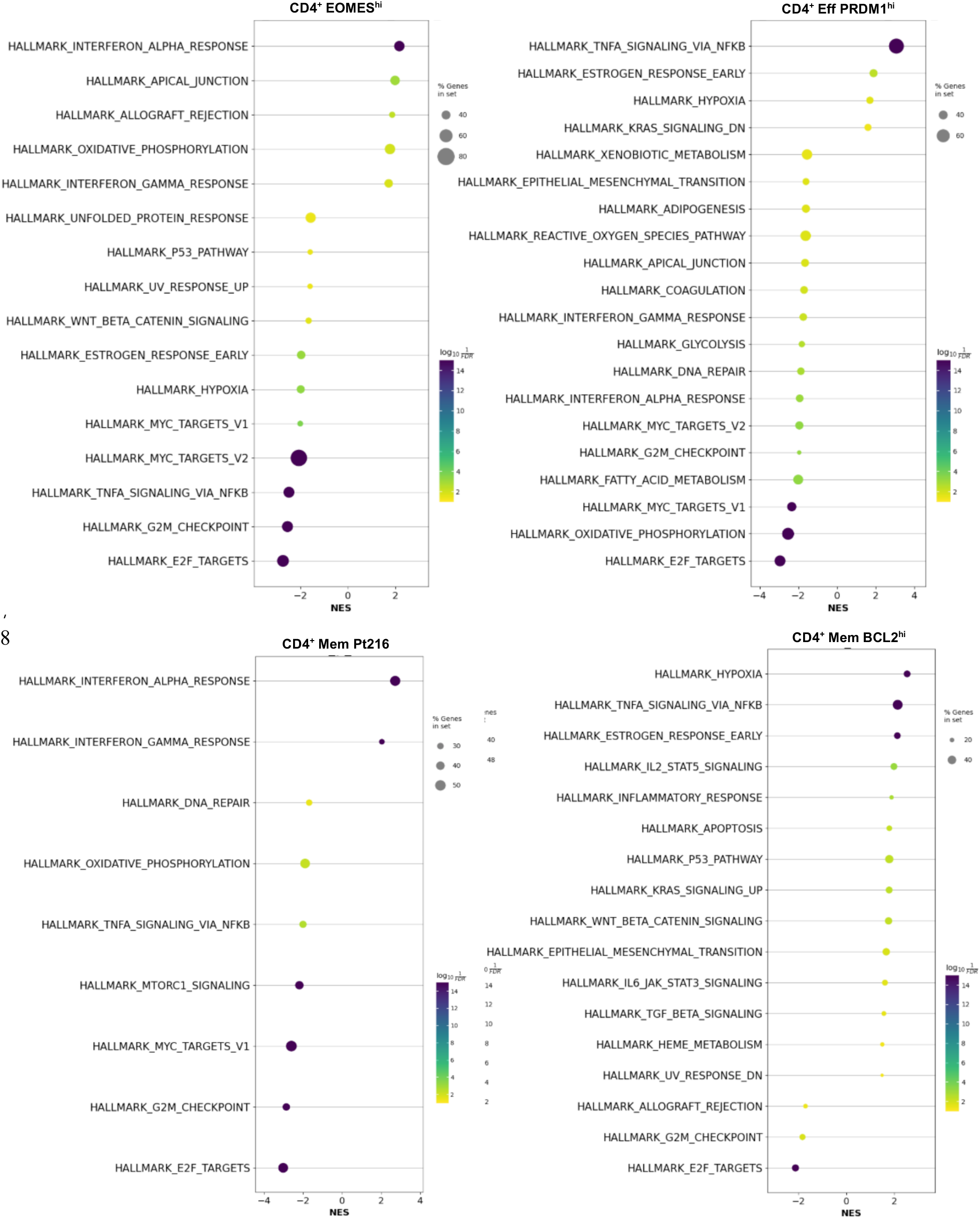
Hallmark gene set analysis. Hallmark gene sets associated with scRNA-seq defined CD4^+^ CAR T cell clusters. scRNA-seq = single cell RNA sequencing; CAR = chimeric antigen receptor

**Figure S12:**
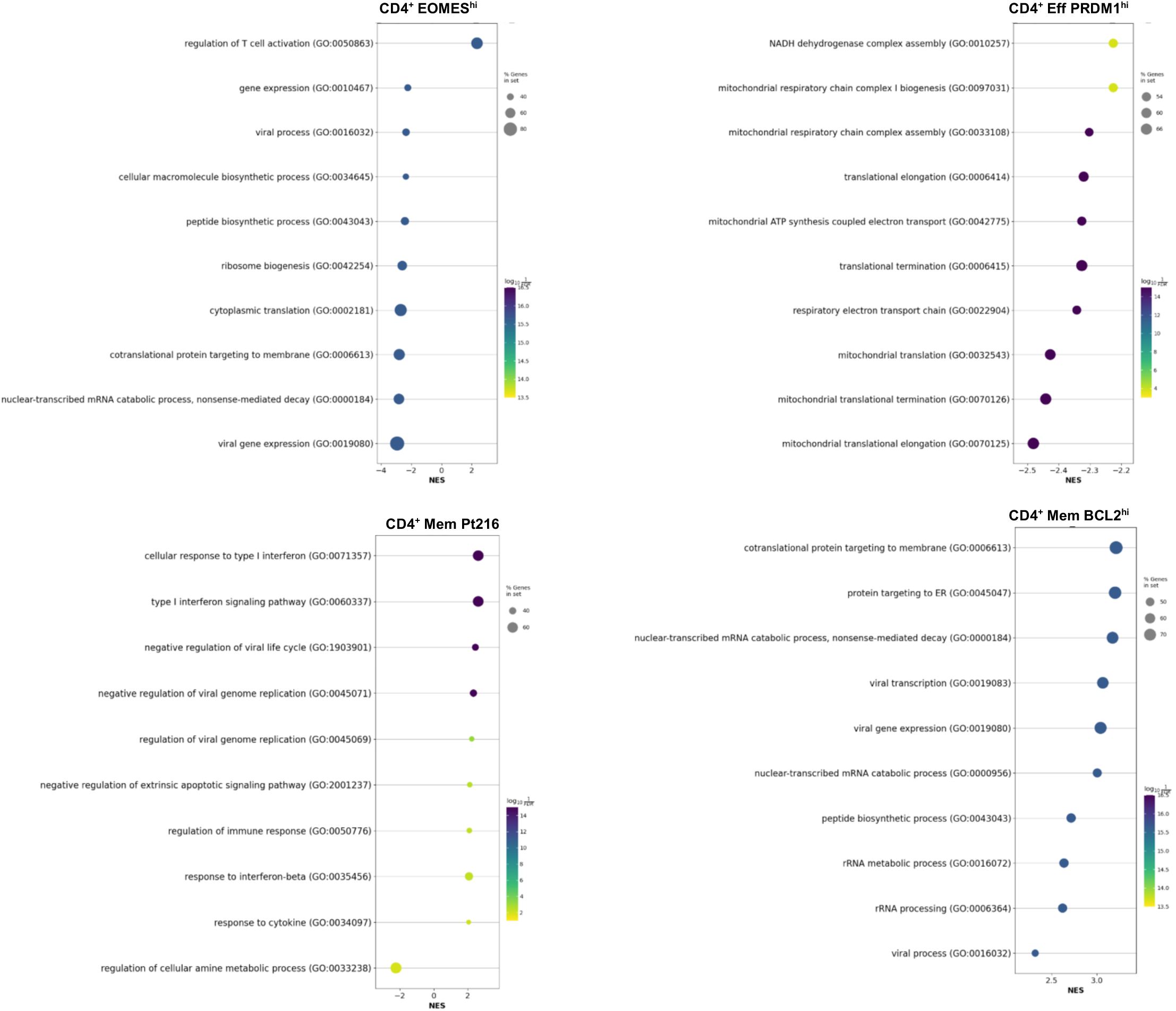
Gene ontology (GO) gene set analysis. GO gene sets associated with scRNA-seq defined CD4^+^ CAR T cell clusters. scRNA-seq = single cell RNA sequencing; CAR = chimeric antigen receptor

**Figure S13:**
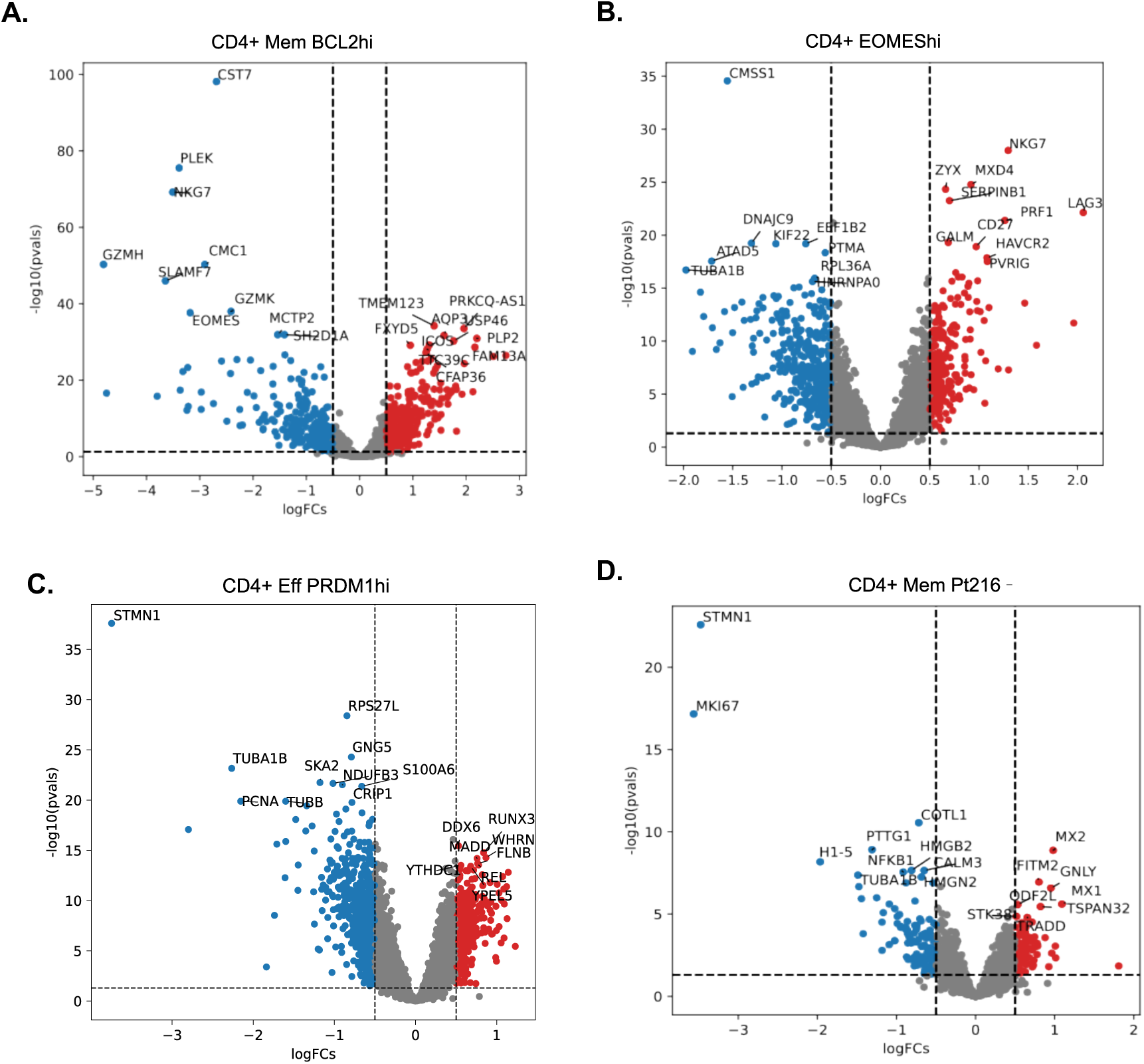
CD4+ Mem BCL demonstrates low GZMK expression. **(A-D)** Volcano plots depicting differential gene expression in the CL enriched CD4+ CAR T cell clusters including **(A)** CD4^+^ Mem BCL^hi^ **(B)** CD4^+^ EOMES^hi^ **(C)** CD4^+^ Eff PRDM1^hi^, and **(D)** CD4^+^ Mem Pt216

**Figure S14:**
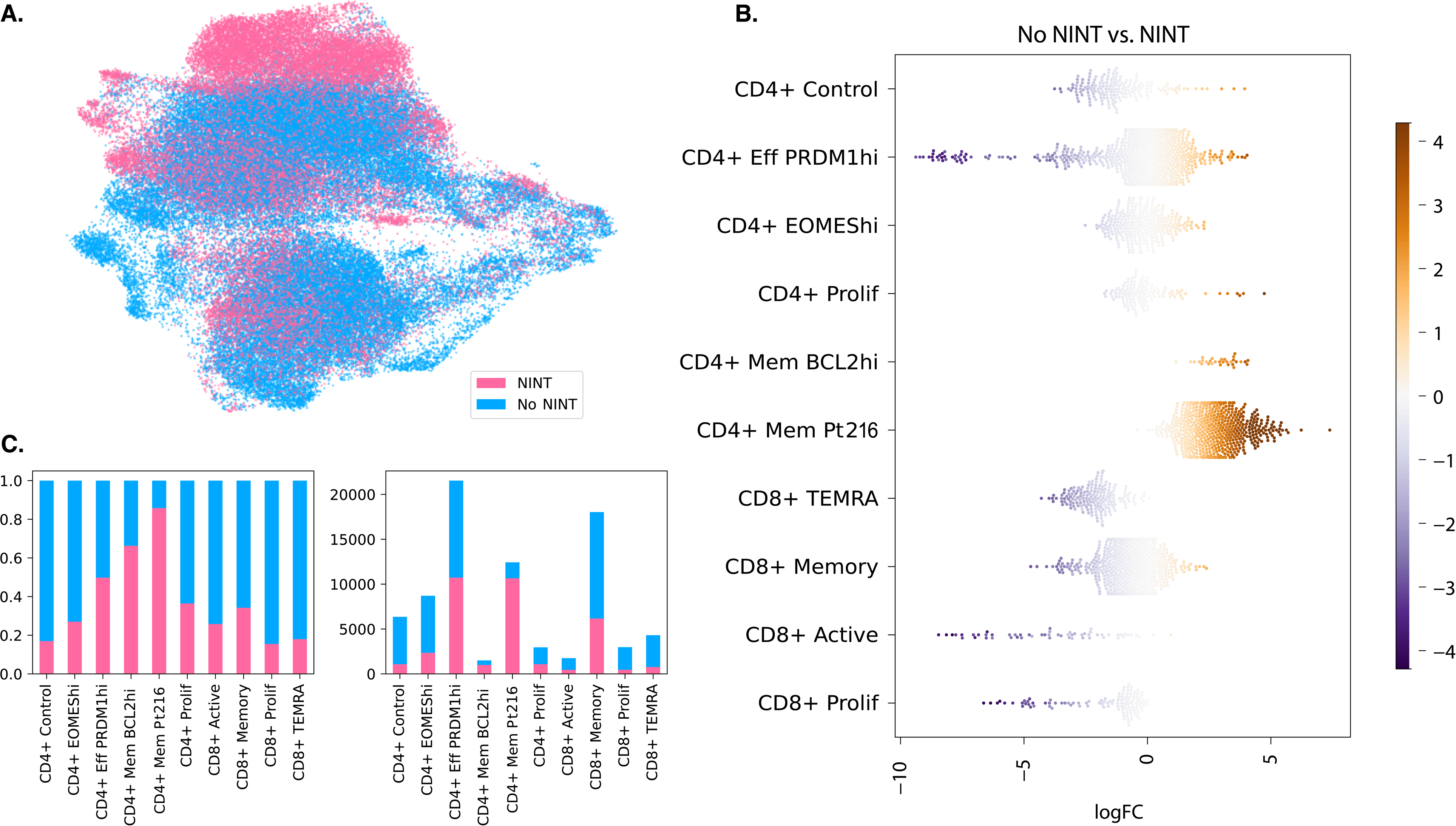
Identifying NINT enriched clusters. **(A)** UMAP projection color coded identifying CAR T cell from control (blue) vs NINT (pink) patients. **(B)** Abundance testing using Milo identifies clusters enriched in CAR T cell from control (blue) vs NINT (pink) patients. **(C)** Relative (left) and absolute (right) proportion of CAR T cells from control and NINT patients comprising each cluster. CAR = chimeric antigen receptor; NINT = non-ICANS neurotoxicity; UMAP = Uniform Manifold Approximation and Projection

**Figure S15:**
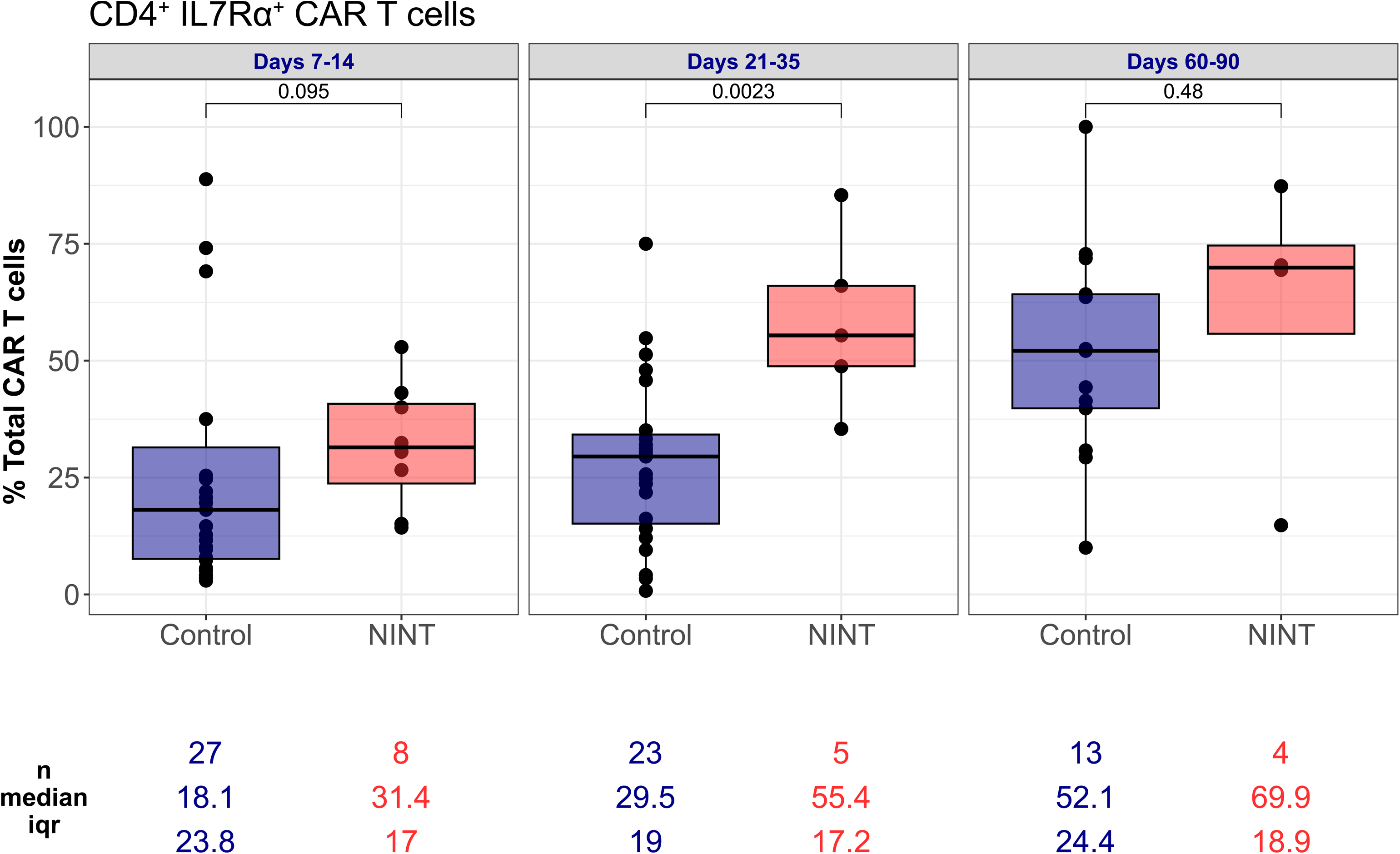
Patients with NINT are enriched in CD4+ IL-7Rα^+^ CAR T cells. CD4^+^ CD127/IL7rα^+^ CD4 CAR T cells measured by flow cytometry in control vs NINT patients over time

**Table S1:**
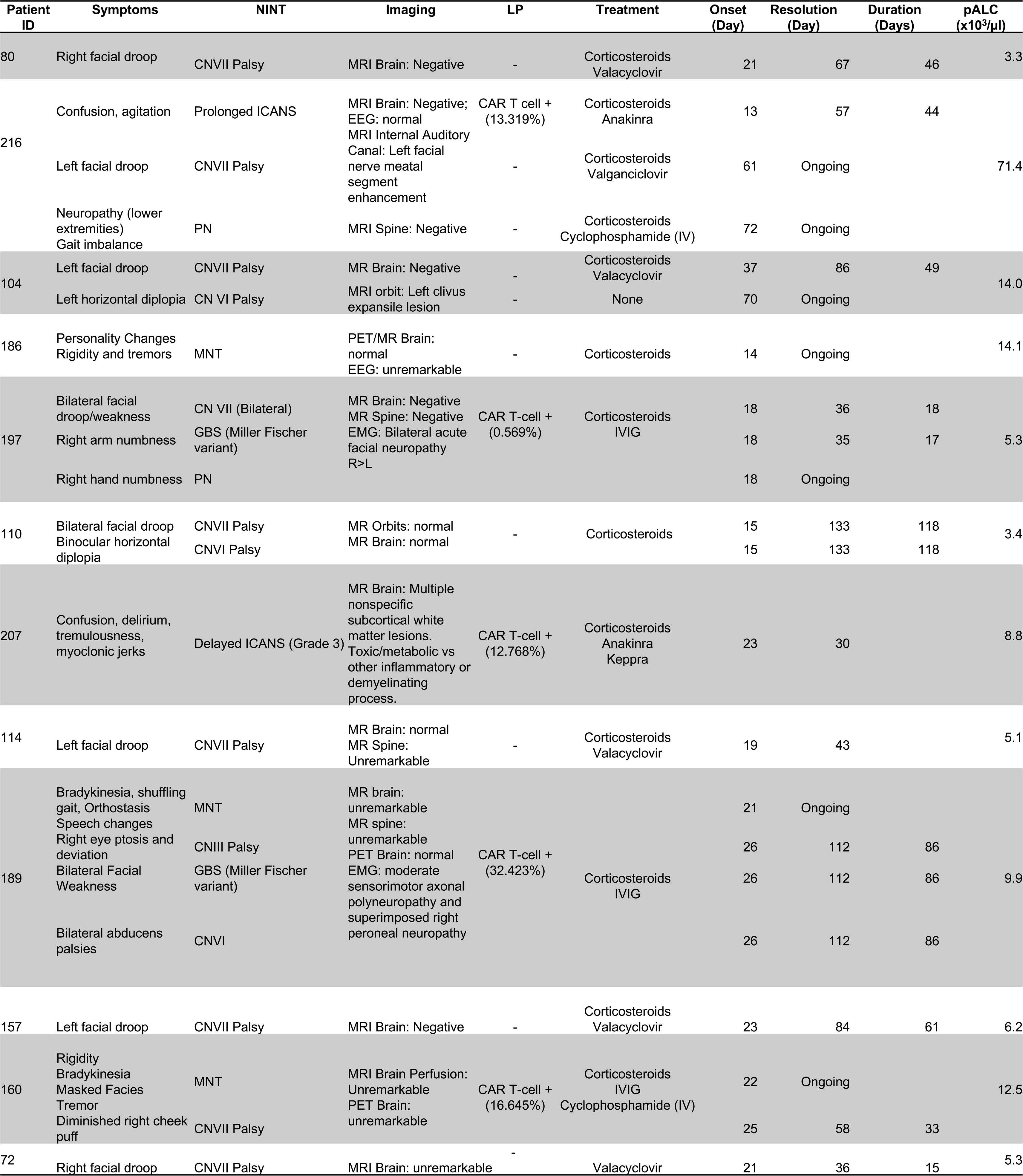
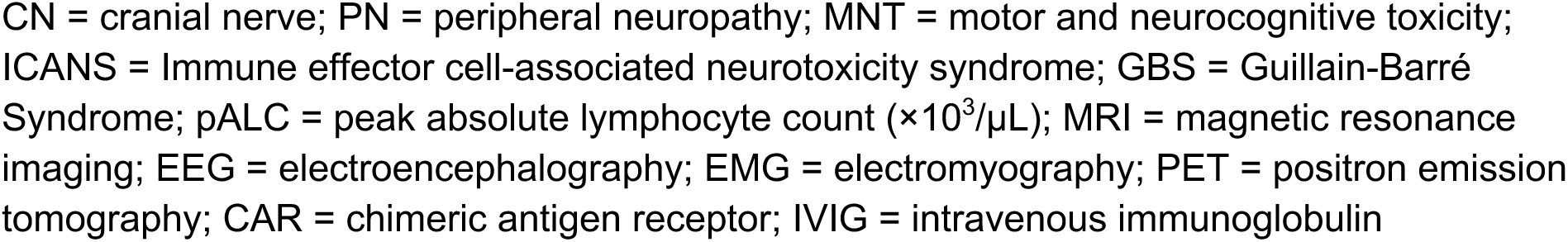
Clinical descriptions of patients diagnosed with NINTs.

**Table S2:**
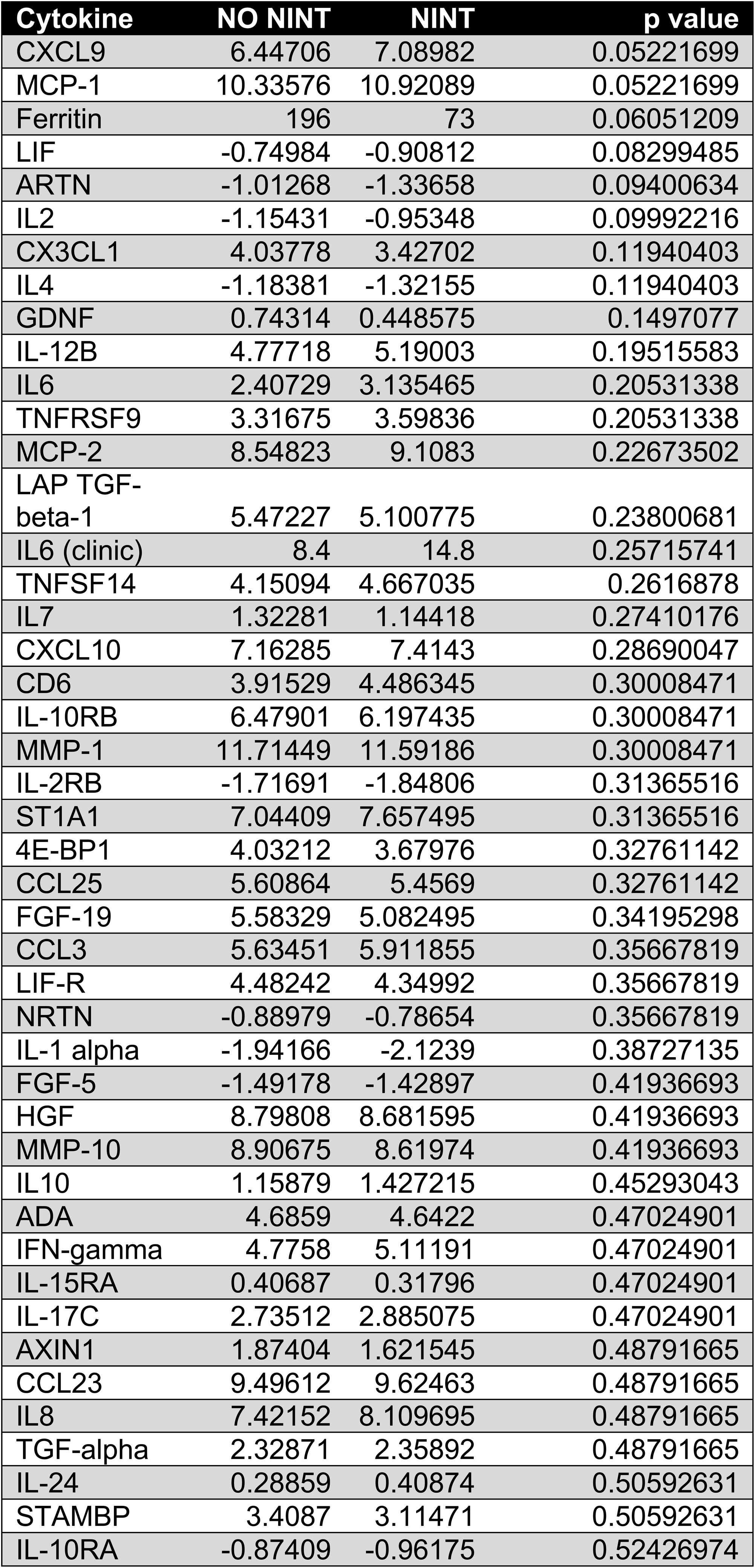
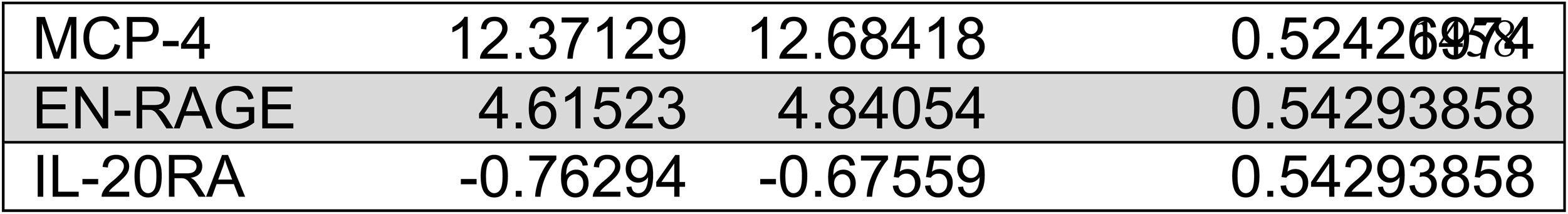
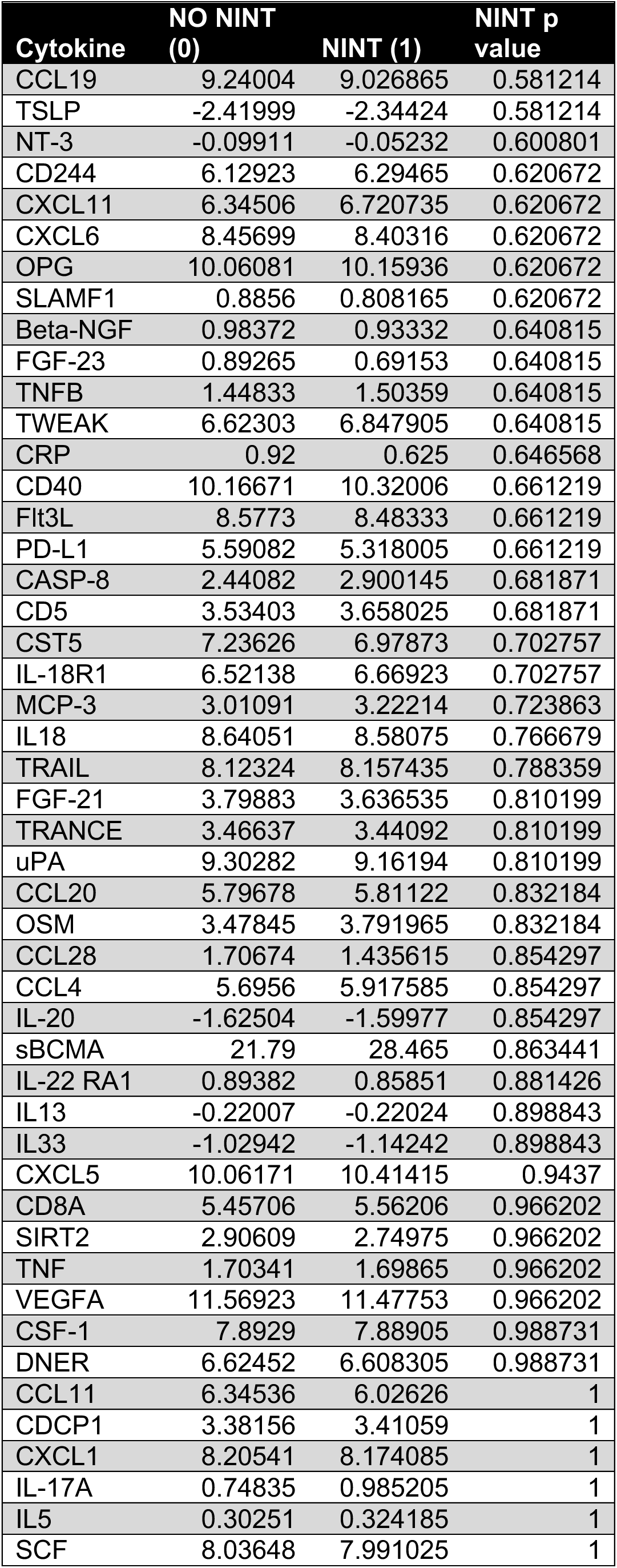
Olink Cytokine analysis comparing baseline cytokines in patients with NINT vs without NINT.

**Table S3:**
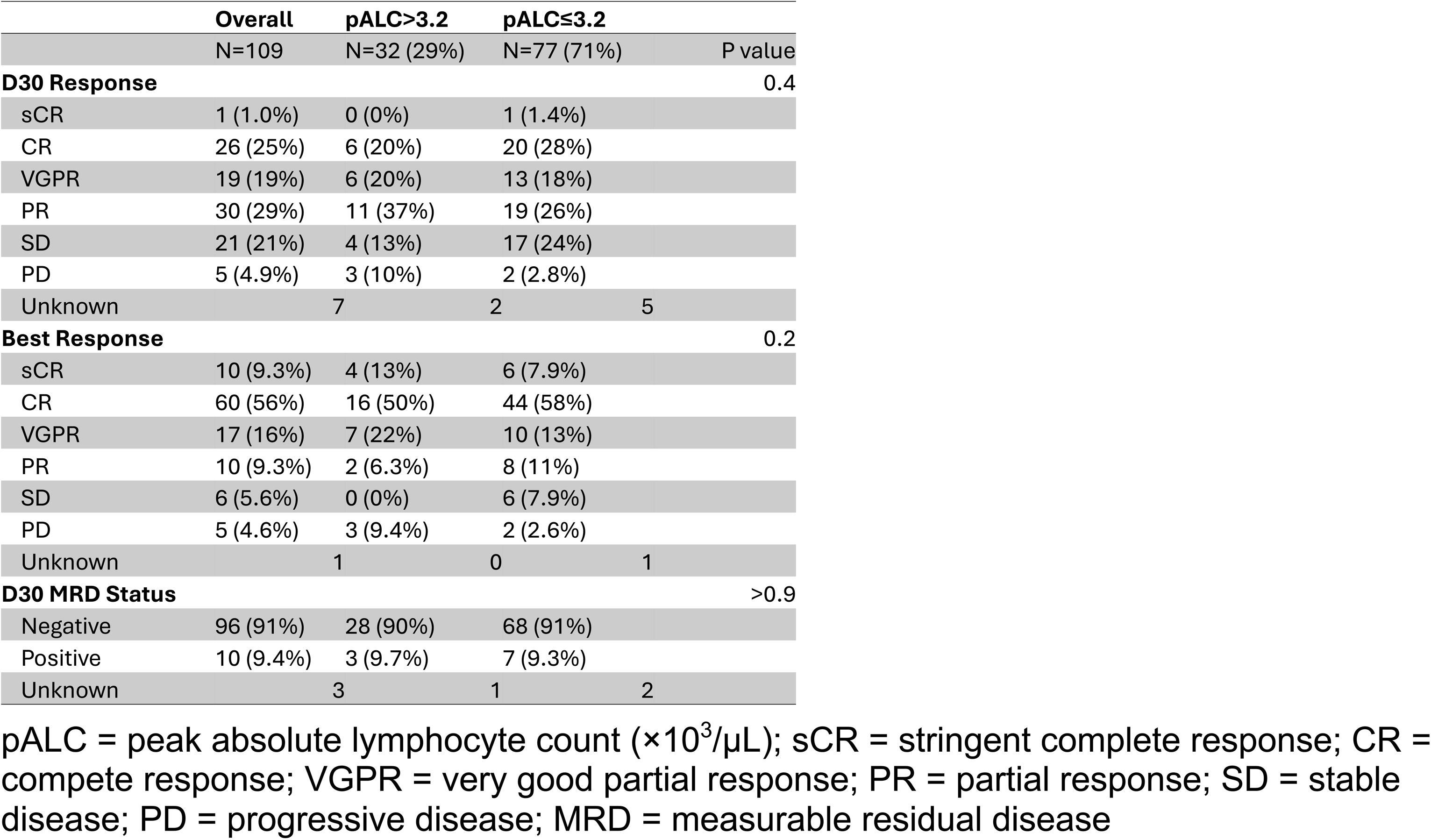
Patient Responses by pALC.

**Table S4.**
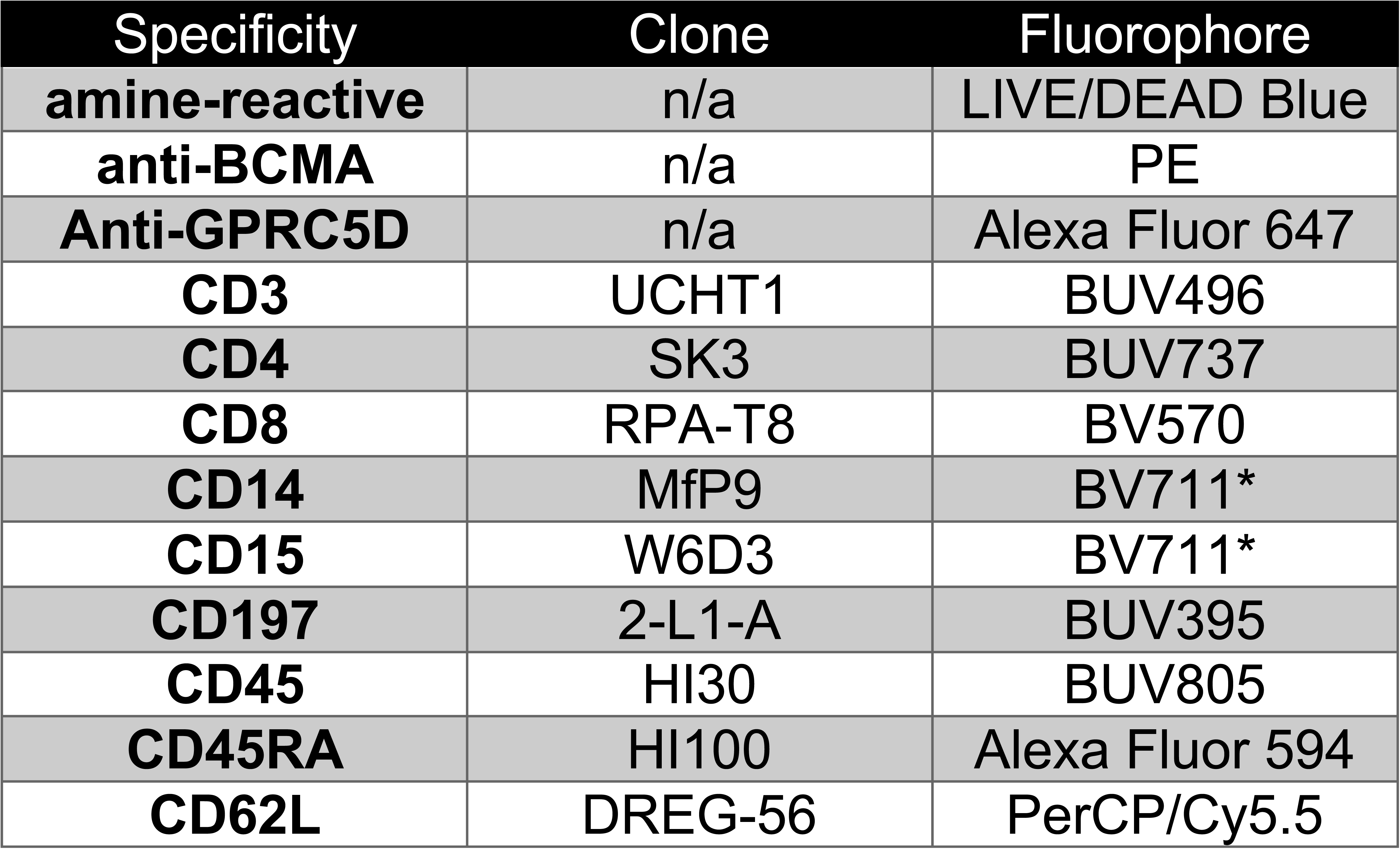
IDMS-022 CAR-T cell Screening Panel.

**Table S5.**
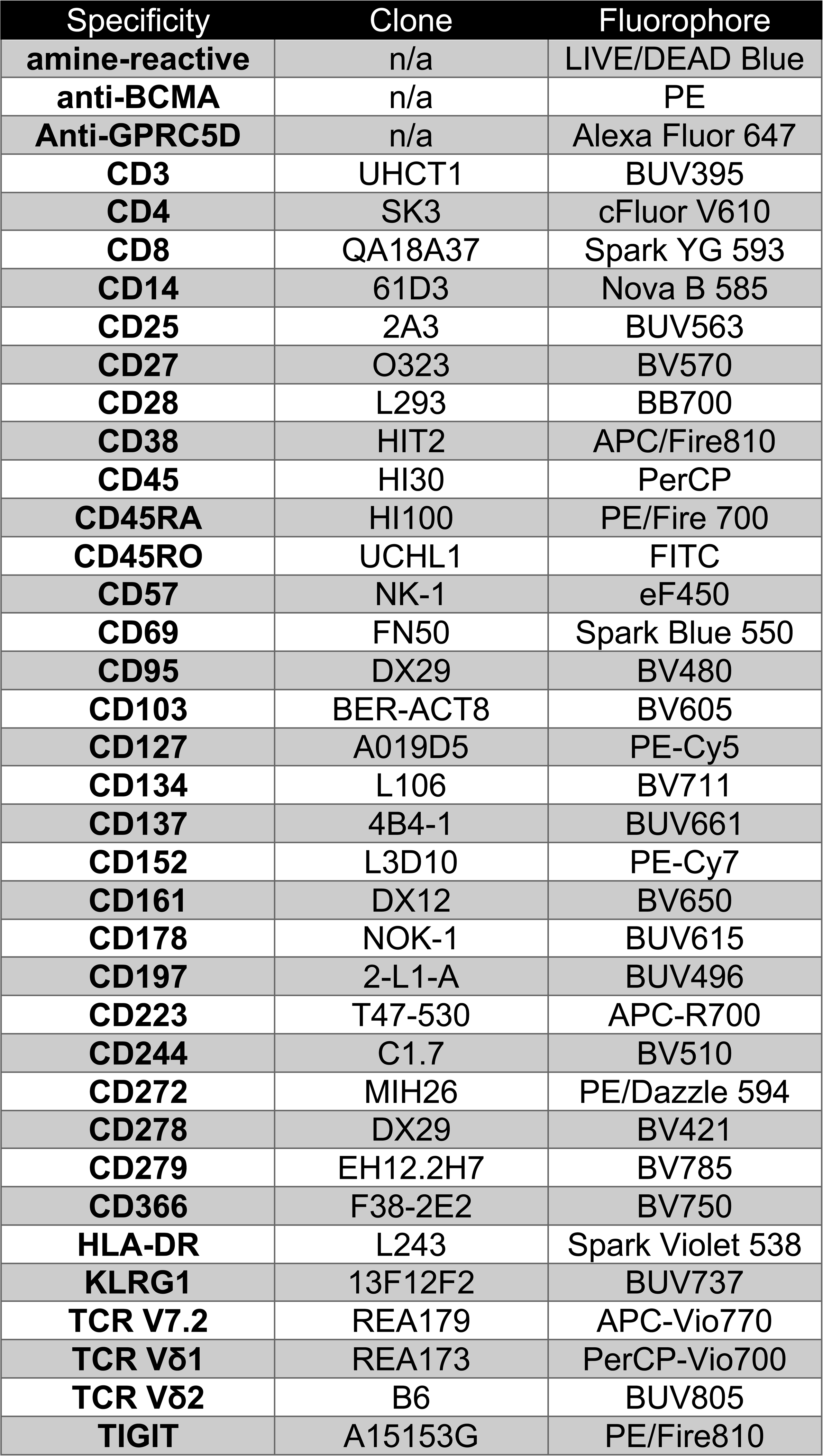
IDMS T ExAct Panel.

## Notes

### Competing Interest Statement

E.M.J. Grants: ASCO YIA and the IMS.
R.S.F. Grants: ASCO YIA and the IMS.
A.M.L. Grants: Novartis, BMS, Trillium Therapeutics, Pfizer, Janssen; Personal fees: Trillium
Therapeutics, Janssen; Patent (number US20150037346A1) with royalties paid.
N.K. Research funding: Amgen, Janssen, Epizyme, AbbVie; Consultancy: Clinical Care
Options, OncLive, and Intellisphere Remedy Health; Advisory board: Janssen,
MedImmune.
C.R.T. Research funding: Janssen, Takeda. Personal fees: Physician Educations Resource
and MJH Life Sciences; Advisory boards: Janssen and Sanofi.
H.H. Grants: Celgene, Takeda, and Janssen.
H.H. Advisory boards: Sanofi, BMS, and Janssen.
K.M. Funding: Sebia, Binding site, and Siemens.
U.A.S. Research support: Celgene/BMS and Janssen; Personal fees: MashUp MD, Janssen
Biotech, Sanofi, BMS, MJH Life Sciences, Intellisphere, Phillips Gilmore Oncology
Communications, i3 Health, and RedMedEd.
M.H. Research funding: Daiichi Sankyo, Cosette Pharmaceuticals, GlaxoSmithKline, Abbvie,
Beigene; Honoraria for consultancy/participated in advisory boards for Curio Science
LLC, Projects in Knowledge, Intellisphere LLC, Bristol Myers Squibb, Janssen, and
GlaxoSmithKline.
S.A.G. Personal fees and advisory board: Actinium, Celgene, BMS, Sanofi, Amgen, Pfizer,
GSK, Jazz, Janssen, Omeros, Takeda, and Kite.
G.L.S. Research funding: Janssen, Amgen, BMS, Beyond Spring; serves on the data safety
monitoring board (DSMB) for ArcellX; and receives research funding to the institution
from Janssen, Amgen, BMS, Beyond Spring, and GPCR.
H.J.L. Consultancy: Takeda, Genzyme, Janssen, Karyopharm, Pfizer, Celgene, Caelum
Biosciences; Research support: Takeda.
M.S. Consultancy: McKinsey & Company, Angiocrine Bioscience, Inc, and Omeros
Corporation; Research funding: Angiocrine Bioscience, Inc, Omeros Corporation, and
Amgen, Inc; Advisory boards: Kite, a Gilead company; Honoraria: i3 Health, Medscape,
and CancerNetwork for CME-related activity.
S.Z.U. Grants and personal fees: AbbVie, 404 Amgen, BMS, Celgene, GSK, Janssen, Merck,
Mundipharma, Oncopeptides, 405 Pharmacyclics, Sanofi, Seattle Genetics, SkylineDX,
and Takeda.
K.P. Intellectual Property Rights: NexImmune; Professional Services and Activities: Pfizer,
Inc.; RxCure LLC
S.M. Consultancy: Evicore, Optum, BioAscend, Janssen Oncology, BMS, AbbVie, HMP
Education, and Legend Biotech; Honoraria: OncLive, Physician Education Resource,
MJH Life Sciences, and Plexus Communications. Research funding: Janssen Oncology,
BMS, Allogene Therapeutics, Fate Therapeutics, Caribou Therapeutics, and Takeda
Oncology.

### Author Declarations

This study was approved by the Memorial Sloan Kettering Cancer Center Institutional Review Board. Patient samples were collected and analyzed in accordance with the Declaration of Helsinki.

