## Supplemental Materials for "Robust CD4^+^ CAR T cell Expansion Is Associated with Non-ICANS Neurotoxicities Following Ciltacabtagene Autoleucel"

**A. Supporting Information Figures**

*Figure S1. Overall Survival by pALC*

*Figure S2. Analysis Scheme for CAR-T cell Screening Panel*

*Figure S3. Analysis Scheme for IDMS T ExAct Panel.*

*Figure S4. Flow Cytometry Gating*

*Figure S5. Absolute CAR T cells by Flow*

*Figure S6. CD4/CD8 CAR T cell Subsets in Patients with NINT*

*Figure S7. Patients with CL are enriched in CAR T cells expressing memory markers*

*Figure S8. CD8^+^ TEMRA CAR T cells are enriched in patients without vs with CL*

*Figure S9. Inhibitory and exhaustion marker expression*

*Figure S10. Distribution of CAR T cells from individual patients included in CITE-seq analysis*

*Figure S11. Hallmark gene set analysis*

*Figure S12. Gene ontology gene set analysis*

*Figure S13. CD4^+^ Mem BCL demonstrates low GZMK expression*

*Figure S14. Figure S14: Identifying NINT enriched clusters*

*Figure S15: Patients with NINT are enriched in CD4+ IL-7Rα^+^ CAR T cells*

**B. Supporting Information Tables**

*Table S1: Clinical descriptions of patients diagnosed with NINTs*

*Table S2: Olink Baseline Cytokine Analysis*

*Table S3:* *Patient Responses by pALC*

*Table S4: IDMS-022 CAR-T cell Screening Panel.*

*Table S5: IDMS T ExAct Panel.*

**A.**

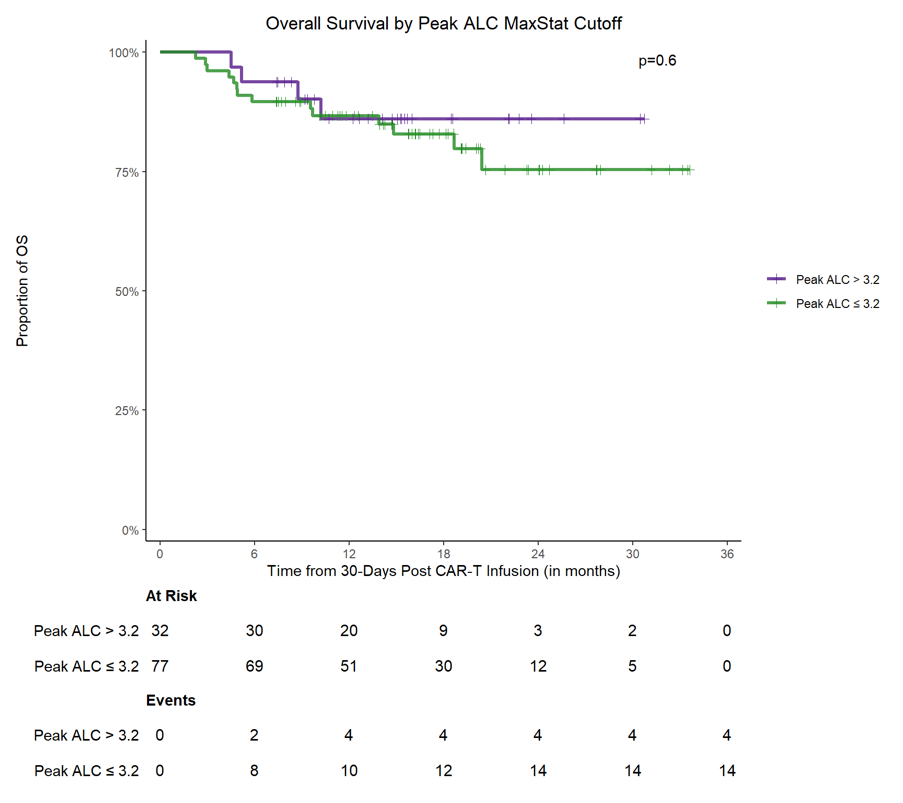

**B.**

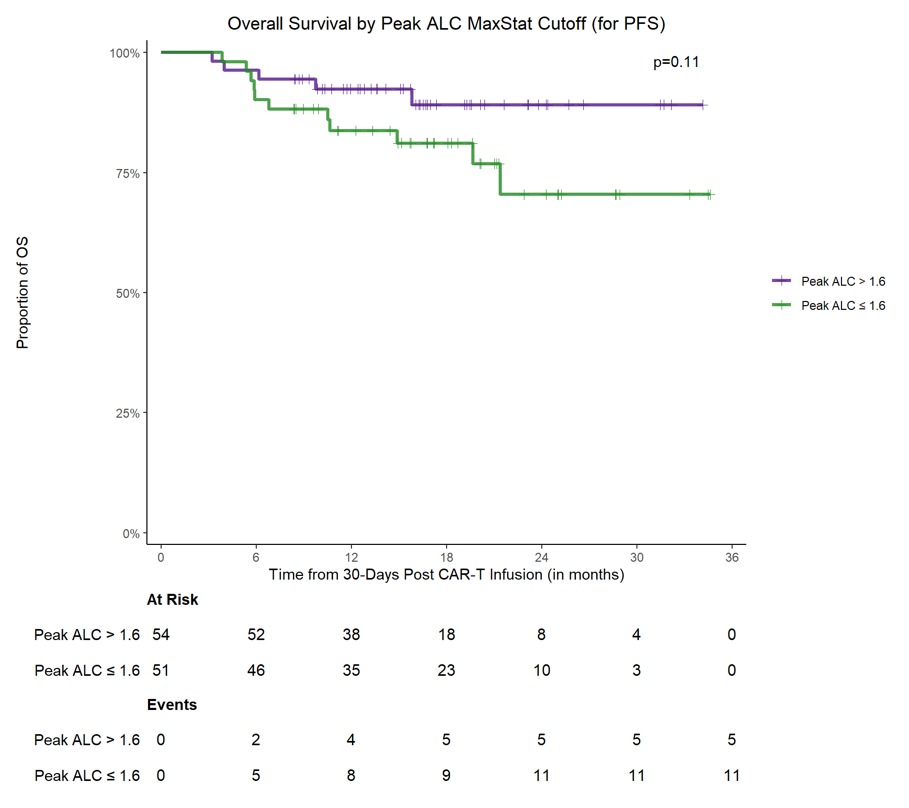

**Figure S1: OS by pALC. A,** OS of peak ALC groups separated by the identified NINT risk cutoff (3.2×10^3^/µL). **b,** OS of peak ALC groups separated by the identified optimal PFS cutoff (1.6×10^3^/µL). OS = overall survival; pALC = peak absolute lymphocyte count

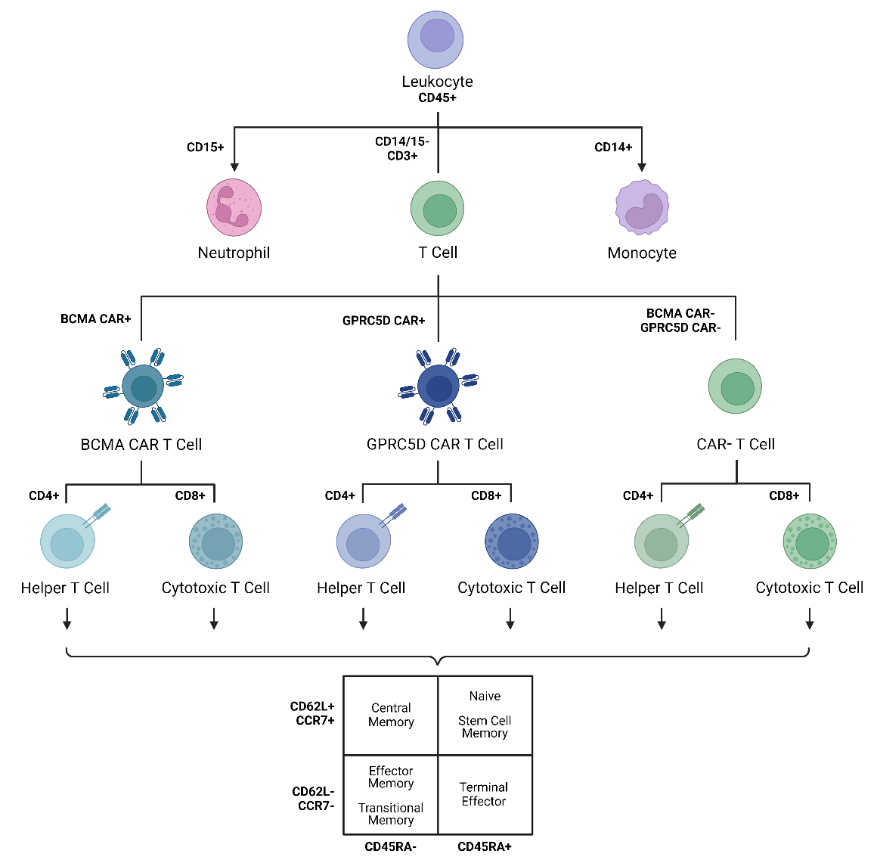

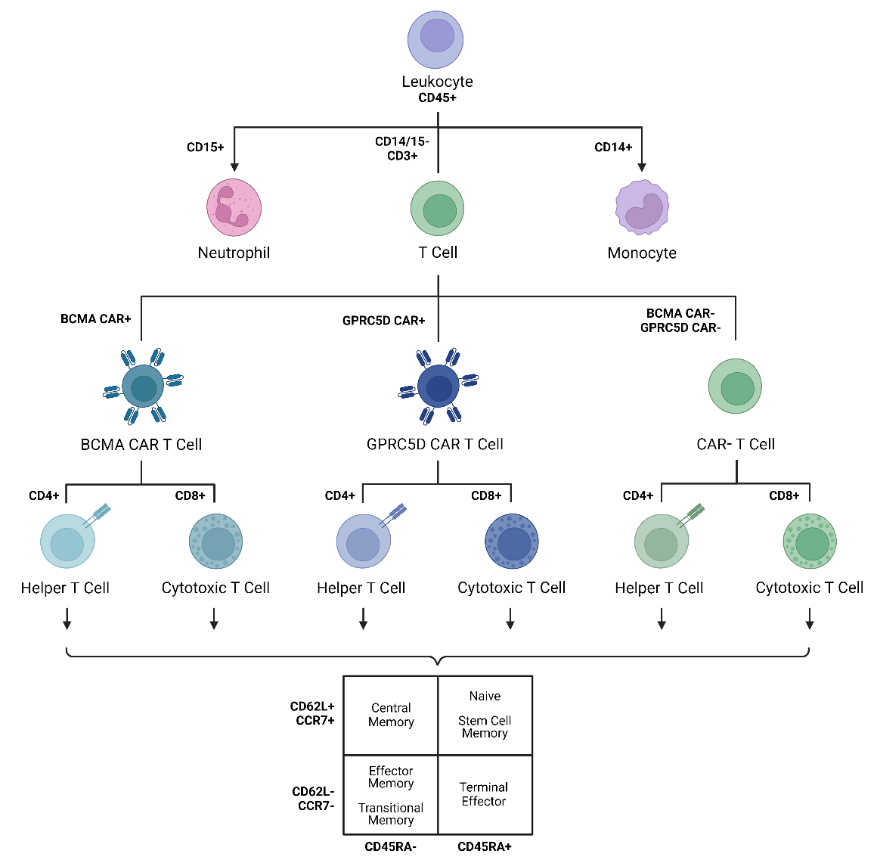

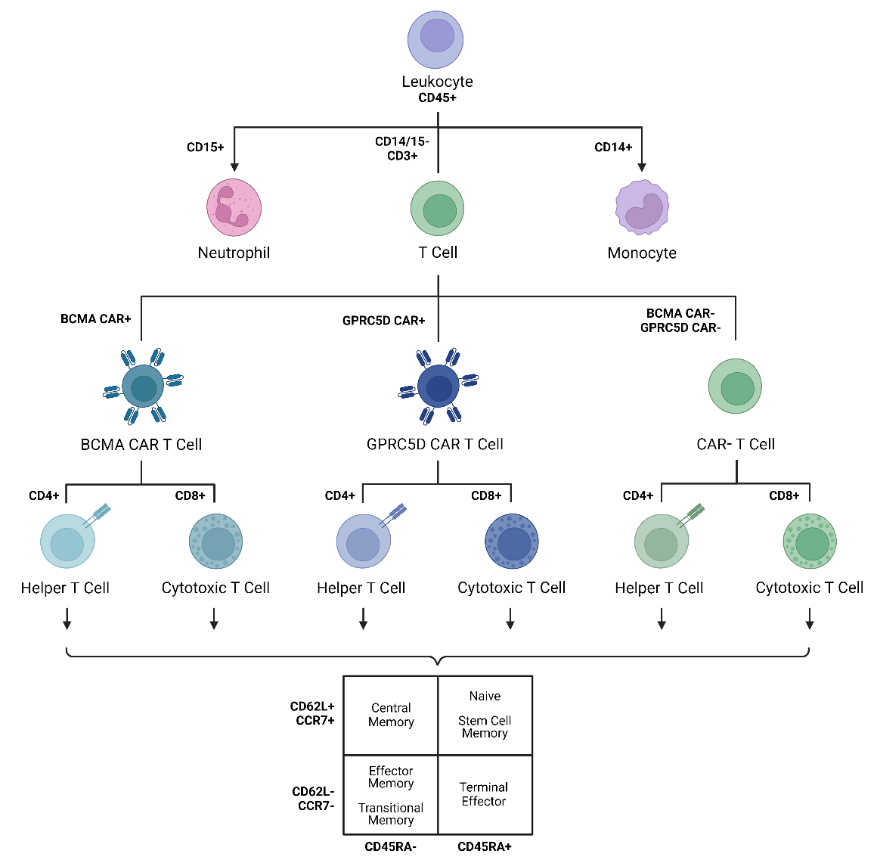

**Figure S2.** Analysis Scheme for CAR T cell Screening Panel.
BCMA = B cell maturation antigen; CAR = chimeric antigen receptor

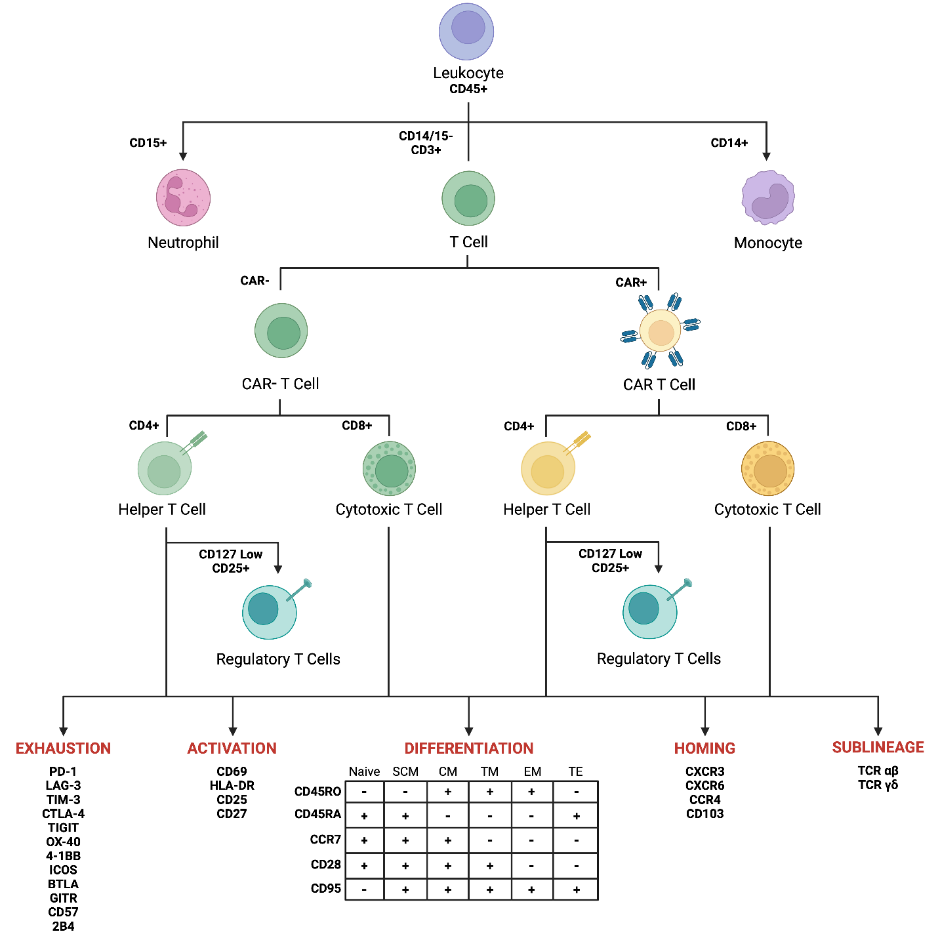

**Figure S3.** Analysis Scheme for IDMS T ExAct Panel.
CAR = chimeric antigen receptor

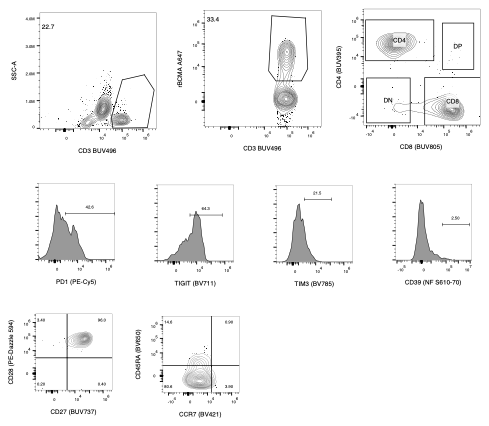

**A.**

**B.**

**C.**

**D.**

**E.**

**F.**

**G.**

**H.**

**I.**

**Figure S4: Flow cytometry gating strategy for patient peripheral blood CAR T cell immunophenotyping.** **a.** CD3 selection identified T cells **b.** recombinant BCMA isolated CAR^+^ T cells which were than categorized based on **c.** CD4 and CD8 expression. CAR T cells were further characterized by cells surface expression of **d,** PD-1, **e,** TIGIT, **f,** TIM-3, **g,** CD39, **h,** CD27 and CD28, **i,** CD45RA and CCR7.

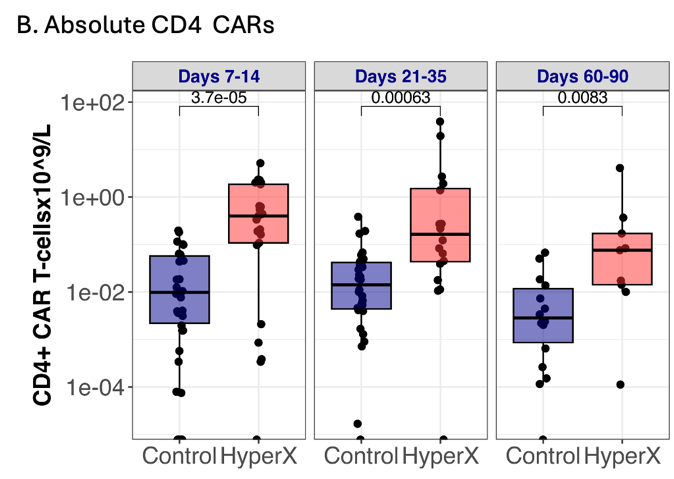

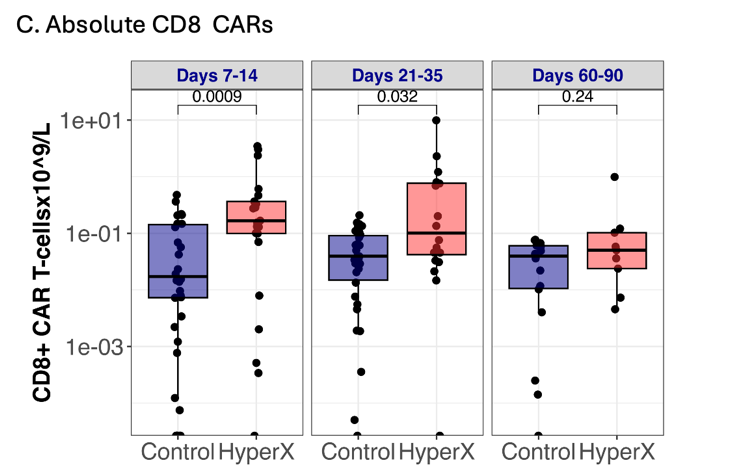

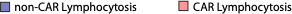

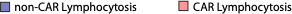

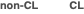

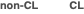

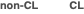

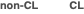

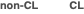

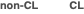

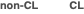

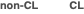

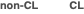

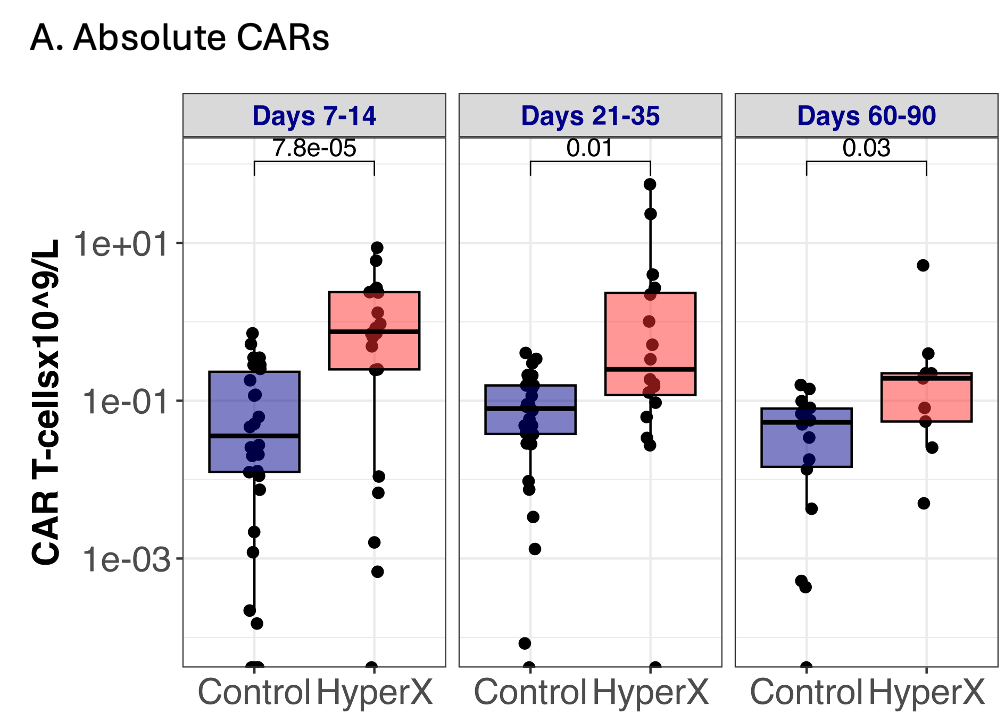

**Figure S5: Patients with CL Exhibit Higher Peripheral Absolute CAR T cells. a,** Absolute total CAR T cells, b, CD4^+^ CAR T cells, and c, CD8^+^ CAR T cells in patients with CAR lymphocytosis (red) vs without CAR lymphocytosis (blue) during three time windows; Days 7-14 (n_non-CL_=30, n_CL_=21), Days 21-35 (n_non-CL_=30, n_CL_=17), Days 60-90 (n_non-CL_=14, n_CL_=9). P values were calculated using Mann-Whitney U test and depicted above boxes. CAR = chimeric antigen receptor

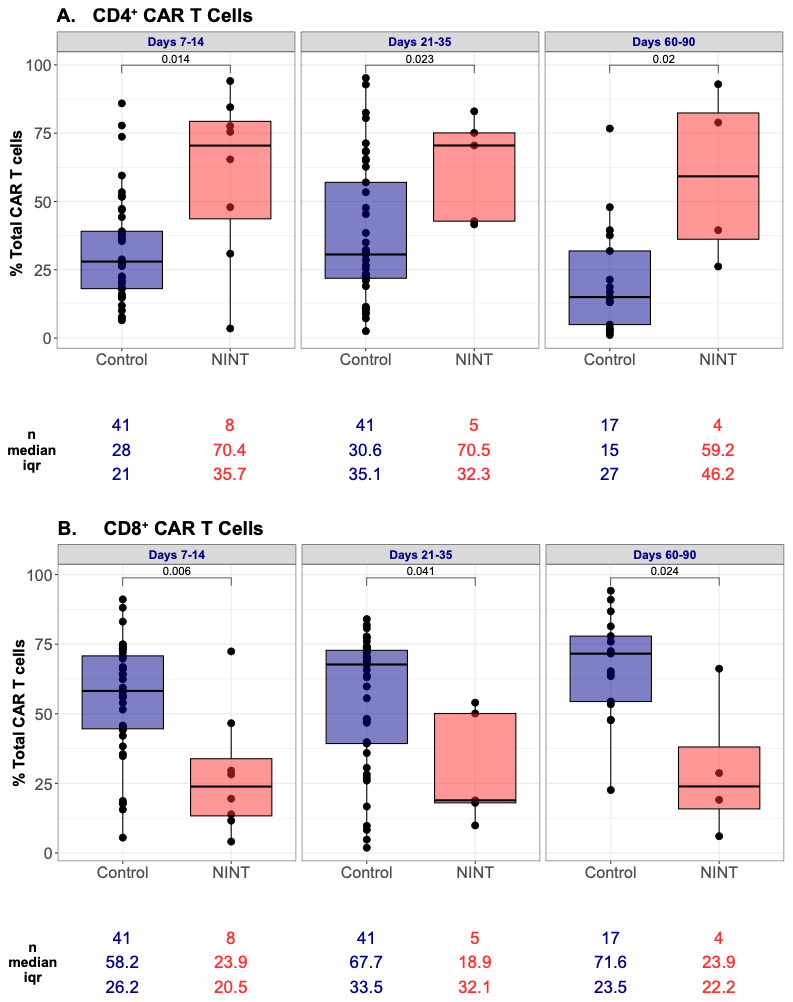

**Figure S6: Patients with NINT exhibit bias towards CD4^+^ CAR T cells. a,** Proportion of peripheral blood CD4^+^ CAR T cells and **b,** CD8^+^ CAR T cells in patients with NINT (red) vs without NINT (Control; blue) measured by flow cytometry during three time windows; Days 7-14 (n_control_=41, n_nint_=8), Days 21-35 (n_control_=41, n_nint_=5), Days 60-90 (n_control_=17, n_nint_=4). P values were calculated using Mann-Whitney U test and depicted above boxes. CAR = chimeric antigen receptor

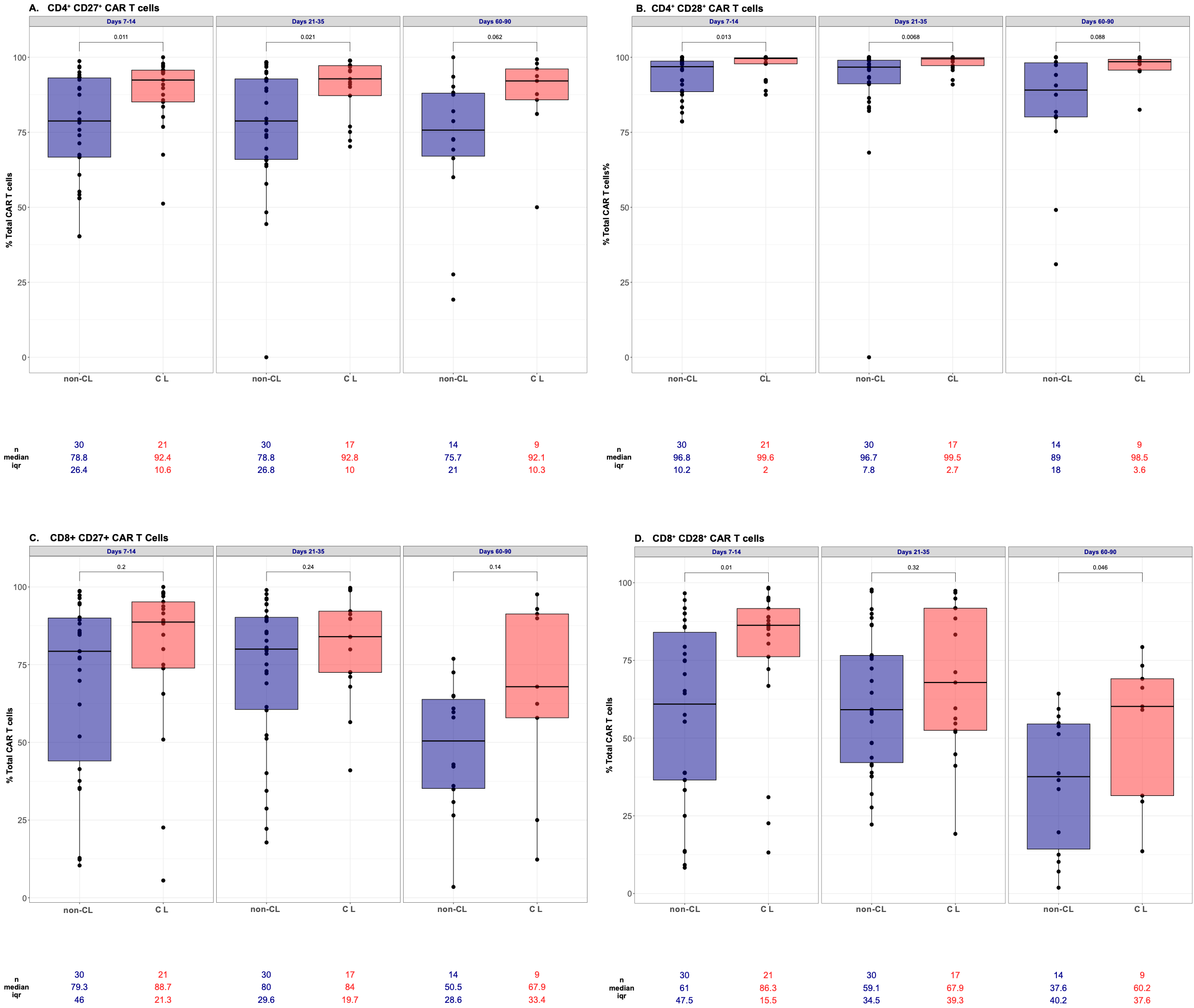

**Figure S7: Patients with CL are enriched in CAR T cells expressing memory markers CD27 and CD28. a,** Proportion of peripheral blood CD4^+^ CD27^+^ CAR T cells, **b,** CD4^+^ CD28^+^ CAR T cells, **c**, CD8^+^ CD27^+^ CAR T cells, and **d,** CD8^+^ CD28^+^ CAR T cells in patients with CL (red) vs without CL (blue) measured by flow cytometry during three time windows; Days 7-14 (n_non-CL_=30, n_CL_=21), Days 21-35 (n_non-CL_=30, n_CL_=17), Days 60-90 (n_non-CL_=14, n_CL_=9). P values were calculated using Mann-Whitney U test and depicted above boxes. CAR = chimeric antigen receptor; CL = CAR lymphocytosis

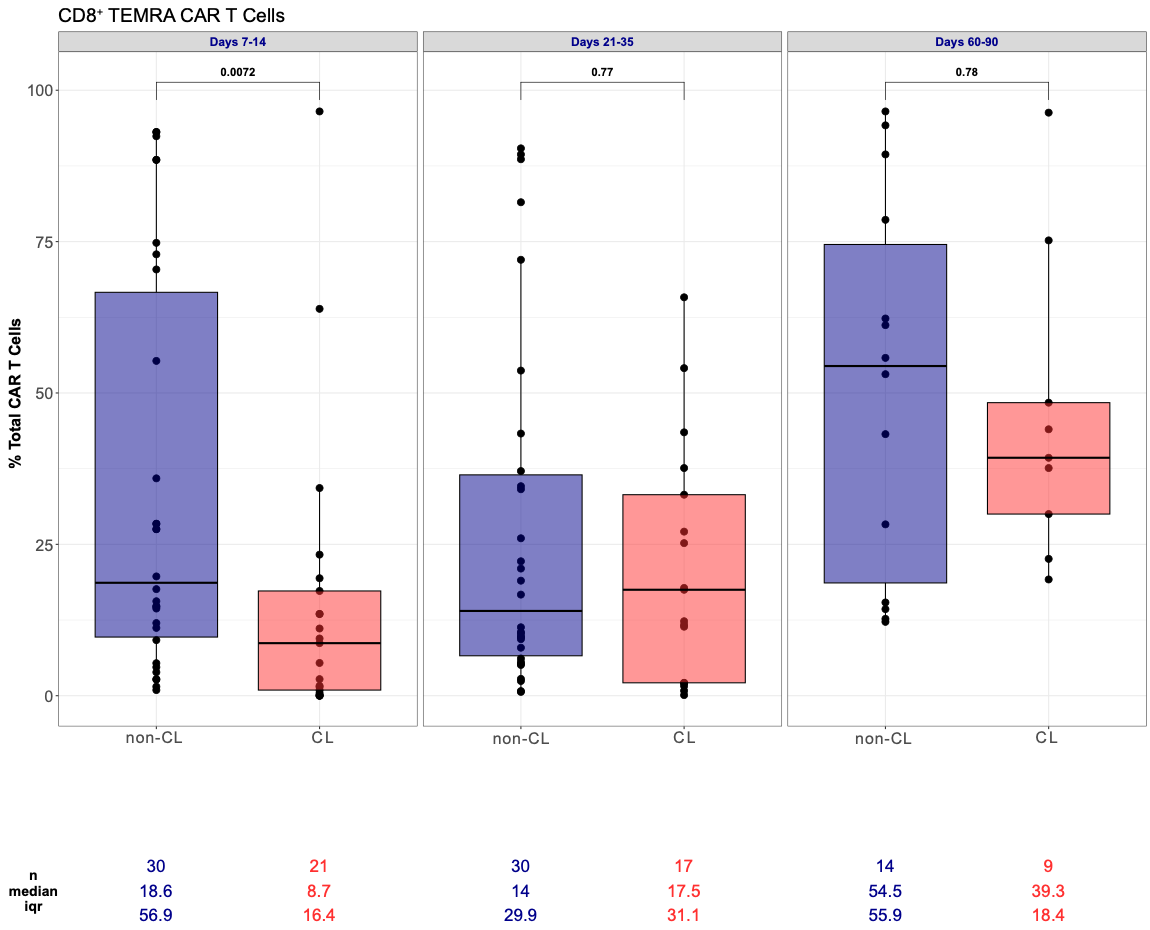

**Figure S8: CD8^+^ TEMRA CAR T cells are enriched in patients without vs with CL. A,** Peripheral blood CD8^+^ TEMRA CAR T cells in patients with CL (red) vs without CL (blue) measured by flow cytometry as a percentage of total CAR T cells during three time windows; Days 7-14 (n_non-CL_=30, n_CL_=21), Days 21-35 (n_non-CL_=30, n_CL_=17), Days 60-90 (n_non-CL_=14, n_CL_=9). P values were calculated using Mann-Whitney U test and depicted above boxes. CAR = chimeric antigen receptor; CL = CAR lymphocytosis

**Figure S9: Inhibitory and exhaustion marker expression was similar amongst patients with and without CL a,** Proportion of peripheral blood CD4+ PD-1+ CAR T cells, **b,** CD8+ PD-1+ CAR T cells, **c**, CD4+ TIGIT+ CAR T cells, and **d,** CD8+ TIGIT+ CAR T cells, **e,** CD4+ CD39+ CAR T cells, **f**, CD8+ CD39+ CAR T cells, **g,** CD4+ TIM-3+ CAR T cells, and **h,** CD8+ TIM-3+ CAR T cells in patients with CL (red) vs without CL (blue) measured by flow cytometry during three time windows; Days 7-14 (n_non-CL_=30, n_CL_=21), Days 21-35 (n_non-CL_=30, n_CL_=17), Days 60-90 (n_non-CL_=14, n_CL_=9). For CD39+ CAR T cells: Days 7-14 (n_non-CL_=20, n_CL_=16), Days 21-35 (n_non-CL_=19, n_CL_=10), Days 60-90 (n_non-CL_=10, n_CL_=7). P values were calculated using Mann-Whitney U test and depicted above boxes. CAR = chimeric antigen receptor; CL = CAR lymphocytosis

**(cont) Figure S9: Inhibitory and exhaustion marker expression was similar amongst patients with and without CL a,** Proportion of peripheral blood CD4+ PD-1+ CAR T cells, **b,** CD8+ PD-1+ CAR T cells, **c**, CD4+ TIGIT+ CAR T cells, and **d,** CD8+ TIGIT+ CAR T cells, **e,** CD4+ CD39+ CAR T cells, **f**, CD8+ CD39+ CAR T cells, **g,** CD4+ TIM-3+ CAR T cells, and **h,** CD8+ TIM-3+ CAR T cells in patients with CL (red) vs without CL (blue) measured by flow cytometry during three time windows; Days 7-14 (n_non-CL_=30, n_CL_=21), Days 21-35 (n_non-CL_=30, n_CL_=17), Days 60-90 (n_non-CL_=14, n_CL_=9). For CD39+ CAR T cells: Days 7-14 (n_non-CL_=20, n_CL_=16), Days 21-35 (n_non-CL_=19, n_CL_=10), Days 60-90 (n_non-CL_=10, n_CL_=7). P values were calculated using Mann-Whitney U test and depicted above boxes. CAR = chimeric antigen receptor; CL = CAR lymphocytosis

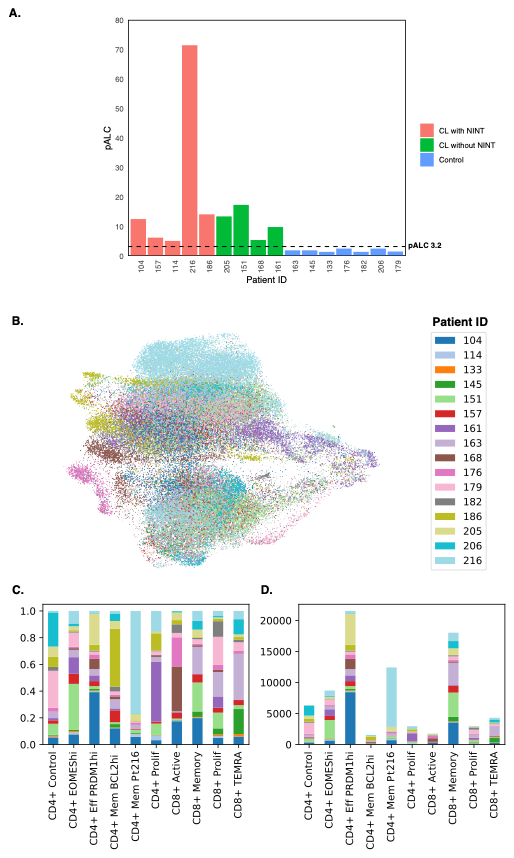

**CD4^+^ memory Pt216**

**Figure S10: Distribution of CAR T cells from individual patients included in CITE-seq analysis. a,** Individual patients arranged on x axis with pALC depicted on y axis, patients are grouped according to history of CL with NINT (red), CL without NINT (green), or neither CL nor NINT (control; blue). **b,** UMAP projection color coded identifying CAR T cells from individual patients (legend on right). CD4^+^ memory Pt216 cluster is highlighted demonstrating near complete composition from patient 216. **c,** Relative and **d,** absolute measurement of CAR T cells from individual patients in each cluster. CAR = chimeric antigen receptor; pALC = peak absolute lymphocyte count; NINT = non-ICANS neurotoxicity; UMAP = Uniform Manifold Approximation and Projection

CD4^+^ EOMES^hi^

CD4^+^ Eff PRDM1^hi^

CD4^+^ Control

CD4^+^ Mem Pt216

CD4^+^ EOMES^hi^

CD4^+^ Eff PRDM1^hi^

CD4^+^ Mem Pt216

CD4^+^ Control

CD4^+^ Mem BCL2^hi^

CD4^+^ EOMES^hi^

CD4^+^ Eff PRDM1^hi^

CD4^+^ Mem Pt216

CD4^+^ Control

CD4^+^ Mem BCL2^hi^

CD4^+^ EOMES^hi^

CD4^+^ Eff PRDM1^hi^

CD4^+^ Mem Pt216

CD4^+^ Control

CD4^+^ Mem BCL2^hi^

CD4^+^ EOMES^hi^

CD4^+^ Eff PRDM1^hi^

CD4^+^ Mem Pt216

CD4^+^ Control

CD4^+^ Mem BCL2^hi^

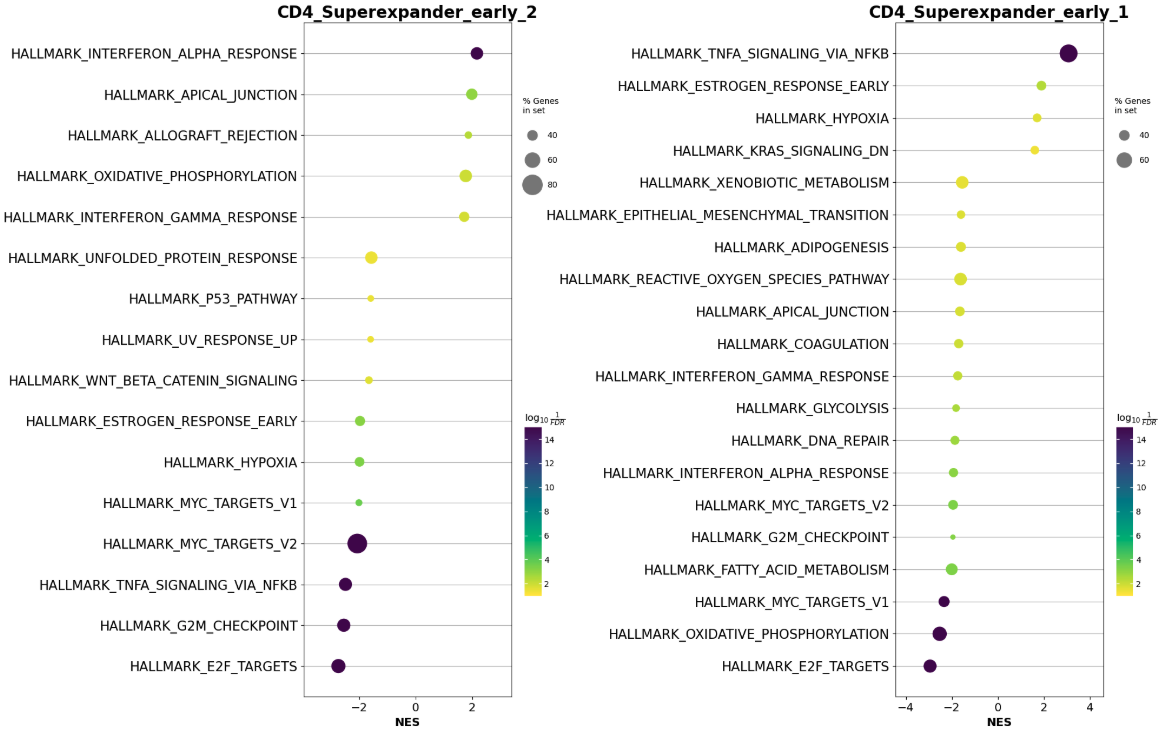

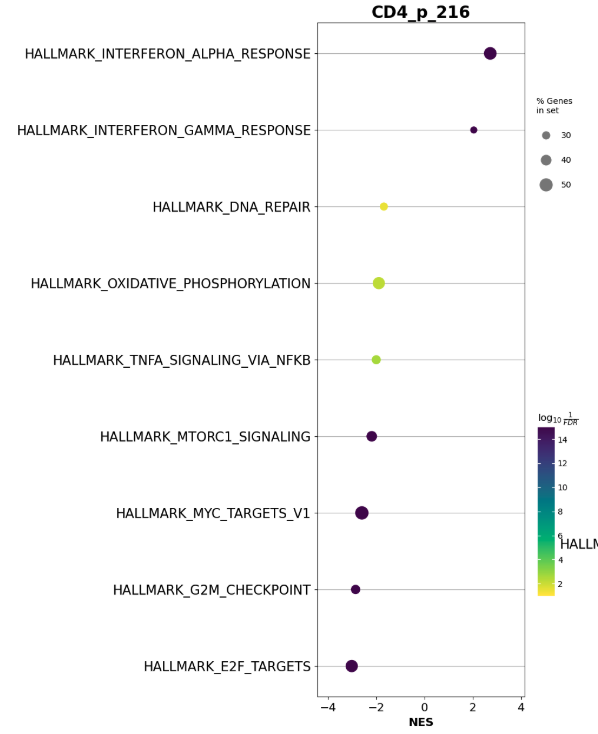

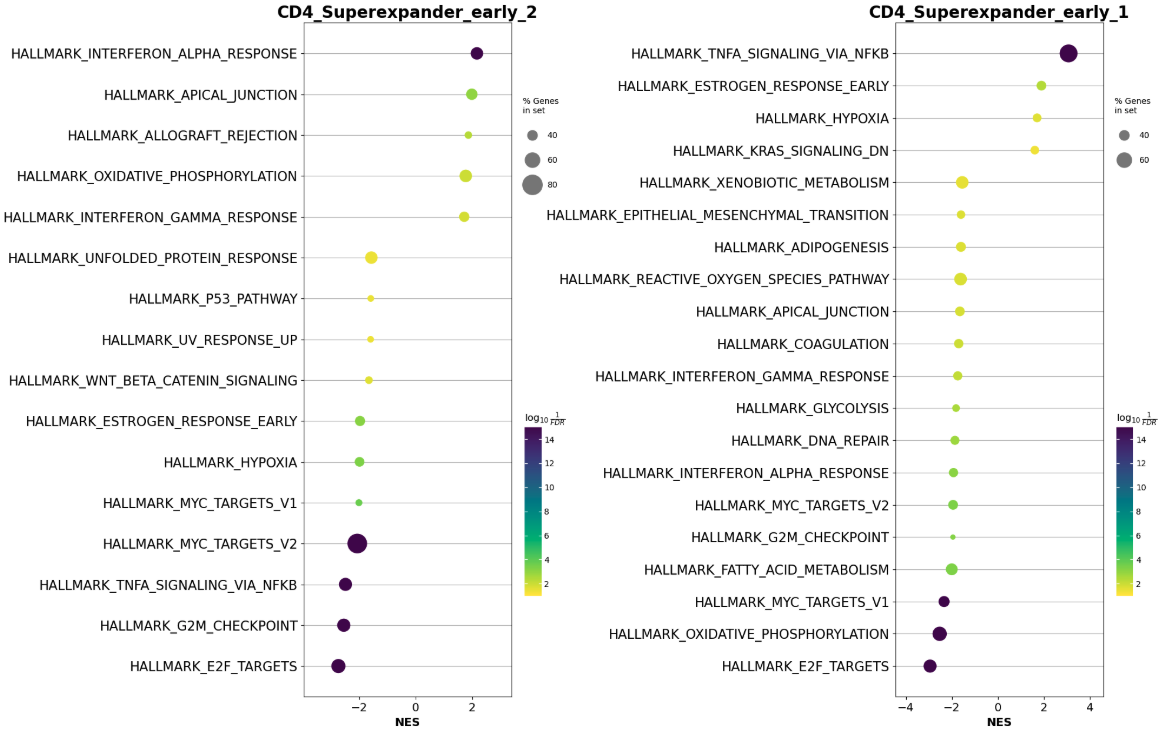

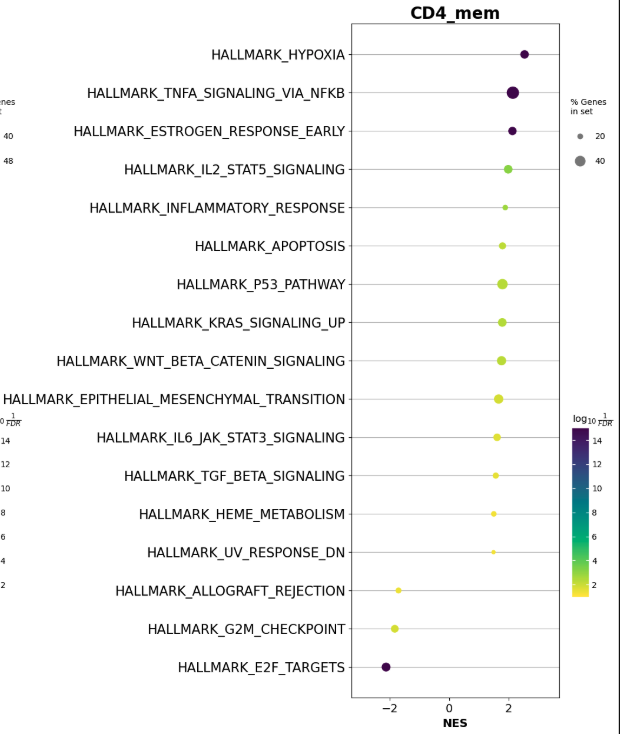

**CD4^+^ Mem Pt216**

**CD4^+^ Mem BCL2^hi^**

**CD4^+^ EOMES^hi^**

**CD4^+^ Eff PRDM1^hi^**

**Figure S11: Hallmark gene set analysis**. Hallmark gene sets associated with scRNA-seq defined CD4^+^ CAR T cell clusters. scRNA-seq = single cell RNA sequencing; CAR = chimeric antigen receptor

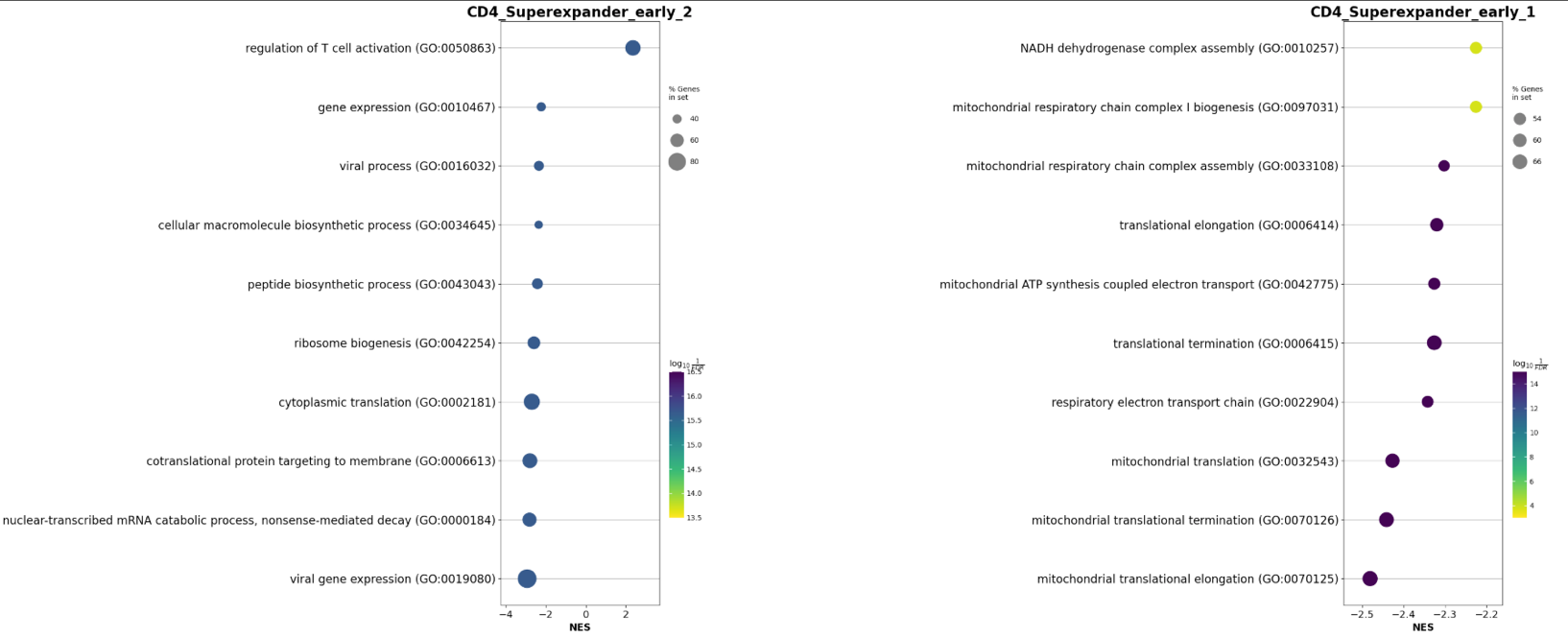

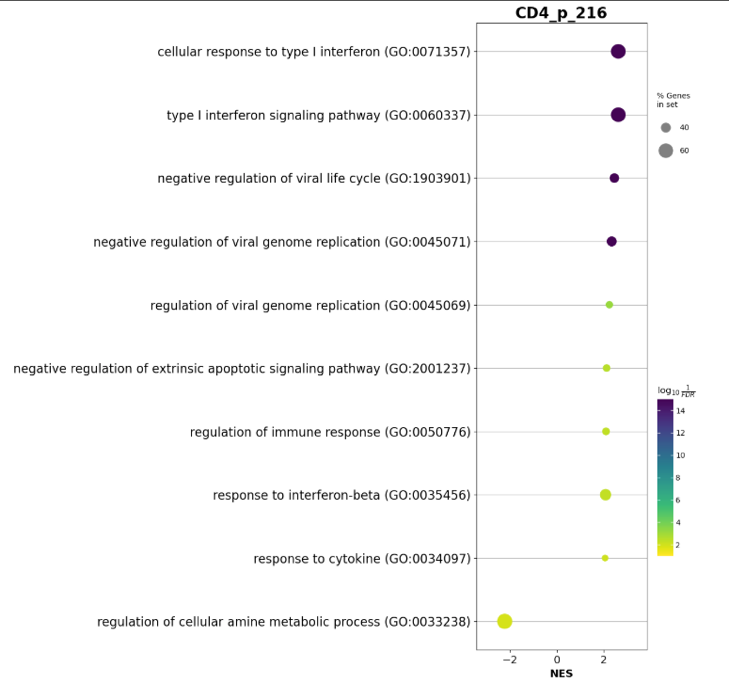

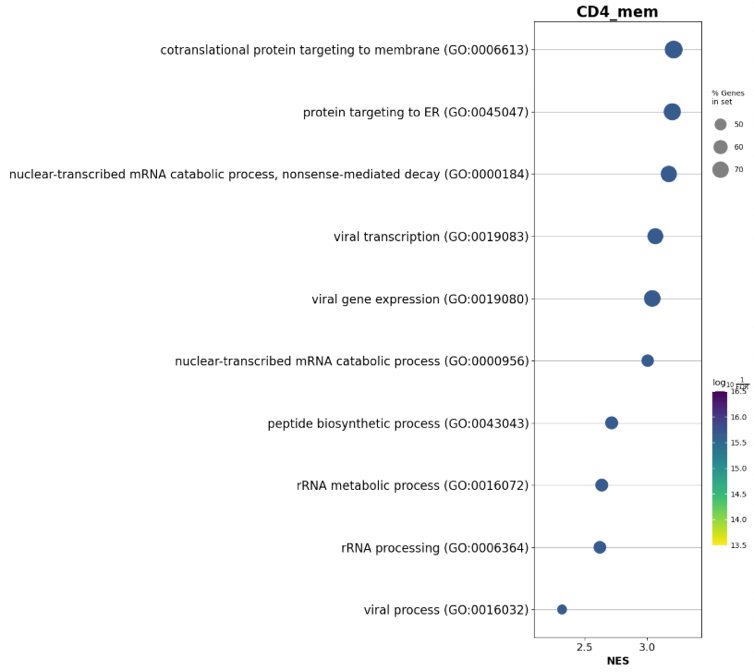

**CD4^+^ EOMES^hi^**

**CD4^+^ Eff PRDM1^hi^**

**CD4^+^ Mem Pt216**

**CD4^+^ Mem BCL2^hi^**

**Figure S12: Gene ontology (GO) gene set analysis**. GO gene sets associated with scRNA-seq defined CD4^+^ CAR T cell clusters. scRNA-seq = single cell RNA sequencing; CAR = chimeric antigen receptor

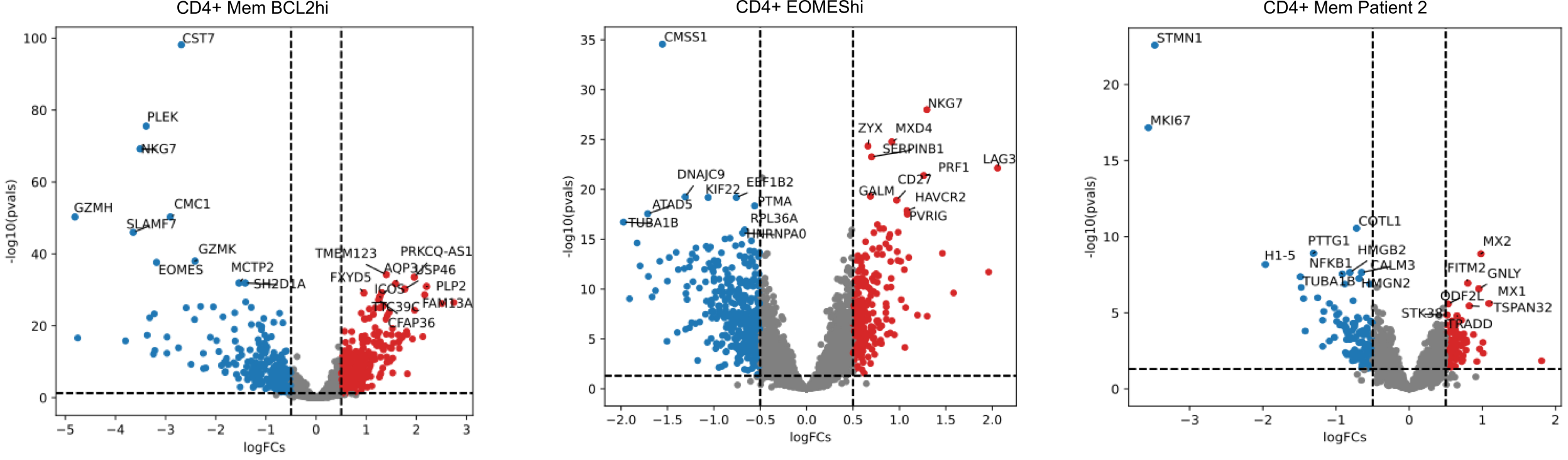

**B.**

**A.**

**C.**

**D.**

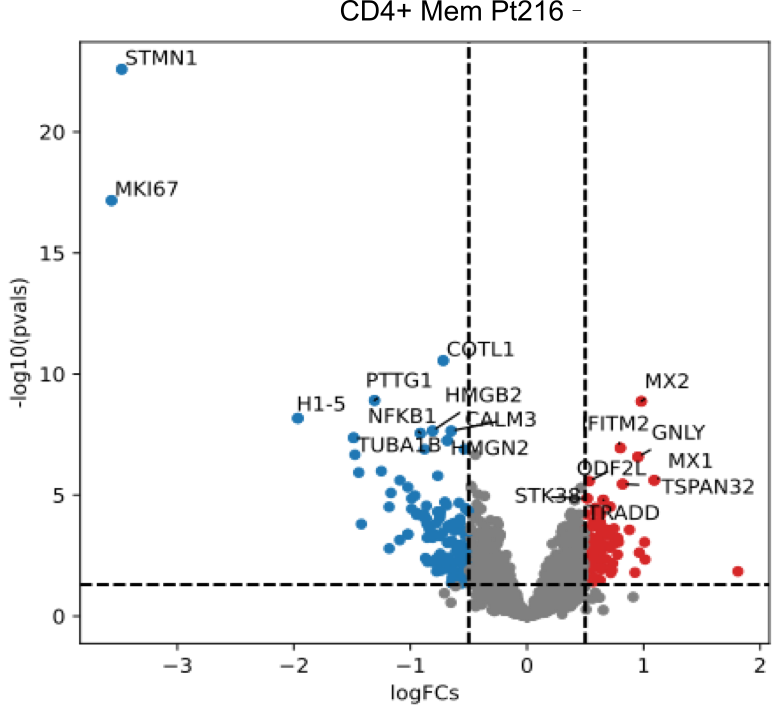

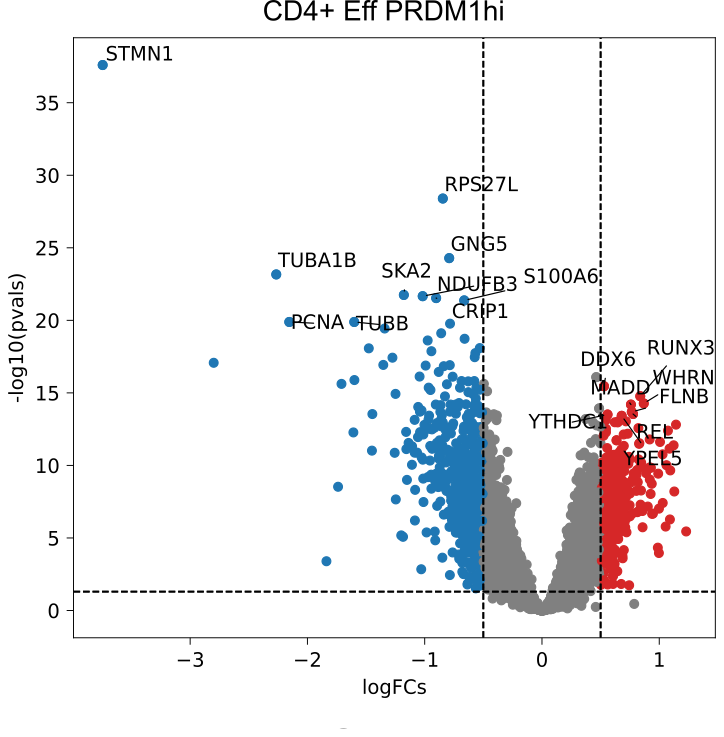

**Figure S13: CD4+ Mem BCL demonstrates low GZMK expression.** Volcano plots depicting differential gene expression in the CL enriched CD4+ CAR T cell clusters including **a,** CD4^+^ Mem BCL^hi^ **b,** CD4^+^ EOMES^hi^ **c,** CD4^+^ Eff PRDM1^hi^, and **d,** CD4^+^ Mem Pt216

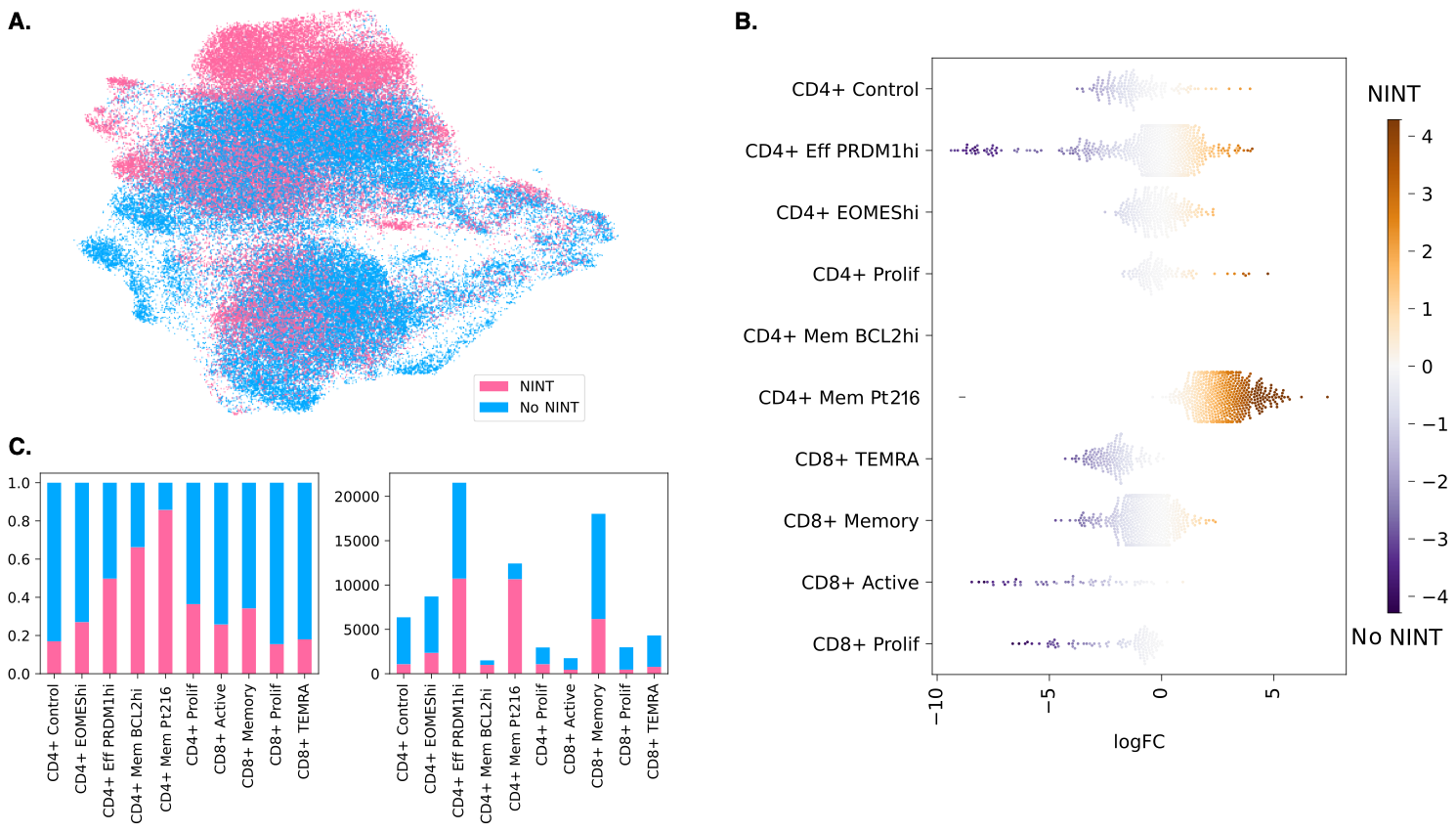

**Figure S14:** **Identifying NINT enriched clusters. A,** UMAP projection color coded identifying CAR T cell from control (blue) vs NINT (pink) patients. **b,** Abundance testing using Milo identifies clusters enriched in CAR T cell from control (blue) vs NINT (pink) patients. **c.** Relative (left) and absolute (right) proportion of CAR T cells from control and NINT patients comprising each cluster. CAR = chimeric antigen receptor; NINT = non-ICANS neurotoxicity; UMAP = Uniform Manifold Approximation and Projection

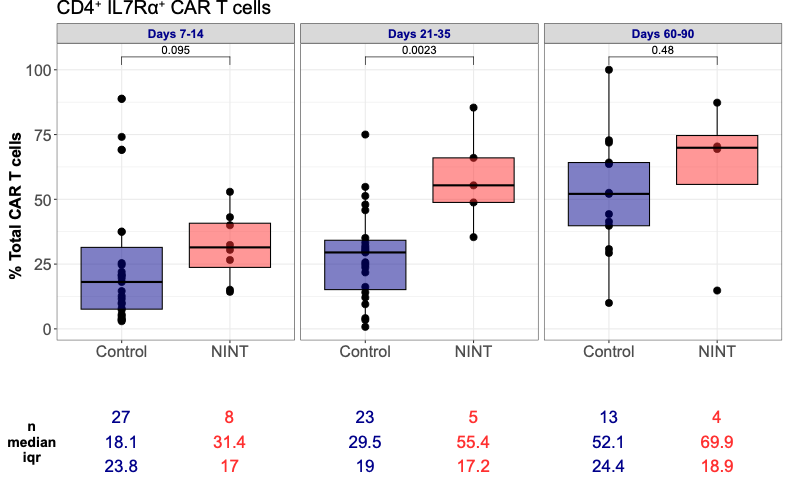

**Figure S15: Patients with NINT are enriched in CD4+ IL-7Rα^+^ CAR T cells.** CD4^+^ CD127/IL7rα^+^ CD4 CAR T cells measured by flow cytometry in control vs NINT patients over time

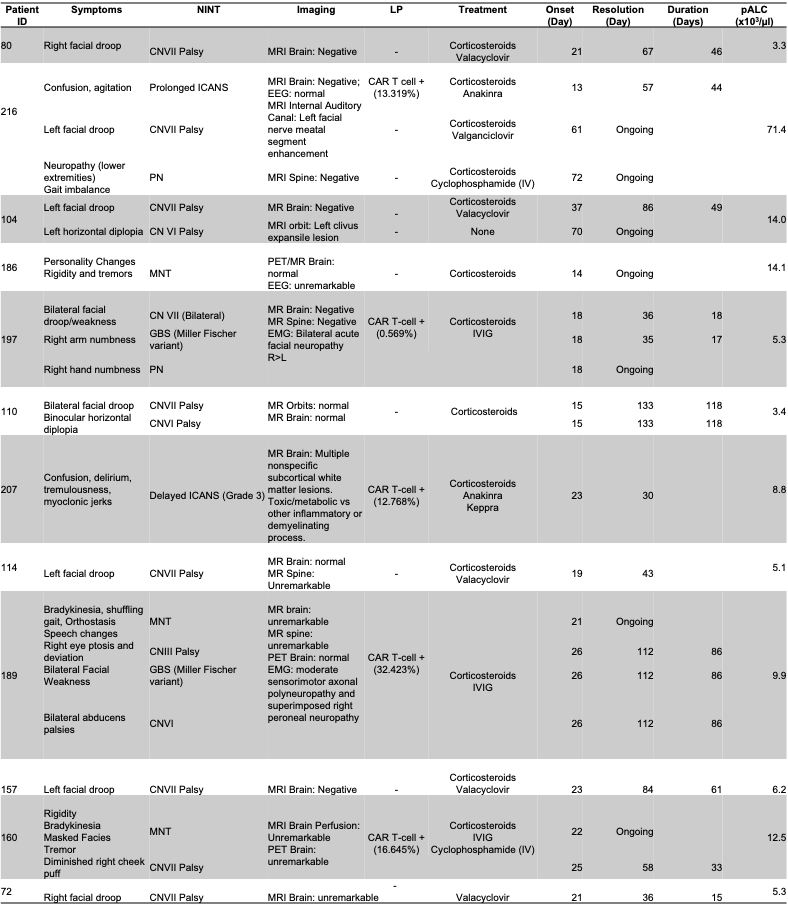
**Table S1: Clinical descriptions of patients diagnosed with NINTs**

CN = cranial nerve; PN = peripheral neuropathy; MNT = motor and neurocognitive toxicity; ICANS = Immune effector cell-associated neurotoxicity syndrome; GBS = Guillain-Barré Syndrome; pALC = peak absolute lymphocyte count (×10^3^/µL); MRI = magnetic resonance imaging; EEG = electroencephalography; EMG = electromyography; PET = positron emission tomography; CAR = chimeric antigen receptor; IVIG = intravenous immunoglobulin

**Table S2: Patient Responses by pALC**

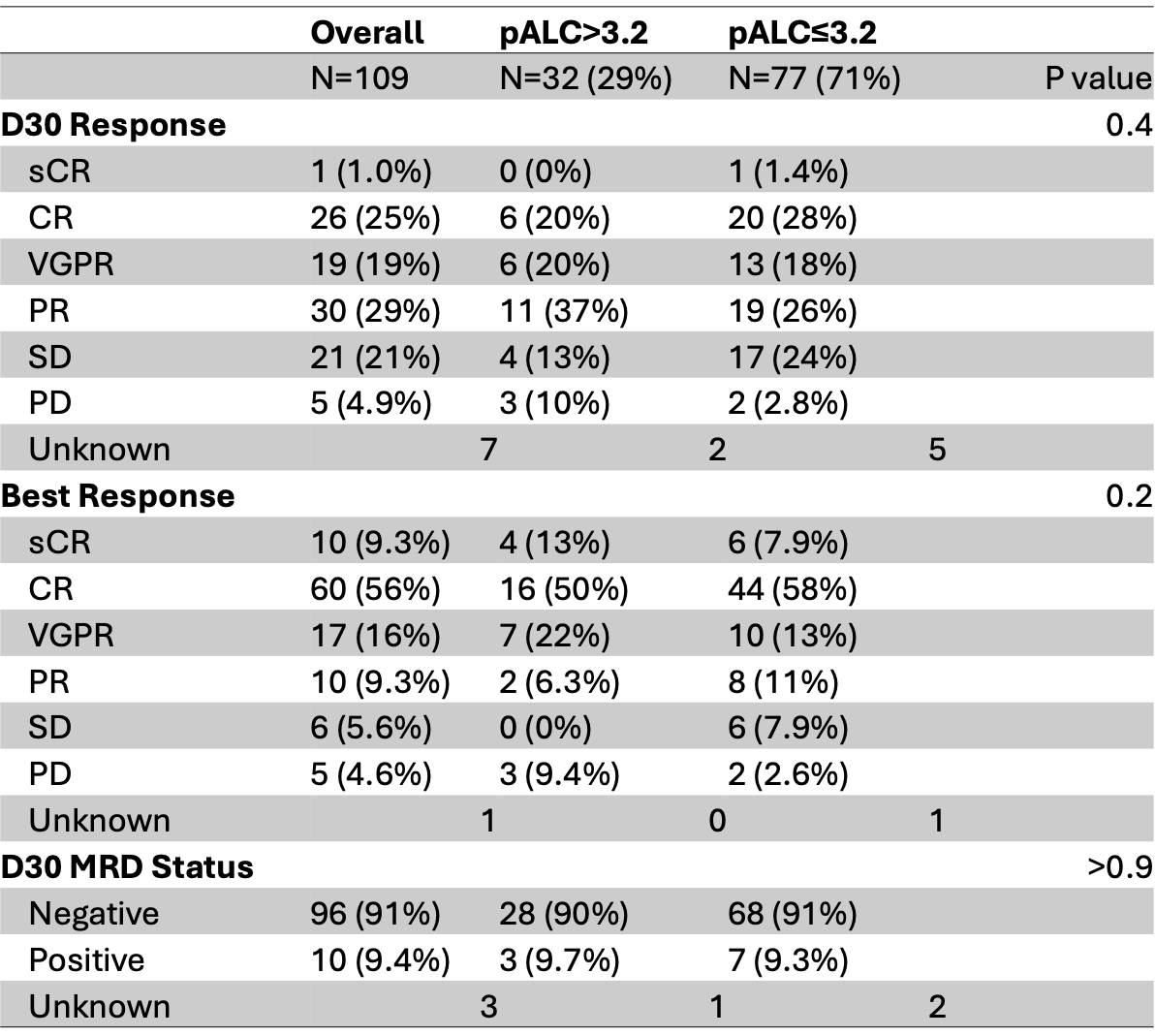

pALC = peak absolute lymphocyte count (×10^3^/µL); sCR = stringent complete response; CR = compete response; VGPR = very good partial response; PR = partial response; SD = stable disease; PD = progressive disease; MRD = measurable residual disease

**Table S2:** Olink Cytokine analysis comparing baseline cyotkines in patients with NINT vs without NINT

| **Cytokine** | **NO NINT** | **NINT** | **p value** |
| --- | --- | --- | --- |
| CXCL9 | 6.44706 | 7.08982 | 0.05221699 |
| MCP-1 | 10.33576 | 10.92089 | 0.05221699 |
| Ferritin | 196 | 73 | 0.06051209 |
| LIF | -0.74984 | -0.90812 | 0.08299485 |
| ARTN | -1.01268 | -1.33658 | 0.09400634 |
| IL2 | -1.15431 | -0.95348 | 0.09992216 |
| CX3CL1 | 4.03778 | 3.42702 | 0.11940403 |
| IL4 | -1.18381 | -1.32155 | 0.11940403 |
| GDNF | 0.74314 | 0.448575 | 0.1497077 |
| IL-12B | 4.77718 | 5.19003 | 0.19515583 |
| IL6 | 2.40729 | 3.135465 | 0.20531338 |
| TNFRSF9 | 3.31675 | 3.59836 | 0.20531338 |
| MCP-2 | 8.54823 | 9.1083 | 0.22673502 |
| LAP TGF-beta-1 | 5.47227 | 5.100775 | 0.23800681 |
| IL6 (clinic) | 8.4 | 14.8 | 0.25715741 |
| TNFSF14 | 4.15094 | 4.667035 | 0.2616878 |
| IL7 | 1.32281 | 1.14418 | 0.27410176 |
| CXCL10 | 7.16285 | 7.4143 | 0.28690047 |
| CD6 | 3.91529 | 4.486345 | 0.30008471 |
| IL-10RB | 6.47901 | 6.197435 | 0.30008471 |
| MMP-1 | 11.71449 | 11.59186 | 0.30008471 |
| IL-2RB | -1.71691 | -1.84806 | 0.31365516 |
| ST1A1 | 7.04409 | 7.657495 | 0.31365516 |
| 4E-BP1 | 4.03212 | 3.67976 | 0.32761142 |
| CCL25 | 5.60864 | 5.4569 | 0.32761142 |
| FGF-19 | 5.58329 | 5.082495 | 0.34195298 |
| CCL3 | 5.63451 | 5.911855 | 0.35667819 |
| LIF-R | 4.48242 | 4.34992 | 0.35667819 |
| NRTN | -0.88979 | -0.78654 | 0.35667819 |
| IL-1 alpha | -1.94166 | -2.1239 | 0.38727135 |
| FGF-5 | -1.49178 | -1.42897 | 0.41936693 |
| HGF | 8.79808 | 8.681595 | 0.41936693 |
| MMP-10 | 8.90675 | 8.61974 | 0.41936693 |
| IL10 | 1.15879 | 1.427215 | 0.45293043 |
| ADA | 4.6859 | 4.6422 | 0.47024901 |
| IFN-gamma | 4.7758 | 5.11191 | 0.47024901 |
| IL-15RA | 0.40687 | 0.31796 | 0.47024901 |
| IL-17C | 2.73512 | 2.885075 | 0.47024901 |
| AXIN1 | 1.87404 | 1.621545 | 0.48791665 |
| CCL23 | 9.49612 | 9.62463 | 0.48791665 |
| IL8 | 7.42152 | 8.109695 | 0.48791665 |
| TGF-alpha | 2.32871 | 2.35892 | 0.48791665 |
| IL-24 | 0.28859 | 0.40874 | 0.50592631 |
| STAMBP | 3.4087 | 3.11471 | 0.50592631 |
| IL-10RA | -0.87409 | -0.96175 | 0.52426974 |
| MCP-4 | 12.37129 | 12.68418 | 0.52426974 |
| EN-RAGE | 4.61523 | 4.84054 | 0.54293858 |
| IL-20RA | -0.76294 | -0.67559 | 0.54293858 |

| **Cytokine** | **NO NINT (0)** | **NINT (1)** | **NINT p value** |
| --- | --- | --- | --- |
| CCL19 | 9.24004 | 9.026865 | 0.581214 |
| TSLP | -2.41999 | -2.34424 | 0.581214 |
| NT-3 | -0.09911 | -0.05232 | 0.600801 |
| CD244 | 6.12923 | 6.29465 | 0.620672 |
| CXCL11 | 6.34506 | 6.720735 | 0.620672 |
| CXCL6 | 8.45699 | 8.40316 | 0.620672 |
| OPG | 10.06081 | 10.15936 | 0.620672 |
| SLAMF1 | 0.8856 | 0.808165 | 0.620672 |
| Beta-NGF | 0.98372 | 0.93332 | 0.640815 |
| FGF-23 | 0.89265 | 0.69153 | 0.640815 |
| TNFB | 1.44833 | 1.50359 | 0.640815 |
| TWEAK | 6.62303 | 6.847905 | 0.640815 |
| CRP | 0.92 | 0.625 | 0.646568 |
| CD40 | 10.16671 | 10.32006 | 0.661219 |
| Flt3L | 8.5773 | 8.48333 | 0.661219 |
| PD-L1 | 5.59082 | 5.318005 | 0.661219 |
| CASP-8 | 2.44082 | 2.900145 | 0.681871 |
| CD5 | 3.53403 | 3.658025 | 0.681871 |
| CST5 | 7.23626 | 6.97873 | 0.702757 |
| IL-18R1 | 6.52138 | 6.66923 | 0.702757 |
| MCP-3 | 3.01091 | 3.22214 | 0.723863 |
| IL18 | 8.64051 | 8.58075 | 0.766679 |
| TRAIL | 8.12324 | 8.157435 | 0.788359 |
| FGF-21 | 3.79883 | 3.636535 | 0.810199 |
| TRANCE | 3.46637 | 3.44092 | 0.810199 |
| uPA | 9.30282 | 9.16194 | 0.810199 |
| CCL20 | 5.79678 | 5.81122 | 0.832184 |
| OSM | 3.47845 | 3.791965 | 0.832184 |
| CCL28 | 1.70674 | 1.435615 | 0.854297 |
| CCL4 | 5.6956 | 5.917585 | 0.854297 |
| IL-20 | -1.62504 | -1.59977 | 0.854297 |
| sBCMA | 21.79 | 28.465 | 0.863441 |
| IL-22 RA1 | 0.89382 | 0.85851 | 0.881426 |
| IL13 | -0.22007 | -0.22024 | 0.898843 |
| IL33 | -1.02942 | -1.14242 | 0.898843 |
| CXCL5 | 10.06171 | 10.41415 | 0.9437 |
| CD8A | 5.45706 | 5.56206 | 0.966202 |
| SIRT2 | 2.90609 | 2.74975 | 0.966202 |
| TNF | 1.70341 | 1.69865 | 0.966202 |
| VEGFA | 11.56923 | 11.47753 | 0.966202 |
| CSF-1 | 7.8929 | 7.88905 | 0.988731 |
| DNER | 6.62452 | 6.608305 | 0.988731 |
| CCL11 | 6.34536 | 6.02626 | 1 |
| CDCP1 | 3.38156 | 3.41059 | 1 |
| CXCL1 | 8.20541 | 8.174085 | 1 |
| IL-17A | 0.74835 | 0.985205 | 1 |
| IL5 | 0.30251 | 0.324185 | 1 |
| SCF | 8.03648 | 7.991025 | 1 |

**Table S2 (cont.)**

**Table S3: Patient Responses by pALC**

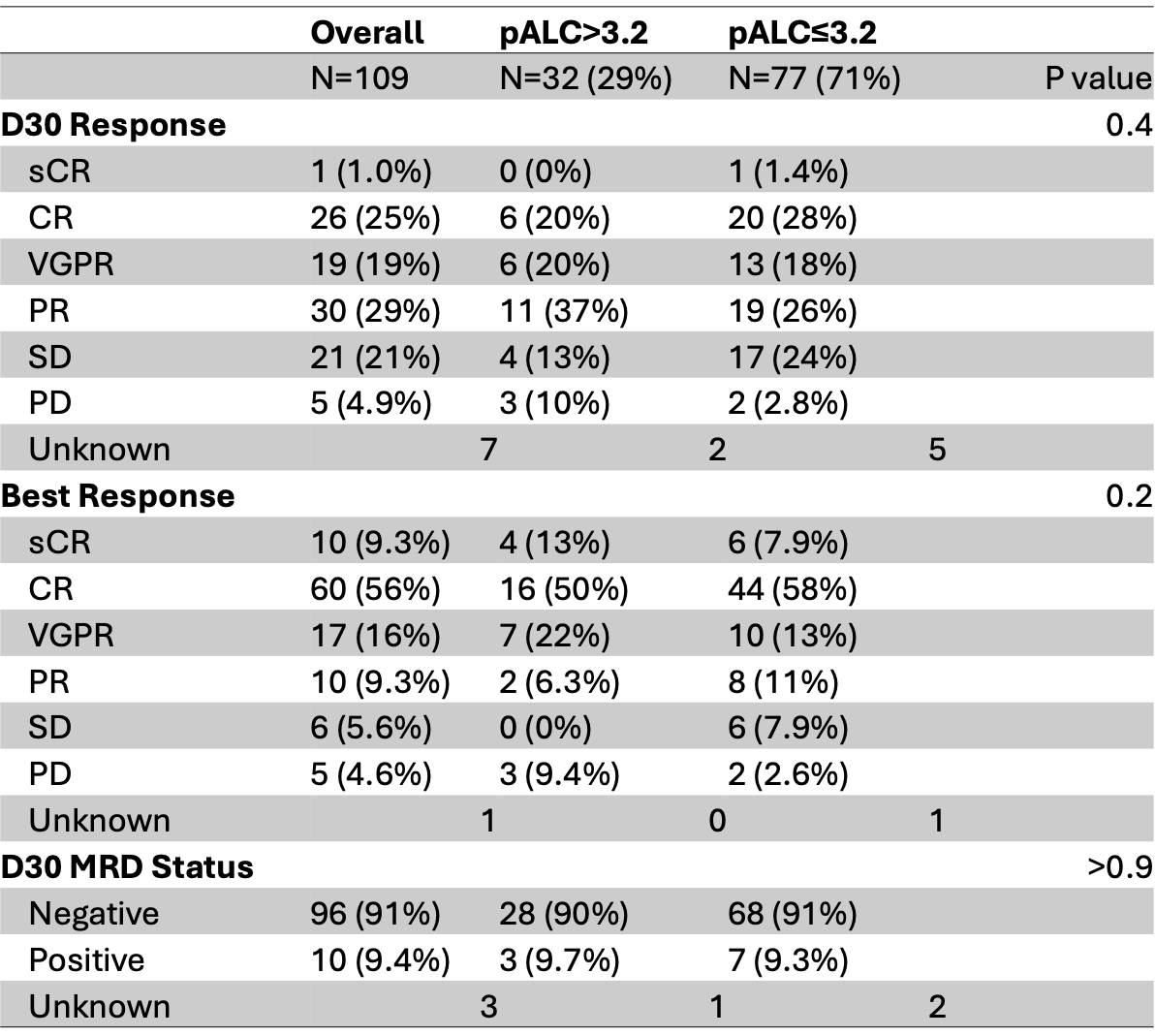

pALC = peak absolute lymphocyte count (×10^3^/µL); sCR = stringent complete response; CR = compete response; VGPR = very good partial response; PR = partial response; SD = stable disease; PD = progressive disease; MRD = measurable residual disease

**Table S4.** IDMS-022 CAR-T cell Screening Panel.

| Specificity | Clone | Fluorophore |
| --- | --- | --- |
| amine-reactive | n/a | LIVE/DEAD Blue |
| anti-BCMA | n/a | PE |
| Anti-GPRC5D | n/a | Alexa Fluor 647 |
| CD3 | UCHT1 | BUV496 |
| CD4 | SK3 | BUV737 |
| CD8 | RPA-T8 | BV570 |
| CD14 | MfP9 | BV711* |
| CD15 | W6D3 | BV711* |
| CD197 | 2-L1-A | BUV395 |
| CD45 | HI30 | BUV805 |
| CD45RA | HI100 | Alexa Fluor 594 |
| CD62L | DREG-56 | PerCP/Cy5.5 |

**Table S5.** IDMS T ExAct Panel.

| Specificity | Clone | Fluorophore |
| --- | --- | --- |
| amine-reactive | n/a | LIVE/DEAD Blue |
| anti-BCMA | n/a | PE |
| Anti-GPRC5D | n/a | Alexa Fluor 647 |
| CD3 | UHCT1 | BUV395 |
| CD4 | SK3 | cFluor V610 |
| CD8 | QA18A37 | Spark YG 593 |
| CD14 | 61D3 | Nova B 585 |
| CD25 | 2A3 | BUV563 |
| CD27 | O323 | BV570 |
| CD28 | L293 | BB700 |
| CD38 | HIT2 | APC/Fire810 |
| CD45 | HI30 | PerCP |
| CD45RA | HI100 | PE/Fire 700 |
| CD45RO | UCHL1 | FITC |
| CD57 | NK-1 | eF450 |
| CD69 | FN50 | Spark Blue 550 |
| CD95 | DX29 | BV480 |
| CD103 | BER-ACT8 | BV605 |
| CD127 | A019D5 | PE-Cy5 |
| CD134 | L106 | BV711 |
| CD137 | 4B4-1 | BUV661 |
| CD152 | L3D10 | PE-Cy7 |
| CD161 | DX12 | BV650 |
| CD178 | NOK-1 | BUV615 |
| CD197 | 2-L1-A | BUV496 |
| CD223 | T47-530 | APC-R700 |
| CD244 | C1.7 | BV510 |
| CD272 | MIH26 | PE/Dazzle 594 |
| CD278 | DX29 | BV421 |
| CD279 | EH12.2H7 | BV785 |
| CD366 | F38-2E2 | BV750 |
| HLA-DR | L243 | Spark Violet 538 |
| KLRG1 | 13F12F2 | BUV737 |
| TCR V7.2 | REA179 | APC-Vio770 |
| TCR Vδ1 | REA173 | PerCP-Vio700 |
| TCR Vδ2 | B6 | BUV805 |
| TIGIT | A15153G | PE/Fire810 |
